# The retina across the psychiatric spectrum: a systematic review and meta-analysis

**DOI:** 10.1101/2024.11.07.24316925

**Authors:** Nils M. Kallen, Giacomo Cecere, Dario Palpella, Finn Rabe, Foivos Georgiadis, Paul Badstübner, Victoria Edkins, Miriam Trindade, Stephanie Homan, Wolfgang Omlor, Erich Seifritz, Philipp Homan

## Abstract

The identification of structural retinal layer differences between patients diagnosed with certain psychiatric disorders and healthy controls has provided a potentially promising route to the identification of biomarkers for these disorders. Optical coherence tomography has been used to study whether retinal structural differences exist in schizophrenia spectrum disorders (SSD), bipolar disorder (BPD), major depressive disorder (MDD), obsessive-compulsive disorder (OCD), attention deficit hyperactivity disorder (ADHD), and alcohol and opiate use disorders. However, there is considerable variation in the amount of available evidence relating to each disorder and heterogeneity in the results obtained. We conducted the first systematic review and meta-analysis of evidence across all psychiatric disorders for which data was available. The quality of the evidence was graded and key confounding variables were accounted for. Of 381 screened articles, 87 were included. The evidence was of very low to moderate quality. Meta-analyses revealed that compared to healthy controls, the peripapillary retinal nerve fiber layer (pRNFL) was significantly thinner in SSD (SMD = -0.32; p<0.001), BPD (SMD = -0.4; p<0.001), OCD (SMD = -0.26; p=0.041), and ADHD (SMD = -0.48; p=0.033). Macular thickness was only significantly less in SSD (SMD = -0.59; p<0.001). pRNFL quadrant analyses revealed that reduced pRNFL thickness in SSD and BPD was most prominent in the superior and inferior quadrants. Macular subfield analyses indicated that BPD may have region-specific effects on retinal thickness. In conclusion, these findings suggest substantial retinal differences in SSD and BPD, reinforcing their potential as biomarkers in clinical settings.

## 1. Introduction

The efficacy of current pharmacological options for many psychiatric illnesses is limited. This is in part due to the heterogeneity with which these illnesses are presented and in part due to an incomplete understanding of their neurochemical bases [1–3]. The identification of biomarkers for psychiatric disorders offers potential for improving diagnosis, therapeutics, and prognosis.

In the search for such biomarkers, oculomics research examining the retina has gained traction in the last decade. The retina and the brain are embryologically and structurally similar. The retina is immuno-privileged, since it is shielded from external and internal threats by its blood-retinal barriers and an immune-suppressive microenvironment. Furthermore, it utilizes the same neurotransmitters and has a layered structure, similar to that of the brain. These features, in addition to the opportunity afforded by optical coherence tomography (OCT) to image the retina non-invasively, have led to the exploration of its use as a surrogate marker to understand or detect structural changes in the brain.

The retina captures incoming photons and transmits them as electrical and chemical signals along neural pathways to the occipital lobe, enabling the brain to create visual images. The retina comprises ten distinct layers. From innermost to outermost, these layers are the inner limiting membrane; the retinal nerve fiber layer (RNFL); the ganglion cell layer (GCL); the inner plexiform layer (IPL); the outer plexiform layer; the outer nuclear layer; the external limiting membrane; the photoreceptor layer; and the retinal pigment epithelium. The macula, the most visually sensitive area of the retina, sits at the center of the retina [4]. See Figure 1 for a simplified diagram of the retinal layers of interest.

**Figure 1.**
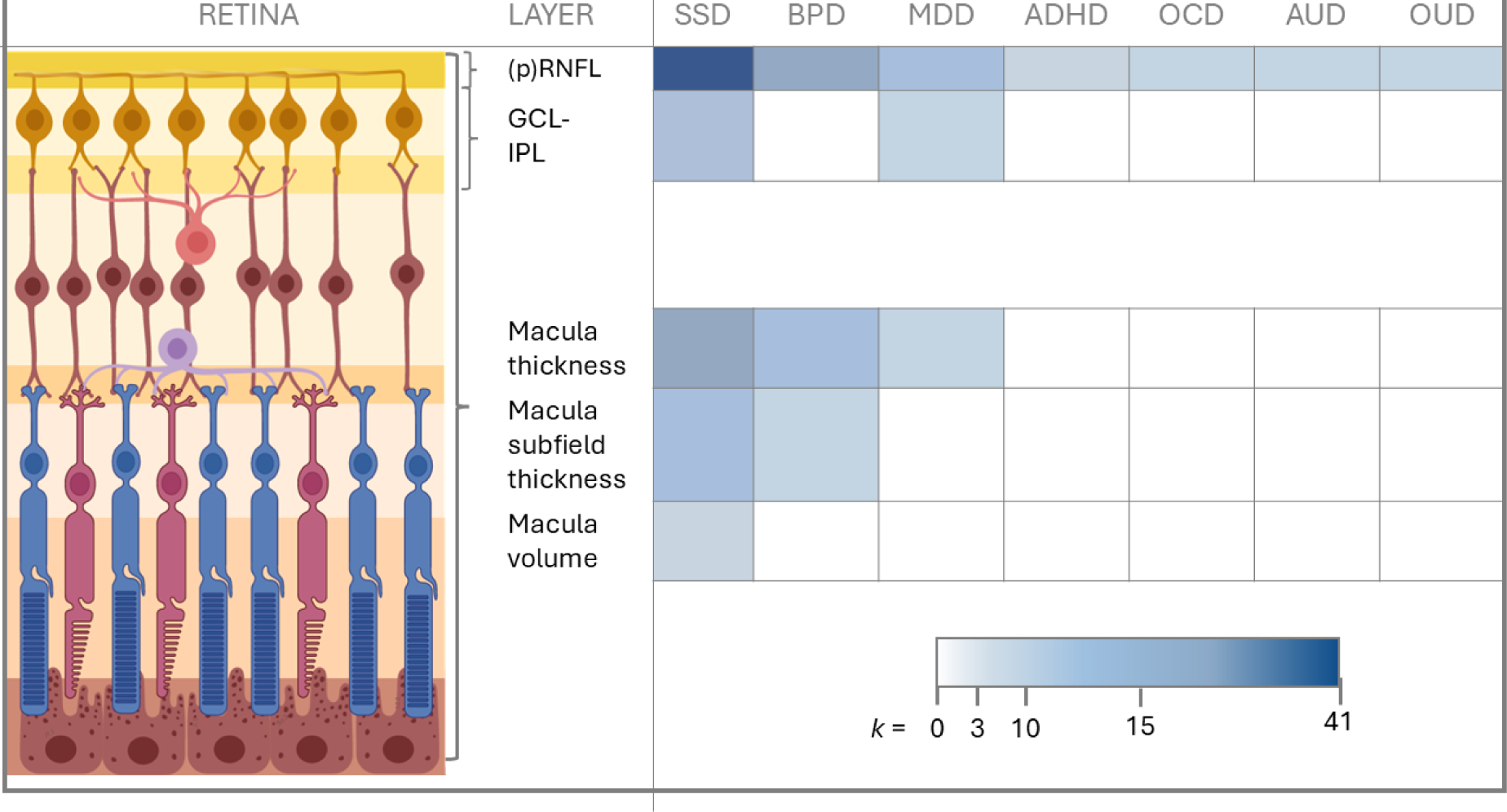
Retinal layers and data availability per diagnosis. Left part: cross-section of the retina in the macula region, centered around the fovea. Dark yellow cells = ganglion cells forming the retinal nerves with their axons. Light red cells = amacrine cells (immune cells). Dark red cells = bipolar cells. Purple cells = horizontal cells. Rods (blue) and cones (pink). Lowest layer shows cells of the retinal pigment epithelium. Right part: an overview of data availability graded on a blue scale: The lightest blue indicates the minimum number of studies (3) used for analyses. The darkest blue indicates the highest number (41). White boxes indicates that fewer than three studies were available and analyses was not performed.

Research across various neurological conditions, including multiple sclerosis [5], Parkinson’s disease [6–8], and Alzheimer’s disease [9], has found that thinning of retinal layers correlates with brain volume loss and disease progression. Similarly, the retina has emerged as a novel focus in psychiatric research.

In the last decade, an increasing number of studies have used OCT to investigate possible relationships between psychiatric diagnoses and measurements of retinal structures [10–96]. Most studies have focused on RFNL thickness, macular thickness (MT), or macular volume (MV) in people diagnosed with schizophrenia spectrum disorders (SSD) (see Supplementary (hereafter denoted as ‘S in table and figure references) Table S1). Fewer have examined retinal differences in patients with bipolar disorder (BPD) (Table S2) or major depressive disorder (MDD) (Table S3). A limited number of studies have considered other disorders such as attention-deficit/hyperactivity disorder (ADHD) (Table S4), obsessive-compulsive disorder (OCD) (Table S5), or substance use disorders such as alcohol use disorder (AUD) or opiate use disorder (OUD) (Table S6). Collectively, the findings of these studies have been inconclusive due to a lack of data with respect to some disorders and inconsistencies in the results obtained.

The heterogeneity of results obtained to date might be attributable to several factors. First, it could be explained by confounding factors such as patients’ ages, disease severity, disease stage (e.g. first episode or recurrent presentation), or smoking which is more common in patients with some psychiatric disorders than in the general population [97–100]. In the few studies that made retinal comparisons between different psychiatric disorders [11, 35, 58, 62], it was rare to account for the higher rates of comorbidities such as hypertension and diabetes in some illnesses compared to others [27, 41, 51]. Second, heterogeneity could have resulted from inconsistencies and differences in the measurements taken. While most studies reported peripapillary RNFL (pRNFL) thickness, some only reported macular RNFL thickness or did not specify the location of measurements (see Tables S18 - S20 for summary). Finally, the eye or eyes measured and how these measurements were reported varied considerably. Most studies reported values for each eye separately, but some reported the values for only the right eye or the mean for both eyes. In some cases, the laterality of the eye measured was not reported [33, 71] or the laterality of the eye measured was randomized [66, 93], despite significant differences between the right and left eyes in healthy controls [101, 102].

To prove useful as a diagnostic biomarker, different diagnoses would ideally show different retinal phenotypes. A study utilizing UK Biobank data [103] compared retinal thickness differences observed in SSD, BPD, and MDD. Deviations across diagnostic groups were driven by SSD and BPD with a significant difference in MT observed between the two. A systematic review and thematic synthesis of studies examining retinal differences in patients with SSD, BPD and MDD [104] found that abnormal findings were most common in SSD, followed by BPD, with no difference found in MDD. Whilst collectively these results could suggest retinal thinning to be a trait marker for SSD and BPD, the observational study designs used are not suitable to distinguish cause and effect for the observed parameters and are prone to bias in the selection of patients, controls and covariates. Clinical studies with a diagnostic design would help address the question of diagnostic accuracy and reproducibility. Unfortunately, such studies are lacking, as the area of clinical application (e.g. diagnosis, prognosis, assessment of disease progression or evaluation of therapeutic response) remains unclearly defined and, as highlighted, the question of whether psychiatric diagnoses present distinct RNLF and/or macular thinning profiles remains unanswered.

Meta-analysis provides an ideal means by which to attempt to clarify the existence of differential phenotypes across psychiatric disorders, enabling the utilization of studies with different foci and methodologies whilst simultaneously providing a more accurate prediction of effect size. Although meta-analyses carried out have provided valuable insights, methodological and scope-related limitations curtail their ability to provide a comprehensive understanding. Meta-analyses to date have included a limited number of psychiatric conditions, namely SDD and BPD [105, 106] or SDD, BPD and MDD [104]. Several studies [104–109] did not assess the certainty of evidence using the GRADE framework which would have helped to clarify the strength of conclusions drawn. In the one case where grading of the evidence was performed [110], it was done partially, without considering upgrading factors such as the magnitude of the effect or dose-response relationships. This would have limited the maximal quality of evidence achievable to “low”.

The aim of this systematic review and meta-analysis was to provide understanding of the retinal differences that can be observed across all psychiatric disorders for which data are available – SDD, BPD, MDD, ADHD, OCD, AUD and OUD. Key confounding variables (age, sex, disease duration and smoking status) and ocular laterality were accounted for, and a full GRADE assessment with consideration of upgrading factors was performed. The analysis provides a comprehensive overview of the available evidence relating to the existence of potential retinal biomarker candidates. It highlights both promising biomarker candidates and informs future research directions.

## 2. Methods

### 2.1 Study design and protocol

The current systematic review and meta-analysis is based on the Preferred Reporting Items for Systematic Reviews and Meta-Analyses (PRISMA) 2020 Statement and registered on the international prospective register of systematic reviews PROSPERO (https://www.crd.york.ac.uk/prospero/, ID: CRD42023442718) [111].

### 2.2. Eligibility criteria

We included all studies that met the following eligibility criteria: (1) a study population with any psychiatric diagnosis according to the International Statistical Classification of Diseases and Related Health Problems 10th Revision (ICD-10), or the Diagnostic and Statistical Manual of Mental Disorders (DSM) IV or V criteria; (2) inclusion of data from at least one OCT measurement; (3) inclusion of a healthy control group; (4) a minimum participant number of 10 per condition. Studies were excluded which (i) included samples with a mean age of <18 years; (ii) utilized post-mortem measurements; (iii) only reporting OCT angiography, choroidal layer, or optic cup/disk parameters; (iv) had not undergone peer review (e.g., conference abstracts and dissertations); (v) reported incomplete data; or (vi) exclusively used biobank data.

### 2.3 Search strategy and study selection

We systematically searched Embase, MEDLINE, and APA PsycINFO databases for articles indexed prior to June 2024, without language or publication date restrictions. The full search strategy is reported in Supplementary File 1. We carried out an additional, post hoc, non-systematic search on Google Scholar to check for further studies. We also performed a manual search of the reference lists of two relevant and recent reviews [104, 110].

### 2.4 Data extraction

We used a standard template to extract key information for all eligible studies: year of publication; country of research; inclusion and exclusion criteria; sample size, mean age and sex ratio of participants; diagnosis; duration of treatment and/or illness; chlorpromazine equivalents; participants’ smoking status; OCT device used; laterality of the measurements (left, right, or both); and retinal measurements included. Only baseline measurements were included if the study reported multiple longitudinal retinal assessments. In studies where multiple groups were compared (e.g., age groups [80] or clinical presentations [11, 16, 55, 71, 81, 89, 112]), we extracted data for all groups and the results were incorporated individually in the consecutive analyses. In studies that compared measurements from the same participants using two devices [55, 112], we used the exhaustive measurements that came from the most common device among all studies. We extracted the study with the larger sample size in case of partial participant overlap, or the study which contained the most participant details in the case of complete overlap. In studies where medication dosage was not reported in chlorpromazine equivalents [44, 58, 93], we used the formulas from Leucht et al. [113, 114] to convert them for comparability. Four authors independently extracted data for a blind check of accuracy.

### 2.5 Evaluation of Study Quality

The quality of each study included in our systematic review and meta-analysis was evaluated using the Newcastle-Ottawa Scale (NOS). [115] Higher scores on the NOS indicate better study quality. Two researchers independently assessed each study using the NOS. In cases where their assessments differed, they resolved the discrepancies through discussion to reach a consensus on the final NOS score. For study summaries including NOS scores see Tables S1 – S6.

### 2.6 Data analysis

Meta-analyses were performed for each retinal measure and diagnosis when data from at least three samples were available. When studies reported the means and standard deviations (SD) of retinal measurements separately for the left and right eyes, we calculated a combined mean and SD representing both eyes and used this for the main analyses. Meta-analysis of each retinal measurement was based on standardized mean difference (SMD) with a 95% confidence interval (CI). Pooled estimates were obtained by weighing each study according to a random-effects (RE) model. Heterogeneity across studies was evaluated according to standard cut-offs for I^2^ statistics. [116] Publication bias was assessed using Egger’s regression test [117] for correlates when data was available from at least 10 studies [118]. We used the trim-and-fill method for analyses which provided an Egger’s test score of p<0.10. For analyses with at least 10 studies which showed statistical significance (p<0.05) but high heterogeneity (I2 > 50%), we performed a sequential sensitivity analysis. The minimum number of studies required to reduce the I2 value below 50% were then removed and a further assessment carried out to determine whether the new estimates remained significant [119].

Influential studies were identified using differences in fits (DFFITS), differences in beta values (DFBETAS), cook’s distances, and hat values according to standard thresholds [120]. Influential studies were examined in detail to understand whether the results were due to specific bias, a measurement modality, or patient characteristics. The respective meta-analyses were repeated after influential study exclusion to assess their effect on the main results.

If a meta-analysis for a specific retinal parameter was statistically significant and included at least 10 samples, we conducted meta-regression analyses. These analyses explored the influence of factors such as age, sex, disease duration, and chlorpromazine equivalents, NOS ratings, and Positive and Negative Syndrome Scale for Schizophrenia (PANSS) scores on the retinal parameters.

All analyses were performed in R (4.2.3) using the metafor package [121].

### 2.7 Grading of the evidence

We used the standard GRADE items approach [122] for non-interventional observational studies, to classify the certainty of the evidence as high, moderate, low, or very low, for each variable showing a statistically significant estimate (p < 0.05). First, we evaluated the effect according to the standard cut-offs for SMD magnitude (0.2 small, 0.5 medium, 0.8 large) We downgraded or upgraded the certainty of the evidence by one level if the magnitude was small (SMD < 0.2) or large (SMD > 0.8) respectively. We also upgraded for a dose-response relationship if the meta-regression analysis assessing the effect of disease duration was significant.

Second, we assessed the impact of “risk of bias” on our findings by using meta-regression analyses to determine whether lower-quality studies, as identified by the NOS, were linked to greater differences in retinal measurements. The same approach was applied to account for potential biases due to age or sex differences between patients and controls. If any of these analyses revealed significant bias and the resulting controlled meta-analyses showed no significant results, we downgraded the quality of evidence by one level.

Third, we evaluated the precision of the effect estimates using a minimally contextualized approach [123]. We defined a minimal important difference, set at Δ=-0.2 for all retinal parameters, corresponding to an optimal information size of 800 participants (α=0.05; β=0.2). We then examined the upper CI of the pooled estimates and downgraded the certainty of the evidence if the upper CI reached or exceeded the minimal important difference. In the case that the upper CI did not encompass the minimal important difference, we downgraded by one level if the total sample was less than 50% of the optimal information size. In addition, we assessed the “consistency” of findings according to the I^2^ value. We downgraded the quality of evidence by one level if inconsistency was estimated (I^2^ ⩾ 50%) and the sequential between-study heterogeneity sensitivity analysis was non-significant.

Finally, we estimated the risk of “publication bias”, downgrading the quality of evidence by one level if fewer than 10 studies were included or Egger’s test p-value was <0.10, and the trim-and-fill method did not show significant differences in the retinal measurement. We thus labeled effect estimates as (i) high, (ii) moderate, (iii) low, or (iv) very low quality, depending on whether they (i) lay close, (ii) were likely to be close but possibly substantially different, (iii) might be substantially different, or (iv) were likely to be substantially different to the true effect [124]. The quality was addressed independently by three authors and any disagreement was resolved through discussion.

## 3. Results

### 3.1 Descriptive Statistics

Our search generated 605 records (328 from Embase, 188 from PubMed, and 89 from PsycINFO). After removing duplicates, 382 unique articles were screened. Following title and abstract review, 160 potentially eligible studies were retrieved for full-text assessment. Of these, 79 studies were excluded: 29 were grey literature, 21 lacked the predefined measures of interest, 10 included heterogeneous samples, 6 did not provide sufficient data for meta-analysis, 3 were Biobank studies, 2 lacked a healthy control group, 1 had fewer than 10 participants in one or more groups, and 7 had samples which overlapped with an included study. Additionally, from the reference list of Komatsu et al. (2024), seven relevant studies missing from our search were identified, of which one was excluded due to sample overlap. The remaining 87 studies were included in the meta-analysis [10–96]. The study selection process is presented in Figure S1. For an overview of the studies included, see Tables S1 - S6. Due to the limited number of studies in OCD, AUD, OUD and ADHD only restricted statistical analyses of publication bias using Egger’s test were possible.

Figure 1 summarizes the data available for the meta-analysis for each layer of interest and for each psychiatric diagnosis. The most complete data was available for SDD, followed by BPD and then MDD. For ADHD, OCD, AUD and OUD there were three studies for each disorder which reported pRNFL thickness; insufficient data pertaining to other layers was available. For an overview of data availability per study see Tables S18 – S20.

For pRNFL average thickness and pRNFL thickness in each of the four quadrants, contour-enhanced funnel plots and an Egger’s regression test showed no significant publication bias in studies of SSD (Figure S23 and S24 and Table S7**Fehler! Verweisquelle konnte nicht gefunden werden.**) or MDD (see Figures S36 and S37 and Table S9). In BPD, testing for influential studies revealed that Khalil et al. (2017) [22] substantially contributed to heterogeneity (LOO reduced I^2^ from 86.1% to 59.8% and estimated SMD from -0.50 to -0.40). Due to its influence on heterogeneity and the risk of it leading to an overestimation of effect size, the study was excluded from further analyses. For funnel plots and Egger’s test results for pRNFL average thickness and pRNFL thickness in each quadrant in studies of BPD, see Figures S31 and S32 and Table S8.

For MT and macular subfields, the contour-enhanced funnel plot indicates publication bias in MT (Egger’s test p=0.02) and central foveal thickness (Egger’s test p<0.01) in studies of SSD (Figures S25 - S28 and Table S7). Testing for influential studies and LOO revealed that the study by Sarkar et al. (2021) contributed considerably to heterogeneity in the central foveal subfield (LOO reduced I^2^ from 84.4% to 27.1%). Publication bias was non-significant after removing the study and re-calculating the regression models. For studies of BPD, funnel plots did not provide evidence of publication bias for MT or macular subfields (see Figure S33 - S35 and Table S8).

In relation to SSD: For MV, funnel plots and an Egger’s test did not point to publication bias (Figure S29). For GCL-IPL, there was evidence of a slight publication bias (Egger’s test p= 0.04), favoring a conservative estimate (Figure S30 and Table S7). Heterogeneity was low (I2 28.3%) and testing for influential studies did not reveal any that contributed considerably to heterogeneity.

### 3.2 Meta-Analysis

#### Comparative Results for pRNFL thickness

We used a RE model to calculate the pooled SMDs between pRNFL thickness for each disorder and healthy controls. Figure 2 displays these RE model results. A significant difference with a small-to medium-effect size was observed whereby thickness was less in SSD (SMD -0.32, CI -0.42 – 0.21, p<0.01) and BPD (SMD -0.40, CI -0.56 - -0.23, p<0.01) than in controls. We did not find evidence of significant pRNFL differences in MDD (SMD 0.09, CI -0.22 – 0.04, p=0.2). In OCD and ADHD the pRNFL was significantly thinner in patients than in controls (OCD: SMD -0.26, CI -0.5 – 0.01, p=0.04, ADHD: SMD -0.48, CI -0.92 - -0.04, p=0.03). No significant difference was found in AUD (SMD -0.45, CI -1.25 – 0.36, p=0.28) or OUD (SMD 0.11, CI -0.54 – 0.76, p=0.74). For individual forest plots see Figures S3 – S9).

**Figure 2.**
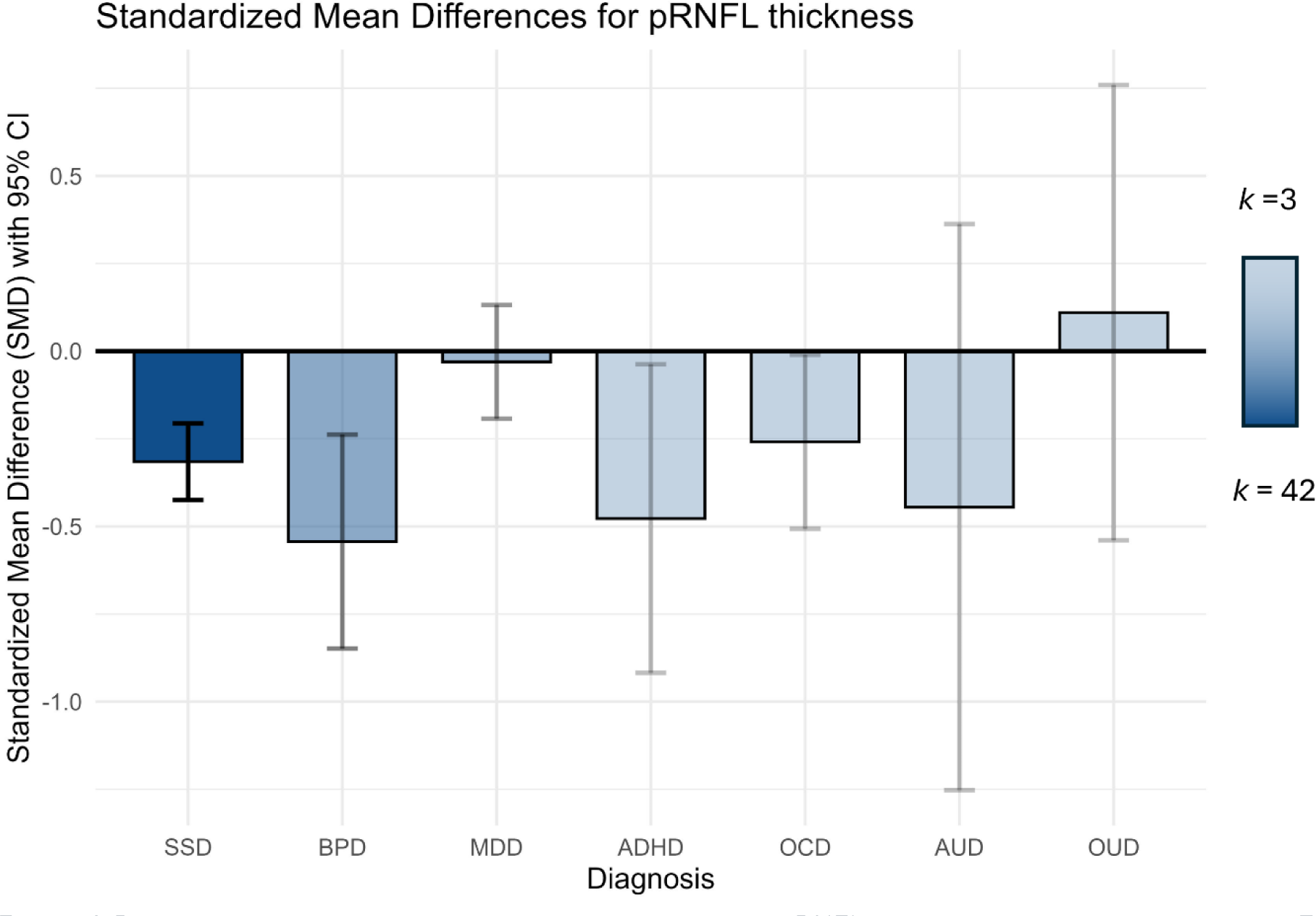
Random effects model results of the meta-analysis for pRNFL for different psychiatric diagnoses. Each bar plot shows the result of a meta-analysis of multiple studies with the estimated effect size as standardized mean differences and corresponding confidence intervals on the y-axis. Color transparency of bar plots and confidence intervals corresponds to the number of studies available for final analysis with higher color saturation, meaning a higher number of studies. SSD: 42 samples from 36 studies included; BPD: 16 studies included in the meta-analysis; MDD:, 10 samples from 9 studies included; ADHD: 3 studies available; AUD: 3 studies available and; OUD: 3 studies available.

Meta-regression analyses to evaluate the effect of covariates revealed a negative correlation between disease duration and overall pRNFL thickness and a slight positive correlation between chlorpromazine equivalents and overall pRNFL thickness. (See Tables S10 – S16.)

##### Comparative results for pRNFL quadrants

Figure 3 shows comparative SMDs for SSD, BPD and MDD. In SSD, all pRNFL quadrants were significantly thinner than in controls. The observed effect was strongest for the superior and inferior quadrants (superior: SMD -0.25, CI -0.38 - -0.13, p < 0.001; inferior: SMD -0.25, CI -0.38 - -0.13, p = 0.001) and less pronounced for the temporal and nasal subfields (temporal: SMD -0.20, CI -0.30 - - 0.09, p<0.001; nasal: SMD -0.14, CI -0.26 - -0.02, p = 0.027). A negative correlation between pRNFL thickness and disease duration was observed in the superior quadrant (mirroring the correlation observed for overall pRNFL thickness), but not in the temporal, inferior or nasal quadrants. A negative correlation with PANSS total score was observed in the nasal quadrant. See Tables S10 – S16, for results of meta regressions, and Figure S10 for the forest plots.

**Figure 3.**
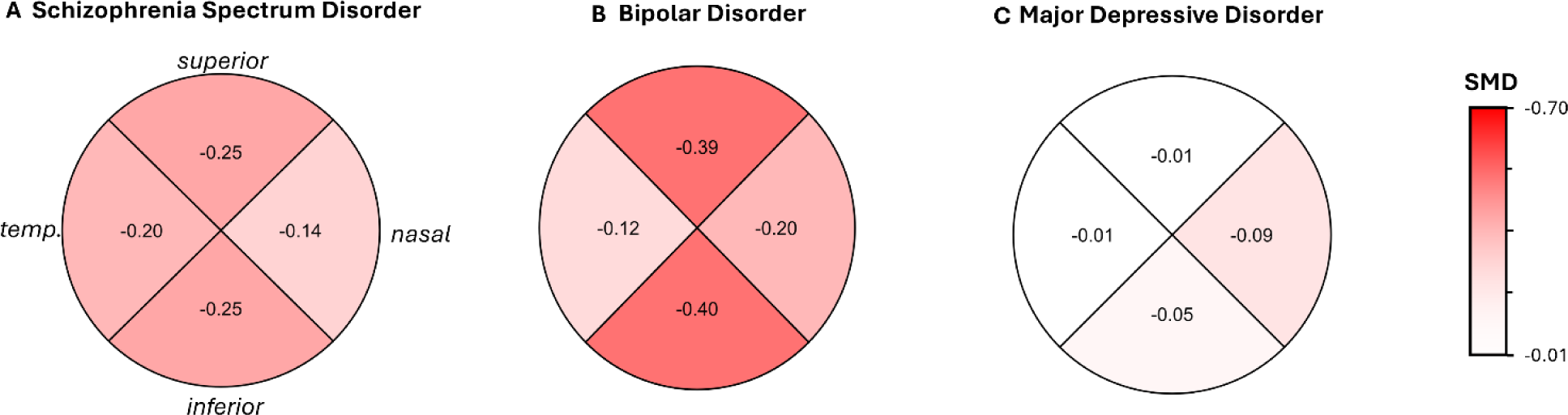
Random effects model results of the meta-analysis for pRNFL quadrants. Clockwise: superior (top), nasal (right), inferior (bottom), and temporal (left). (A) schizophrenia spectrum disorders, (B) bipolar disorder, and (C) major depressive disorder. Color intensity corresponds to effect size, with darker colors signifying more pronounced effects and lighter colors signifying less pronounced effects.

For BPD there were only significant differences in some quadrants: The effect sizes were greatest in the superior and inferior pRNFL quadrants (SMD -0.39, CI -0.52 - -0.25, p<0.001, and SMD -0.40, CI - 0.54 - -0.26, p<0.001 respectively), and comparatively small in the nasal quadrant (SMD -0.20, CI -0.39 - -0.01, p=0.04). There was no significant difference in the temporal quadrant (SMD -0.12, CI -0.28 - - 0.05, p= 0.169). (See Figure S11 for forest plots.)

We found no significant difference between patients and controls in any pRNFL quadrant measure in MDD (superior: SMD -0.01, CI -0.19 - 0.16, p = 0.87; inferior: SMD -0.05, CI -0.42 – 0.32, p=0.79; temporal: SMD -0.01, CI -0.30 – 0.29, p=0.96; nasal: SMD -0.09, CI -0.36 – 0.18, p=0.48). For forest plots see Figure S12.

##### Comparative Results for Overall Macula and Subfields

MT was significantly less in SSD than in controls (SMD -0.59, CI: -0.78 - -0.39, p<0.01); this effect size was medium. No significant difference was found for BPD (SMD -0.2, CI -0.51 – 0.10, p=0.18) or MDD (SMD -0.05, CI -0.26 – 0.16, p=0.64). For forest plots, see Figures S13 – S15. Meta-regression analysis showed a positive correlation of MT differences with age group differences between patient and control groups for SSD. This indicates a possible selection and recruiting bias (see Tables S10 – S16). For BPD and MDD, there were insufficient studies for meta-regression.

We also found that all macular subfield measurements for SSD were significantly less than for controls. This was the case for both the inner (superior: SMD -0.57, CI -0.87 - -0.28, p<0.001; inferior: SMD - 0.52, CI -0.73 - -0.31, p <0.01; temporal: SMD -0.42, CI -0.61 - -0.22, p<0.01; nasal: SMD -0.49, CI - 0.74 - -0.25, p<0.001) and outer rings (superior: SMD -0.57, CI -0.87 - -0.28, p<0.01; inferior: SMD - 0.52, CI -0.73 - -0.31, p<0.01; temporal: SMD -0.42, CI -0.61 - -0.22, p<0.01; nasal: SMD -0.49, CI - 0.74 - -0.25, p<0.01).

Only three studies were available for macular subfield analysis in BPD. Of the inner ring macular subfields, the superior, nasal and temporal subfield measurements were significantly less in patients than controls (superior: SMD -0.43, CI -0.85 - -0.01, p=0.04; nasal: SMD -0.44, CI -0.63 - -0.24, p<0.01; temporal: SMD -0.47, CI -0.67 - -0.28, p<0.01). There was no significant difference for the inferior subfield (SMD -0.36, -0.76 – 0.03, p=0.07). For the outer ring macular subfields, only the superior subfield was significantly less than in controls (SMD -0.70, CI -0.95 - -0.45, p <0.01). There were no significant differences in the nasal, inferior, and temporal subfields (nasal: SMD -0.24, CI -0.67 – 0.20 p=0.29; inferior: SMD -0.13, CI -0.63 – 0.37, p=0.61; temporal: SMD -0.21, CI -0.66 – 0.24, p=0.35). For an overview of macular differences in SSD and BPD, see Figure S16. For individual forest plots for macula subfields for SSD and BPD, see Figures S17 – S20.

For MV, sufficient data was only available to conduct a meta-analysis for SSD. The difference between patients and controls was significant with a medium effect size (SMD -0.52, CI -0.67 - -0.37, p<0.01). See Figure S21.

##### GCL-IPL

Research on combined GCL-IPL was limited, with data available only for SSD and MDD. For SSD, a significant thickness difference was observed (SMD -0.40, CI -0.57 - -0.23, p<0.01). In MDD the difference was almost significant (SMD -0.24, CI: -0.48 - 0.01, p=0.06).

### 3.3 Grading of the evidence

According to the GRADE assessment, no evidence was deemed to be of high quality. The strongest evidence was for SSD, which demonstrated moderate certainty for overall pRNFL thinning. The evidence for SSD was of low-quality for all macular measurements (10 out of 10) and of low or very low-quality evidence for the pRNFL quadrant measurements (two each). For BPD, most evidence was rated as “very low” for most measurements. For only three measurements (overall, inferior, and superior RNFL thinning in both eyes) was the evidence rated as “low”. The other diagnostic groups (MDD, OCD, ADHD, AUD, and OUD) consistently showed very low-quality evidence across all statistically significant results. The downgrades were primarily due to imprecise CIs and publication bias resulting from the small number of studies. Smaller effect sizes and inconsistency due to the limited studies also contributed to the downgrades, although to a lesser extent. A small subset of the SSD meta-analyses received upgrades due to a dose-response relationship with disease duration, but no upgrades were made based on a large magnitude of effect. A detailed GRADE assessment report is available in Table S17.

## 4 Discussion

Oculomics is a relatively new and rapidly growing field in psychiatric and neuroscience research that aims to identify ophthalmic biomarkers for systemic disorders. Unfortunately, the limited presence of, and adherence to reporting guidelines has led to a disorderly body of research. Measurement protocols are often inadequately described, and generated data is often incomplete and lacks control for key confounders. Our goal was to disentangle the currently available data to investigate how retinal layers are affected in different psychiatric disorders.

Our meta-analysis reveals significant retinal differences between patient and control groups across various psychiatric conditions, with the most pronounced effects observed in SSD and BPD. These findings go beyond previously published meta-analyses: By including a broad range of psychiatric disorders and adhering to guidelines for evidence synthesis and quality grading, we have provided new neurobiological insights across the psychiatric spectrum.

### Overall pRNFL and pRNFL quadrants

The strongest evidence of a difference in pRNFL thickness between patients and controls was observed for SSD. Analyses of individual pRNFL quadrants also revealed a pattern of thinner inferior and superior quadrants in SSD compared to controls. However, it should be noted that effect sizes were in the small to medium range. There is also evidence of a negative relationship between SSD disease duration and pRNFL thickness. This finding is consistent with studies which have evidenced that thickness differences are not typically present in first-episode patients but are common in chronically ill individuals [43], [48]. The thinning effect could be associated with the neurodegenerative aspects of schizophrenia, reflecting and potentially predicting broader neural deficits. In vivo imaging studies have shown significant brain structural changes in schizophrenia, with consistent findings of reduced cortical grey matter volume and cortical thinning [125, 126] which might also alter retinal structures via retrograde trans-synaptic degeneration [127]. It is also possible that synaptic loss, rather than changes in neuronal or glial numbers might underlie these structural alterations [128–132].

Consistent with previous meta-analyses [104–106], it was found that pRNFL differences also occur in BPD. Although the certainty of the evidence is lower than that for SDD, the size of the difference observed between patients and controls is greater. Analyses of individual pRNFL quadrants in BPD also revealed a pattern of thinner inferior and superior quadrants compared to controls.

While lower statistical accuracy due to the availability of fewer studies could explain the seemingly greater difference observed in BPD than in SSD, there could also be a role for underlying pathomechanisms. As in SSD, dendritic spine loss has been observed in BPD [129] and genetic studies have revealed a significant genetic overlap between BPD and SSD [129, 130]. Disturbances in circadian rhythm are common in BPD and indicate relevant disturbance in the retinohypothalamic pathway responsible for the sleep-wake cycle. [133–136]. The additional degeneration of axons forming the retinohypothalamic pathway could explain the more pronounced effect observed in BPD.

There was no evidence of either overall pRNFL or pRNFL quadrant thickness differences between MDD patients and controls. Our findings relating to the pRNFL in SSD, BPD, and MDD align with research using the UK Biobank dataset, which also identified retinal differences in SSD and BPD, but not MDD, using a normative modeling approach [103].

There is evidence that ADHD and OCD are also associated with a thinner overall pRNFL. However, the effect sizes were small. Retinal thinness in OCD may reflect broader neurobiological alterations associated with this condition, potentially including neurodegeneration or inflammatory processes.

There is evidence that the pRNFL thickness differences evidenced for ADHD in the adult population in our meta-analysis are not apparent for the adolescent population [108]. The finding of pRNFL thickness differences in adults with ADHD, but not in adolescents seems counter to the current hypothesis of delayed cortical maturation in ADHD; the reasons for this discrepancy remain unclear. A possible explanation may relate to the effects of prolonged exposure to dopaminergic medication. Tuenel et al. [68] found the largest effect in older patients (mean age 33) who used medication continuously. In contrast, Kaymak et al. [57] observed a smaller, yet significant, difference in younger patients (mean age 23.21) who had been off medication for at least a year. Surprisingly, Erdogan et al. [54] found no difference in thickness measurements in patients on medication for at least six months, suggesting other disorder-specific factors may influence these outcomes.

There was no evidence of differences in overall pRNFL thickness between those with either alcohol or opiate use disorder and controls. pRNFL quadrant analysis did provide evidence of differential patterns in AUD and OUD: In AUD the temporal pRFNL was significantly thinner than in controls but the nasal pRNFL was not. Conversely in OUD this pattern was reversed. While this may suggest that retinal effects are specific to the substance of abuse, the body of research is very small, we do not have data pertaining to all subfields, and the there is considerable lack of consistency in the results. Research is also complicated for these disorders because of the difficulty in disaggregating the neurobiological bases and the substance-based effects. Before further studies in this field are undertaken, clear justification based on the potential for benefits and applications should be given.

### Overall Macular and Subfields

Overall MT was significantly less than controls for SSD but not for BPD or MDD. The possibility to further differentiate between SSD and BPD is evidenced by the macular subfield analyses: In SSD all macular subfields were significantly thinner than in controls, whereas in BPD only five of nine subfields were significantly thinner (although statistical power in this case was lower than for SSD). It is possible however that these findings were spurious and a consequence of the small number of studies included in the subfield analysis. One alternative possibility is that the differential observations in SSD and BPD relate to different rates of smoking.

Smoking is a risk factor for macular thinning [137] due to its effects on microvasculature. The smoking rate is higher in patients with SSD than in those with BPD [99, 100] and studies show impaired brain and retinal microvasculature and disruption of blood-brain barrier in SSD [138, 139]. While retrograde degeneration supposedly only effects the RNFL and GCL, processes such as loss of dendritic spines, inter-neural connection and aberrant inflammation could affect all retinal layers, therefore causing or contributing to the differences observed between SSD and BPD. More comprehensive evidence from larger and more detailed clinical studies is needed to assess if the subfield differences evidenced in this meta-analysis can be reproduced in larger scale studies or were due to variabilities within previous studies, confounding factors or small sample sizes.

For MDD, subfield analysis was only possible for the central subfield and no significant difference was found. Further research is needed to ascertain whether MT differences are entirely absent for MDD or they are in fact confined to specific subfields.

### Ganglion Cell-Inner Plexiform Layer (GCL-IPL) Thickness

Research on combined GCL-IPL was limited, with selected data available only for SSD and MDD. While a statistically significant difference was observed in SSD, the difference observed for MDD was almost statistically significant (p=0.055), but the overall certainty of evidence was very low. Overall, these findings suggest that GCL-IPL thinness may be a marker which could be used to differentiate psychotic (e.g. SSD) and affective (e.g. MDD) disorders. Indeed, this hypothesis is supported by two recent biobank studies [140, 141] which found an association between polygenic risk scores for schizophrenia in healthy controls and GCL-IPL thickness. However, with only limited studies reporting on combined GCL-IPL or its constituent parts, there is a need for further research to further test this hypothesis and its clinical utility [103, 133, 134].

### Limitations and Future Directions

This meta-analysis primarily used averages of data from left and right eyes to optimize statistical strength. This is because a large portion of studies only reported pooled results. While we additionally analyzed separate right and left eye data wherever possible, overall evidence was weaker due to low study numbers. Given evidence that laterality analysis may offer insights which are not apparent from pooled data [101, 102], future research should consider reporting interocular differences separately.

Also, we grouped all schizophrenia spectrum disorders into one group, comprising a variety of first episode, chronic and treatment resistant subgroups and various syndromes. Thus, a promising area of research may be to focus on retinal differences in specific psychotic syndromes such as schizoaffective or brief psychotic disorders.

Contour-enhanced funnel plots and Egger’s tests were used to ward against publication bias. However, these tools are not perfect, especially with low study numbers. We tried to limit heterogeneity by excluding influential studies that contributed excessively to the estimated effect, and still observed moderate heterogeneity. Whether this is due to variance in relevant covariates such as different clinical subgroups or health-related variables such as body mass index (BMI) should be explored in future research. Unfortunately, to date, studies have been inconsistent in the reporting of disorder-related covariables and potential confounders. Consequently, visual acuity, BMI, level of education, and ocular pressure could not be included in the meta-regression analyses.

The combined use of different OCT device types and layer segmentation software is also limiting because different devices with variances in resolution could provide differing layer measurements. While we conducted meta-regression to examine these effects, studies did not consistently report technical characteristics such as center wavelength and wavelength range, speed of acquisition, number of horizontal and vertical scan lines, raster or radial scan, the dimensions of area of interest mapped and where said area was centered (e.g. fovea or optic cup). While reporting recommendations for OCT studies do exist, for example, the APOSTEL-Recommendations, few completely or even partially adhered to them.

This meta-analysis focused on the retinal structures that have to date received the most attention in psychiatric literature. Future research could broaden the focus to examine for example, the choroid layer, retinal vascularization or electroretinography.

## 5 Conclusion

Our findings suggest that SSD and BPD are associated with a thinner pRNFL while MDD is not. Differences in MT appear to further enable a differentiation between SSD and BPD, as reduced thickness is only observed in the former. For OCD, ADHD, AUD, and OUD, as results remain inconsistent and based on minimal data, more research is needed clarify the potential of retinal markers to act as biomarkers for these disorders.

We need quantitatively more and qualitatively better evidence for the whole spectrum of psychiatric disorders. The range of focus should be extended to other clinically relevant psychiatric conditions such as schizotypal disorders, borderline personality disorder and autism spectrum disorder.

Retinal structural differences observed via OCT show promise as biomarkers to distinguish psychosis-associated endophenotypes from other psychiatric disorders and could therefore serve as a useful adjunct in clinical decision making. Future research should explore the utility of the retinal differences observed as biomarkers in real-world clinical settings, as findings from controlled environments may not always translate to practice. Studies directly comparing patient groups are also necessary to validate discriminatory accuracy.

## Data Availability

All data produced in the present study are available upon reasonable request to the authors

