## Supplementary File for "The retina across the psychiatric spectrum: a systematic review and meta-analysis"

**Full search strategy and PRISMA-Flowchart**

Search date: 31st May 2024.

Publication date limits: none.

Language restrictions: none.

Database: Embase, MEDLINE, APA PsycInfo

Search string:

((adhd OR attention deficit OR asd OR autis* OR schizophren* OR psychos* OR bd OR bipolar disorder OR mdd OR depress* OR ocd OR obsess* OR substance abuse OR schizotyp*) AND (optical coherence tomography OR retinal nerve fiber layer thickness OR RNFL OR macula volume OR macular thickness OR ganglion cell layer OR choroidal layer OR Outer nuclear layer OR Inner nuclear layer)) AND (psychiatr* OR dsm)

**Supplementary Figure 1 PRISMA-Flowchart**

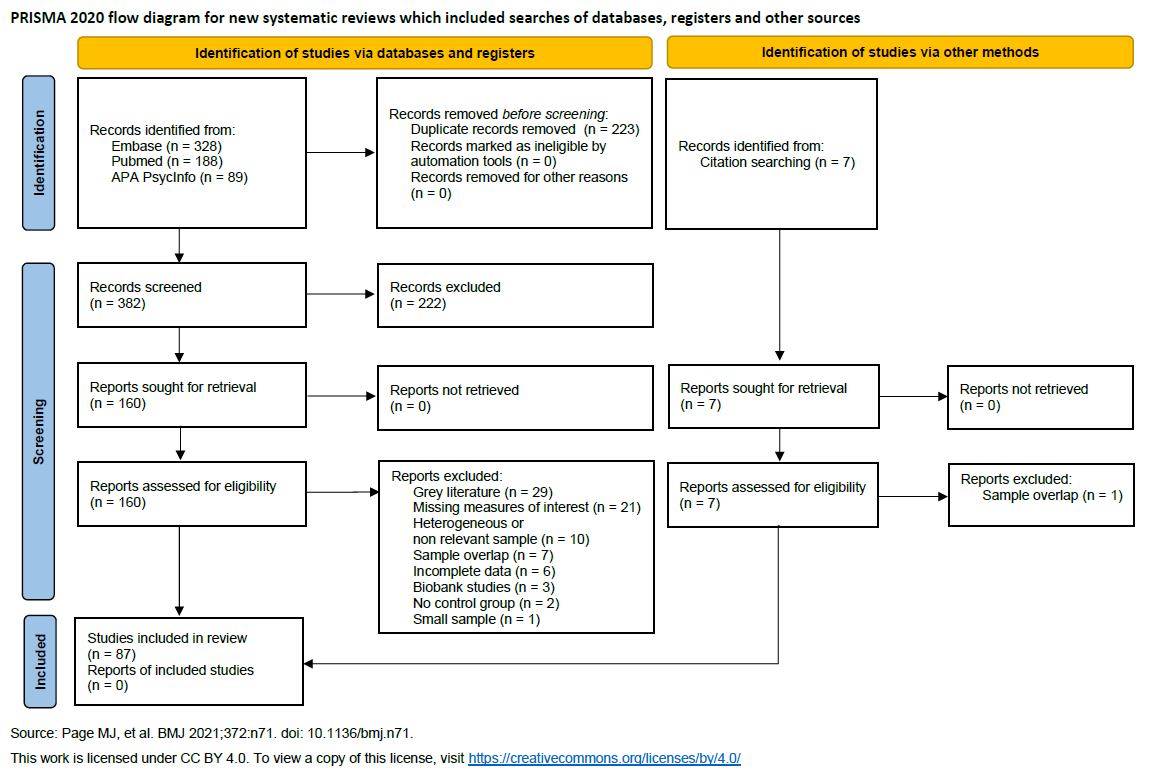
*Figure S 1 Flow Diagram of the systematic review and meta-analysis according to PRISMA-Guidelines*

**Supplementary Tables 1 – 6 Details of Studies Identified Relating to Each Psychiatric Disorder**

*Table S 1 we found 41 studies investigating retinal layers in schizophrenia spectrum disorder. N = number of patients / controls. NOS = Newcastle-Ottawa-Scale score, DoD = Duration of disease .Blank spaces = variable was not reported. Domagala et al compared three different age groups. R+L Left and Right eye reported separately. RL: only pooled values for both eyes reported.*

| Study Characteristics – Schizophrenia Spectrum Disorder | | | | | | | | | | |
| --- | --- | --- | --- | --- | --- | --- | --- | --- | --- | --- |
| **Authors** | **Year** | **Country** | **NOS** | **Age patients (Mean±SD)** | **Age controls (Mean±SD)** | **DoD  (Mean ±SD)** | **N Patients** | **N Controls** | **Laterality** | **Device** |
| Alizadeh M. et al | 2021 | Iran | 8 | 38.26 ±7.90 | 36.4±9.51 | 15.33 ±9.65 | 30 | 15 | R+L | ZEISS |
| Altun IK. et al | 2020 | Turkey | 8 | 44.20 ±10.02 | 39.35±4.27 | 20.3 ±8.91 | 35 | 31 | R+L | OPTOVUE |
| Asanad S. et al | 2021 | USA | 6 | 37.20 ±12.30 | 41.1 ±15.2 |  | 58 | 35 | RL | ZEISS |
| Ascaso F.J. et al | 2010 | Spain | 7 | 39.20 ±13.50 | 39.5 ±13.6 |  | 10 | 10 | RL | ZEISS |
| Ascaso FJ. et al | 2015 | Spain | 7 | 45.10 ±10.40 | 44.5 ±10.9 | 10.9 ±16.3 | 30 | 30 | R+L | ZEISS |
| Bannai D. et al | 2020 | USA | 7 | 36.20 ±13.20 | 39 ±12.6 | 11.88 ±13.47 | 25 | 15 | R+L | HEIDELBERG |
| Boudriot E. et al | 2023 | Germany | 9 | 39.29 ±10.80 | 34.65 ±11.35 | 13.69 ±7.81 | 65 | 72 | RL | HEIDELBERG |
| Boudriot E. et al | 2024 | Germany | 9 | 39.08 ±10.48 | 33.58 ±11.85 | 13.23 ±8.75 | 98 | 125 | R+L | ZEISS |
| Bozali E. et al | 2022 | Turkey | 6 | 37.20 ±9.90 | 36.8 ±9.6 |  | 57 | 57 | RL | NIDEK |
| Budakoglu O. et al | 2021 | Turkey | 5 | 40.80 ±7.90 | 45.2 ±8.1 | 8.3 ±3.3 | 22 | 26 | R | OPTOVUE |
| Carriello MA. et al | 2023 | Brazil | 8 | 36.51 ±12.41 | 36.74 ±11.94 |  | 35 | 35 | R+L | HEIDELBERG |
| Celik M. et al | 2016 | Turkey | 7 | 35.37 ±9.69 | 35.49 ±16 | 12.04 ±9.87 | 41 | 41 | R | HEIDELBERG |
| Chu EM. et al | 2012 | UK | 6 | 29.10 (Not reported  ) | 29.5 ±6.12 | 4.4 ±3.6 | 11 | 40 | R+L | HEIDELBERG |
| Daneshvar R. et al | 2024 | Iran | 7 | 35.86 ±9.29 | 34.36 ±6.56 |  | 22 | 22 | random | Not reported |
| Delıbaş D.H. et al | 2018 | Turkey | 7 | 39.84 ±11.82 | 40.13 ±9.03 | 13.47 ±10.86 | 19 | 19 |  |  |
| Domagała A. et al | 2023 | Poland | 7 | 60.11±5.24^a^/  38.35 ±6.24^b^/  25.50 ±3.67^c^ | 57.22±2.42^a^ /  42.5±3.36^b^ /  28±3.91^c^ | 31.47±11.47^a^/  15.91±8.33^b^ /  5.53±3.81^c^ | 19^a^ / 21^b^ /  20^c^ | 19^a^ /  19^b^ /  22^c^ | R+L | OPTOPOL |
| Gandu S. et al | 2021 | USA | 7 | 36.20 ±12.70 | 37.4 ±11.5 | 12.49±12.41 | 30 | 22 | R+L | HEIDELBERG |
| Hanifi K. et al | 2022 | Turkey | 6 | 34.74 ±10.68 | 34.02 ±10.3 | Not reported | 47 | 50 | R | OPTOVUE |
| Hosak et al. | 2020 | Czech Republic | 6 | 30.50±7.10 | 33.4±9.1 | 9.7±7.1 | 39 | 32 | R+L | ZEISS |
| Jerotic S. et al | 2020 | Serbia | 6 | 33.10 ±6.10 | 32.5 ±9.9 | 5.9 ±3.9 | 33 | 35 | R+L | ZEISS |
| Jerotic S. et al | 2021 | Serbia | 8 | 33.36 ±6.81 | 30.9 ±9.25 | 6.06 ±2.76 | 42 | 39 | R+L | ZEISS |
| Kango A. et al | 2022 | India | 8 | 29.85±7.01a /  31.17±7.73b /  30.33 ±6.47c | 31 ±3.68 | 8.67±6.31^a^ /  6.22±4.68^b^ /  1.4 ±1.78^c^ | 35^a^ / 35^b^ / 21^c^ | 36 | R+L | TOPCON |
| Kaya H. et al | 2022 | Turkey | 9 | 31.54 ±8.09 | 31.93 ±7.97 | 11.97 ±6.89 | 46 | 46 | R+L | OPTOPOL |
| Khalil DH. et al | 2022 | Turkey | 7 | 30.16 ± 5.60 | 29.73 ±5.18 | 7.34 ±2.1 | 30 | 30 | R+L | OPTOVUE |
| Koman-Wierdak E. et al | 2021 | Poland | 7 | 26.33 ±5.26 | 26.8 ±Not reported | 3.55 ±3.50 | 12 | 15 | RL | Not reported |
| Kurt A. et al | 2021 | Turkey | 6 | 47.82 ±9.44 | 45.59 ±9.92 | 24.79 ±15.22 | 44 | 41 | R+L | ZEISS |
| Kurtulmus A. et al | 2020 | Turkey | 7 |  |  |  | 38 | 38 | R | HEIDELBERG |
| Kurtulmus A. et al | 2023 | Turkey | 9 | 42.38 ±7.74 | 38.9 ±11.91 | 18.44 ±9.84 | 50 | 40 | R+L | HEIDELBERG |
| Lai A. et al | 2020 | USA | 5 | 32.49 ±9.03 | 32 ±12.86 | Not reported | 33 | 38 | R+L | ZEISS |
| Lee W. et al | 2013 | Malaysia | 9 | 31.17 (10.67) | 35.97 ±9.1 | Not reported | 30 | 30 | R | ZEISS |
| Liu Y. et al | 2020 | China | 9 | 44.20 ±12.02 | 41.59 ±9.37 | 22.11 ±12.45 | 221 | 149 | R+L | TOPCON |
| Liu Y. et al. | 2021 | China | 8 | 45.09±12.47 | 41.43±9.5 | 21.74±13.51 | 138 | 160 | R+L | TOPCON |
| Miller M. et al | 2020 | USA | 6 | 48.25 ±10.29 | 48.29 ±10.64 | 27.25 ±14.41 | 12 | 12 | R+L | ZEISS/ HEIDELBERG |
| Mota M. et al | 2015 | Portugal | 6 | 32.90 ±11.90 | 33.4 ± 11.2 |  | 20 | 20 | RL | HEIDELBERG |
| Sarkar S. et al | 2021 | India | 8 | 28.60 ±6.30 | 30.25 ±9.22 | 2.52 ±0.87 | 20 | 20 | R | OPTOS |
| Schönfeldt-Lecuona C. et al | 2020 | Germany | 7 | 37.00 ±10.90 | 40.3 ±11.6 | 10.2 ±10.2 | 26 | 23 | RL | HEIDELBERG |
| Silverstein, Steven M. et al | 2017 | USA | 7 | 40.46 ±12.09 | 39.19 ±11.03 | Not reported | 32 | 32 | R+L | Not reported |
| Taşdelen R. et al | 2023 | Turkey | 8 | 29.50 ±not reported | 26 ±26 | 11.77 ±7.38 | 36 | 36 | R+L | OPTOPOL |
| Topcu-Y. et al | 2017 | Turkey | 9 | 34.64 ±9.49 | 32.08 ±12.33 | 10.33 ±Not reported | 59 | 36 |  | HEIDELBERG |
| Yesilkaya U.H. et al | 2023 | Turkey | 7 | 35.90±8.20a /  34.30±8.80b | 37.4±10.1a / 37.4±10.1b | 9.02±9.2a /  11.4±7.4b | 39a /  43b | 40a / 40b | R | OPTOVUE |
| Yılmaz U. et al | 2016 | Turkey | 6 | 39.85 ±10.28 | 38.59 ±9.58 | Not reported | 34 | 30 | RL | ZEISS |

*Table S 2 we found 18 studies investigating bipolar disorder. N = number of patients / controls. NOS = Newcastle-Ottawa-Scale score, DoD = Duration of disease. R+L Left and Right eye reported separately. RL: only pooled values for both eyes reported. Blank spaces = variable was not reported*

| Study Characteristics – Bipolar Disorder | | | | | | | | | | |
| --- | --- | --- | --- | --- | --- | --- | --- | --- | --- | --- |
| **Authors** | **Year** | **Country** | **N Patients** | **N Controls** | **NOS** | **Laterality** | **Age patients in years (Mean ± SD)** | **Age controls in years (Mean ± SD)** | **Machine** | **DoD in years (Mean ± SD)** |
| Alici S. et al | 2019 | Turkey | 80 | 80 | 9 | RL | 37.8 ±10.3 | 36.9 ±8.9 | OPTOVUE | 13.18 ±9.17 |
| Altun IK.et al. | 2020 | Trukey | 41 | 31 | 8 | R+L | 42.17 ±13.39 | 39.35 ±4.27 | OPTOVUE | 14.3 ±1.22 |
| Ayık B. et al | 2022 | Turkey | 31 | 31 | 8 | R+L | 36 ±7.75 | 31.5 ±7.75 | OPTOPOL | 12 ±Not reported |
| Garcia-Martin E. et al | 2019 | UK | 30 | 80 | 7 | random | 49.67 ±11.20 | 49.97 ±8.75 | HEIDELBERG | 16.47 ±6.29 |
| Gokcinar NB. et al | 2020 | Turkey | 70 | 70 | 9 | random | 40.41 ±13.22 | 40.2 ±13.03 | HEIDELBERG | 14.17 ±10.37 |
| Kalenderoglu A. et al | 2016 | Turkey | 43 | 43 | 8 | RL | 35.55 ±10.49 | 40.46 ±15.45 | HEIDELBERG | Not reported |
| Khalil MA. et al | 2017 | Egypt | 40 | 40 | 9 | R+L | 30.90 ±9.31 | 32.85 ±8.77 | OPTOVUE | 7.01 ±0.82 |
| Kilicarslan T. et al | 2022 | Turkey | 50 | 50 | 6 | R+L | 39.12 ±10.19 | 38.18 ±11.02 | HEIDELBERG | 13.06 ±8.25 |
| Koman-Wierdak E. et al | 2021 | Poland | 8 | 15 | 7 | RL | 24.13 ±8.85 | 26.8 ±Not reported | Not reported | 5.89 ±9.67 |
| Kurt A. et al | 2023 | Turkey | 67 | 51 | 5 | R+L | 39.78 ±11.77 | 42.06 ±12.1 | Not reported | 16.6 ±9.16 |
| Liu Y. et al | 2021 | China | 82 | 274 | 8 | R+L | 38.96 ±14.61 | 41.59 ±9.37 | TOPCON | 13.27 ±11.44 |
| Mehraban A. et al | 2016 | Iran | 30 | 30 | 8 | RL | 33.8  ±9.2 | 31.2  ±9.5 | TOPCON | 10.6 ±8.6 |
| Mustafa A. et al | 2022 | Turkey | 48 | 45 | 7 | R | 37.36 ±10.39 | 32.71 ±10.59 | NIDEK | Not reported |
| Özgedik Turhan N. et al | 2024 | Turkey | 42 | 24 | 6 |  | 30.95 ±9.97 | 30.63 ±5.43 | HEIDELBERG | 4.05 ±48.66 |
| Polo V. et al | 2019 | Spain | 23 | 23 | 8 | random | 49.67 ±8.75 | 49.03 ±9.44 | TOPCON | 16.12 ±6.66 |
| Sánchez-Morla EM. et al | 2021 | Spain | 17 | 42 | 8 | random | 51.47 ±11.94 | 49.74 ±17.01 | HEIDELBERG | 20.64 ±6.48 |
| Satue M. et al | 2022 | Spain | 38 | 122 | 8 | random | 48.33 ±11.92 | 47.65 ±16.11 | HEIDELBERG | 20 ±5.55 |
| Torun IM. et al | 2023 | Turkey | 39 | 36 | 7 | R | 36.15 ±9.00 | 33.33 ±7.8 | HEIDELBERG | Not reported |

*Table S 3 we found 10 studies investigating major depressive disorder. N = number of patients / controls. NOS = Newcastle-Ottawa-Scale score, DoD = Duration of disease. R+L Left and Right eye reported separately. RL: only pooled values for both eyes reported. Blank spaces = variable was not reported*

| Study Characteristics – Major Depressive Disorder | | | | | | | | | | |
| --- | --- | --- | --- | --- | --- | --- | --- | --- | --- | --- |
| **Authors** | **Year** | **Country** | **N Patients** | **N Controls** | **NOS** | **Laterality** | **Age patients in years (Mean ± SD)** | **Age controls in years (Mean ± SD)** | **Machine** | **DoD in years (Mean ± SD)** |
| Genc A. et al | 2019 | Turkey | 24 | 24 | 5 | R+L | 44.00 ±9.59 | 44.37 ±9.28 | OPTOVUE | 14.95 ±10.53 |
| Jung KI. et al | 2020 | South Korea | 49 | 50 | 8 | RL | 48.10 ±15.70 | 45.5 ±13.2 | ZEISS | Not reported |
| Kalenderoglu A. et al | 2016 | Turkey | 50 | 50 | 6 | RL | 40.76 ±9.43 | 41.02 ±13.96 | HEIDELBERG | Not reported |
| Liu Y. et al | 2021 | China | 35 | 274 | 7 | R+L | 43.60 ±15.51 | 41.59 ±9.37 | TOPCON | 8.43  ±9.55 |
| Liu X. et al | 2022 | China | 78 | 47 | 6 | RL | 23.08 ±5.72 | 24.34 ±6.67 | Not reported | 1.02 ±0.62 |
| Lubinski. et al | 2023 | Poland | 29 | 29 | 5 | Not reported | 47.30 (Not reported) | 46.8 (Not reported) | ZEISS | Not reported |
| Schönfeldt-Lecuona C. et al | 2017 | Germany | 28 | 20 | 8 | RL | 46.90 ±9.17 | 43.05 ±9.64 | HEIDELBERG | 5.31 ±4.01 |
| Sönmez İ. et al | 2017 | Turkey | 30 | 30 | 8 | R+L | 34.57 ±6.19 | 35.47 ±8.28 | HEIDELBERG | 5.7 ±7.31 |
| Xiao Q. et al | 2024 | China | 29 | 29 | 8 | RL | 26.70 ±7.91 | 26.8 ±8.13 | Not reported | 2.1 ±1.7 |
| Yıldız M. et al | 2016 | Turkey | 58 | 57 | 7 | R | 44.59 ±13.10 | 40.6 ±10.66 | ZEISS | Not reported |

*Table S 4 we found 3 studies investigating attention deficit/hyperactivity disorder. N = number of patients / controls. NOS = Newcastle-Ottawa-Scale score, DoD = Duration of disease. R+L Left and Right eye reported separately. RL: only pooled values for both eyes reported. Blank spaces = variable was not reported*

| Study Characteristics - Attention Deficit/ Hyperactivity Disorder | | | | | | | | | | |
| --- | --- | --- | --- | --- | --- | --- | --- | --- | --- | --- |
| **Authors** | **Year** | **Country** | **N Patients** | **N Controls** | **NOS** | **Laterality** | **Age patients in years (Mean ± SD)** | **Age controls in years (Mean ± SD)** | **Machine** | **DoD in years (Mean ± SD)** |
| Erdogan E. et al | 2021 | Turkey | 33 | 31 | 7 | R+L | 26.52 ±8.02 | 26.45 ±7.11 | ZEISS |  |
| Kaymak D. et al | 2021 | Turkey | 38 | 30 | 8 | R+L | 23.21 ±8.98 | 23.63 ±3.85 | HEIDELBERG |  |
| Tünel M. et al | 2021 | Turkey | 26 | 26 | 7 | RL | 33.58  ±Not reported | 32.69  ±Not reported | OPTOVUE |  |

*Table S 5 we found 3 studies investigating obsessive compulsive disorder. N = number of patients / controls. NOS = Newcastle-Ottawa-Scale score, DoD = Duration of disease. R+L Left and Right eye reported separately. RL: only pooled values for both eyes reported. Blank spaces = variable was not reported*

| Study Characteristics – Obsessive Compulsive Disorder | | | | | | | | | | |
| --- | --- | --- | --- | --- | --- | --- | --- | --- | --- | --- |
| **Authors** | **Year** | **Country** | **NOS** | **N Patients** | **N Controls** | **Laterality** | **Age patients (Mean** ± **SD)** | **Age controls (Mean** ± **SD)** | **Machine** | **DoD in years (Mean** ± **SD)** |
| Onur O.S. et al | 2020 | Turkey | 8 | 42 | 50 | RL | 35.10 ±10.40 | 35.6 ±8.7 | OPTOVUE | 11 ± 8.4 |
| Özen M.E. et al | 2019 | Turkey | 6 | 50 | 50 | R+L | 33.66 ± 9.85 | 37.96 ± 15.88 | HEIDELBERG | Not reported |
| Polat S. et al. | 2019 | Turkey | 8 | 30 | 31 |  | 28.20±9.90 | 29.5 ± 10.1 | ZEISS | Not reported |

*Table S 6 we found 11 studies investigating substance use disorders. Özsoy et al. 2023, Orum et. al. 2021 and Sahin et. al. 2021 included as opiate use disorder subgroup. Yanhong et al. 2021, Özsoy et al 2020 and Orum et al. 2020 included as alcohol use disorder in the final analysis. N = number of patients / controls. Age in years. NOS = Newcastle-Ottawa-Scale score, DoD = Duration of disease. R+L Left and Right eye reported separately. RL: only pooled values for both eyes reported. Blank spaces = variable was not reported*

| Study Characteristics - Substance Use Disorder | | | | | | | | | | |
| --- | --- | --- | --- | --- | --- | --- | --- | --- | --- | --- |
| **Authors** | **Year** | **Country** | **N Pat-ients** | **N Con-trols** | **NOS** | **Laterality** | **Age patients (Mean ± SD)** | **Age controls  (Mean ± SD)** | **DoD in years (Mean ± SD)** | **Machine** |
| Gemelli H. et al | 2019 | Brazil | 17 | 18 | 7 | R | 62.5 ±13.8 | 52.6 ±11.9 | Not reported | ZEISS |
| Kalenderoglu A. et al | 2020 | Turkey | 111 | 45 | 7 | R+L | 23.39  ±5.35 | 28.48 ±5.21 | Not reported | HEIDELBERG |
| Kaya S. et al | 2023 | Turkey | 27 | 30 | 7 | R+L | 28 ±6.4 | 22.5 ±10.6 | Not reported | Huvitz1_F |
| Kulu M. et al | 2021 | Turkey | 30 | 30 | 8 | R+L | 24.17 ±4.64 | 28.07 ±10.61 | Not reported | ZEISS |
| Yanhong Liu. et al | 2021 | China | 26 | 53 | 9 | R+L | 45.46 ±9.89 | 41.49 ±9.39 | 12.9 ±9.08 | TOPCON |
| Özsoy F. et al | 2020 | Turkey | 43 | 43 | 8 |  | 40.86 ±12.88 | 39.53 ±11.58 | Not reported | ZEISS |
| Özsoy F. et al | 2023 | Turkey | 30 | 30 | 9 | R+L | 26.57 ±5.9 | 28.3 ±10.64 | Not reported | Not specified |
| Orum MH. et al | 2020 | Turkey | 38 | 38 | 8 | R+L | 39.50 ±6.17 | 38.76 ±9.12 | 11 ±8.4 | HEIDELBERG |
| Orum MH. et al | 2021 | Turkey | 43 | 43 | 8 | R+L | 24.48 ±2.84 | 25.32 ±3.34 | Not reported | HEIDELBERG |
| Şahin T. et al | 2021 | Turkey | 29 | 29 | 9 | RL | 42.3 ±7.78 | 37.3 ±12.4 | Not reported | HEIDELBERG |
| Talebnejad MR. et al | 2020 | Iran | 55 | 49 | 7 | RL | 44.63 ±0.69) | 43.08 ±0.91 | Not reported | ZEISS |

**Supplementary Tables 7 – 9 Results of Egger’s Regression Tests**

*Table S 7 Results of Eggers Test for schizophrenia spectrum disorders (SSD). Each meta-regression with one row representing one retinal layer and/or subfield. p > 0.05 as statistically significant Result meaning indication of likely publication bias.*

|  |  | **Results Eggers Test** | | | |
| --- | --- | --- | --- | --- | --- |
| **Diagnosis** | **Layer** | **Intercept** | **95% CI** | **p** | **z-stat.** |
| SSD | pRNFL | -0.3426 | -0.751, 0.066 | 0.995 | 0.0065 |
|  | sRNFL | -0.4546 | -0.991, 0.082 | 0.451 | 0.7530 |
|  | tRNFL | 0.0084 | -0.546, 0.563 | 0.463 | -0.7339 |
|  | iRNFL | -0.4480 | -1.111, 0.215 | 0.5573 | 0.5869 |
|  | nRNFL | 0.3611 | -0.228, 0.951 | 0.0922 | -1.6838 |
|  | **Macula** | **0.0528** | **-0.519, 0.625** | **0.0236** | **-2.2634** |
|  | **cfMacula** | **0.7686** | **0.178, 1.359** | **0.0001** | **-3.8553** |
|  | isMacula | -0.2140 | -1.329, 0.901 | 0.4918 | -0.6875 |
|  | osMacula | -0.1397 | -1.147, 0.868 | 0.3754 | -0.8864 |
|  | itMacula | -0.2340 | -0.969, 0.501 | 0.4723 | -0.7188 |
|  | otMacula | 0.0363 | -0.483, 0.556 | 0.0756 | -1.7770 |
|  | iiMacula | -0.2961 | -0.930, 0.338 | 0.4419 | -0.7689 |
|  | oiMacula | -0.0754 | -0.683, 0.532 | 0.1323 | -1.5050 |
|  | inMacula | 0.0216 | -0.764, 0.7211 | 0.1882 | -1.3161 |
|  | onMacula | -0.1525 | -0.939, 0.634 | 0.3698 | -0.8968 |
|  | Macula Volume | -0.1880 | -0.942, 0.566 | 0.3818 | -0.8745 |
|  | **GCL-IPL** | **-0.9854** | **-1.543, -0.428** | **0.0411** | **2.0430** |

*Table S 8 Results of Eggers Test for bipolar disorder (BPD). Each meta-regression with one row representing one retinal layer and/or subfield. p > 0.05 as statistically significant Result meaning indication of likely publication bias.*

|  |  | **Results Eggers Test** | | | |
| --- | --- | --- | --- | --- | --- |
| **Diagnosis** | **Layer** | **Intercept** | **95% CI** | **p** | **z-stat.** |
| BPD | pRNFL | -0.6712 | -1.323, -0.020 | 0.3981 | 0.8451 |
|  | sRNFL | -0.4753 | -0.974, 0.023 | 0.7188 | 0.3601 |
|  | tRNFL | -0.1949 | -0.970, 0.580 | 0.8346 | 0.2088 |
|  | iRNFL | -0.6518 | -1.149, -0.154 | 0.3125 | 1.0100 |
|  | nRNFL | 0.1904 | -0.689, 1.070 | 0.3366 | -0.9608, |
|  | Macula | 0.2033 | -1.167, 1.574 | 0.5455 | -0.6045 |
|  | cfMacula | 0.0034 | -1.129, 1.135 | 0.3617 | -0.9122 |
|  | isMacula | -1.3659 | -2.811, 0.079 | 0.1954 | 1.2948, |
|  | osMacula | -0.2881 | -1.693, 1.116 | 0.5304 | -0.6274 |
|  | itMacula | -0.7712 | -1.476, -0.066 | 0.3848 | 0.8691 |
|  | **otMacula** | **-1.3367** | **-2.400, -0.273** | **0.0410** | **2.0435** |
|  | **iiMacula** | **-1.4376** | **-2.162, -0.713** | **0.0068** | **2.7079** |
|  | **oiMacula** | **-1.5595** | **-2.274, -0.845** | **0.0003** | **3.5899** |
|  | inMacula | -0.7286 | -1.4687, 0.011 | 0.4138 | 0.8172 |
|  | **onMacula** | **-1.4113** | **-2.127, -0.695** | **0.0030** | **2.9704** |

*Table S 9 Results of Eggers Test for major depressive disorder (MDD), attention deficit/hyperactivity disorder (ADHD), obsessive compulsive disorder (OCD), alcohol use disorder (AUD), and opiate use disorder (OUD). Each meta-regression with one row representing one retinal layer and/or subfield. p > 0.05 as statistically significant Result meaning indication of likely publication bias.*

|  |  | **Results Eggers Test** | | | |
| --- | --- | --- | --- | --- | --- |
| **Diagnosis** | **Layer** | **Intercept** | **95% CI** | **p** | **z-stat.** |
| MDD | pRNFL | -0.5563 | -1.362, 0.249 | 0.2488 | 1.1533 |
|  | sRNFL | -0.7993 | -1.931, 0.333 | 0.1686 | 1.3766 |
|  | tRNFL | -1.0980 | -2.541, 0.345 | 0.1342 | 1.4977 |
|  | iRNFL | -1.5763 | -3.132, -0.021 | 0.0530 | 1.9352 |
|  | nRNFL | -0.1041 | -1.788, 1.580 | 0.9899 | 0.0126 |
|  | Macula | 3.5927 | -8.844, 16.029 | 0.5660 | -0.5739 |
| ADHD | pRNFL | 3.0661 | -2.212, 8.344 | 0.1878 | -1.3172, |
| OCD | pRNFL | -0.6355 | -3.346, 2.075 | 0.7835 | 0.2748 |
| **AUD** | **pRNFL** | **6.6206** | **3.178, 10.063** | **< .0001** | **-3.9917** |
| OUD | pRNFL | 1.3714 | -8.282, 11.025 | 0.7966 | -0.2577 |

**Supplementary Tables 10 – 16 Regression Models**

**Regression Models for Age Group Differences**

*Table S 10 linear regression models between retinal measurements and age group differences (age of the patient group – age of the control group). pRNFL: Peripapillary retinal nerve fiber layer; MV: Macular volume; GCL-IPL: Ganglion cell layer – inner plexiform layer; k: Number of samples included for the analysis; 95% CI: confidence interval; I^2^: residual heterogeneity.*

| **Measure** | **Eye** | **k** | **Coefficient** | **95% CI** | **p** | **I^2^** |
| --- | --- | --- | --- | --- | --- | --- |
| ***Schizophrenia Spectrum Disorders*** | | | | |  |  |
| pRNFL | both | 42 | -0.005 | [-0.049 to 0.039] | 0.83 | 50.21 |
|  | right | 29 | 0.002 | [-0.054 to 0.058] | 0.94 | 57.60 |
|  | left | 20 | 0.012 | [-0.062 to 0.087] | 0.75 | 55.50 |
| iRNFL | both | 26 | -0.022 | [-0.075 to 0.03] | 0.40 | 49.34 |
|  | right | 17 | -0.053 | [-0.127 to 0.022] | 0.17 | 51.77 |
| sRNFL | both | 26 | -0.024 | [-0.077 to 0.029] | 0.37 | 49.42 |
|  | right | 17 | -0.023 | [-0.106 to 0.06] | 0.59 | 60.28 |
| nRNFL | both | 27 | -0.015 | [-0.064 to 0.034] | 0.56 | 45.00 |
|  | right | 18 | -0.011 | [-0.075 to 0.053] | 0.74 | 44.47 |
| tRNFL | both | 26 | 0.038 | [-0.005 to 0.081] | 0.08 | 24.41 |
|  | right | 18 | 0.051 | [-0.004 to 0.105] | 0.07 | 22.62 |
| Macula | both | 16 | 0.078 | [0.015 to 0.141] | **0.015** | 52.23 |
|  | right | 13 | 0.078 | [0.016 to 0.141] | **0.014** | 53.46 |
|  | left | 10 | 0.022 | [-0.038 to 0.081] | 0.48 | 24.73 |
| MV | both | 21 | 0.095 | [0.023 to 0.166] | **0.010** | 20.17 |
|  | right | 14 | 0.078 | [-0.028 to 0.184] | 0.15 | 58.02 |
|  | left | 13 | 0.064 | [-0.034 to 0.162] | 0.20 | 19.86 |
| isMacula | both | 11 | 0.095 | [0.037 to 0.153] | **0.001** | 26.17 |
| osMacula | both | 12 | 0.040 | [-0.075 to 0.155] | 0.50 | 81.87 |
| inMacula | both | 11 | 0.120 | [0.071 to 0.168] | **<0.001** | 0.01 |
| onMacula | both | 12 | 0.138 | [0.089 to 0.188] | **<0.001** | 0.00 |
| iiMacula | both | 11 | 0.105 | [0.057 to 0.154] | **<0.001** | 0.00 |
|  | both | 11 | 0.105 | [0.057 to 0.154] | **<0.001** | 0.00 |
| oiMacula | both | 12 | 0.111 | [0.063 to 0.159] | **<0.001** | 0.00 |
| itMacula | both | 11 | 0.120 | [0.071 to 0.169] | **<0.001** | 0.00 |
| otMacula | both | 11 | 0.098 | [0.049 to 0.147] | **<0.001** | 0.00 |
| cfMacula | both | 22 | 0.078 | [-0.03 to 0.186] | 0.15 | 83.66 |
|  | right | 17 | 0.085 | [-0.073 to 0.243] | 0.29 | 92.35 |
|  | left | 14 | 0.037 | [-0.027 to 0.102] | 0.26 | 37.61 |
| GCL-IPL | both | 10 | -0.048 | [-0.091 to -0.005] | **0.030** | 0.00 |
| ***Bipolar Disorder*** | | |  |  |  |  |
| pRNFL | both | 17 | 0.067 | [-0.05 to 0.184] | 0.26 | 85.52 |
| iRNFL | both | 13 | 0.011 | [-0.051 to 0.073] | 0.74 | 31.01 |
| sRNFL | both | 12 | 0.003 | [-0.06 to 0.065] | 0.93 | 29.58 |
| nRNFL | both | 13 | 0.001 | [-0.085 to 0.087] | 0.99 | 59.48 |
| tRNFL | both | 13 | 0.024 | [-0.05 to 0.097] | 0.53 | 44.34 |
| ***Major Depressive Disorder*** | | | |  |  |  |
| pRNFL | both | 11 | -0.005 | [-0.094 to 0.084] | 0.91 | 43.33 |

**Regression Models for Sex Group Differences**

*Table S 11 linear regression models between retinal measurements and sex group differences (percentage of female patients – percentage of female controls). pRNFL: Peripapillary retinal nerve fiber layer; MV: Macular volume; GCL-IPL: Ganglion cell layer – inner plexiform layer; k: Number of samples included for the analysis; 95% CI: confidence interval; I^2^: residual heterogeneity.*

| **Measure** | **Eye** | **k** | **Coefficient** | **95% CI** | **p** | **I^2^** |
| --- | --- | --- | --- | --- | --- | --- |
| ***Schizophrenia Spectrum Disorders*** | | | | |  |  |
| pRNFL | both | 42 | -0.001 | [-0.013 to 0.01] | 0.81 | 50.01 |
|  | right | 29 | -0.012 | [-0.03 to 0.005] | 0.16 | 52.46 |
|  | left | 20 | -0.019 | [-0.038 to 0.001] | 0.06 | 42.98 |
| iRNFL | both | 26 | 0.012 | [-0.002 to 0.026] | 0.10 | 45.56 |
|  | right | 17 | 0.016 | [-0.007 to 0.039] | 0.18 | 53.25 |
| sRNFL | both | 26 | 0.006 | [-0.009 to 0.021] | 0.41 | 50.74 |
|  | right | 17 | 0.018 | [-0.005 to 0.042] | 0.12 | 55.34 |
| nRNFL | both | 27 | -0.006 | [-0.021 to 0.008] | 0.39 | 43.42 |
|  | right | 18 | 0.013 | [-0.011 to 0.037] | 0.30 | 41.73 |
| tRNFL | both | 26 | 0.007 | [-0.006 to 0.02] | 0.31 | 22.08 |
|  | right | 18 | 0.003 | [-0.019 to 0.026] | 0.78 | 31.03 |
| Macula | both | 16 | 0.017 | [-0.004 to 0.038] | 0.12 | 57.21 |
|  | right | 13 | 0.018 | [-0.003 to 0.039] | 0.09 | 59.14 |
|  | left | 10 | 0.010 | [-0.002 to 0.022] | 0.09 | 0.00 |
| MV | both | 21 | 0.002 | [-0.016 to 0.019] | 0.87 | 39.88 |
|  | right | 14 | -0.001 | [-0.027 to 0.025] | 0.94 | 64.77 |
|  | left | 13 | -0.007 | [-0.023 to 0.01] | 0.44 | 19.80 |
| isMacula | both | 11 | -0.002 | [-0.028 to 0.025] | 0.89 | 61.99 |
| osMacula | both | 12 | 0.006 | [-0.032 to 0.045] | 0.74 | 80.63 |
| inMacula | both | 11 | -0.017 | [-0.047 to 0.013] | 0.26 | 68.41 |
| onMacula | both | 12 | -0.005 | [-0.035 to 0.026] | 0.76 | 70.48 |
| iiMacula | both | 11 | -0.005 | [-0.03 to 0.019] | 0.67 | 56.02 |
|  | both | 11 | -0.005 | [-0.03 to 0.019] | 0.67 | 56.02 |
| oiMacula | both | 12 | -0.014 | [-0.039 to 0.01] | 0.25 | 56.07 |
| itMacula | both | 11 | -0.008 | [-0.036 to 0.02] | 0.59 | 64.89 |
| otMacula | both | 11 | -0.013 | [-0.035 to 0.01] | 0.27 | 47.86 |
| cfMacula | both | 22 | -0.015 | [-0.044 to 0.014] | 0.31 | 85.42 |
|  | right | 17 | -0.017 | [-0.058 to 0.024] | 0.42 | 92.48 |
|  | left | 14 | 0.000 | [-0.016 to 0.016] | 0.97 | 47.22 |
| GCL-IPL | both | 10 | -0.003 | [-0.02 to 0.013] | 0.67 | 33.59 |
| ***Bipolar Disorder*** | | |  |  |  |  |
| pRNFL | both | 17 | 0.008 | [-0.006 to 0.021] | 0.26 | 85.98 |
| iRNFL | both | 13 | -0.002 | [-0.013 to 0.01] | 0.77 | 32.66 |
| sRNFL | both | 12 | -0.003 | [-0.014 to 0.009] | 0.62 | 28.81 |
| nRNFL | both | 13 | -0.010 | [-0.024 to 0.004] | 0.17 | 49.84 |
| tRNFL | both | 13 | 0.008 | [-0.004 to 0.02] | 0.20 | 35.91 |
| ***Major Depressive Disorder*** | | | |  |  |  |
| pRNFL | both | 10 | -0.001 | [-0.064 to 0.062] | 0.97 | 51.01 |

**Regression Models for Newcastle-Ottawa-Scale Score**

*Table S 12 linear regression models between retinal thickness and Newcastle-Ottawa-Scale score. pRNFL: Peripapillary retinal nerve fiber layer; MV: Macular volume; GCL-IPL: Ganglion cell layer – inner plexiform layer; k: Number of samples included for the analysis; 95% CI: confidence interval; I^2^: residual heterogeneity.*

| **Measure** | **Eye** | **k** | **Coefficient** | **95% CI** | **p** | **I^2^** |
| --- | --- | --- | --- | --- | --- | --- |
| ***Schizophrenia Spectrum Disorders*** | | | | |  |  |
| pRNFL | both | 42 | -0.015 | [-0.107 to 0.078] | 0.76 | 49.76 |
|  | right | 29 | -0.058 | [-0.18 to 0.063] | 0.34 | 54.53 |
|  | left | 20 | -0.052 | [-0.201 to 0.098] | 0.50 | 50.62 |
| iRNFL | both | 26 | -0.041 | [-0.159 to 0.076] | 0.49 | 50.86 |
|  | right | 17 | -0.205 | [-0.378 to -0.032] | **0.020** | 45.22 |
| sRNFL | both | 26 | -0.070 | [-0.186 to 0.047] | 0.24 | 49.78 |
|  | right | 17 | -0.177 | [-0.364 to 0.011] | 0.07 | 53.32 |
| nRNFL | both | 27 | 0.041 | [-0.073 to 0.154] | 0.48 | 44.03 |
|  | right | 18 | -0.112 | [-0.287 to 0.062] | 0.21 | 39.42 |
| tRNFL | both | 26 | -0.014 | [-0.114 to 0.086] | 0.78 | 27.93 |
|  | right | 18 | 0.012 | [-0.155 to 0.179] | 0.88 | 31.96 |
| Macula | both | 16 | 0.068 | [-0.088 to 0.225] | 0.39 | 62.86 |
|  | right | 13 | 0.129 | [-0.033 to 0.291] | 0.12 | 60.26 |
|  | left | 10 | 0.093 | [-0.029 to 0.214] | 0.13 | 8.48 |
| MV | both | 21 | -0.067 | [-0.199 to 0.066] | 0.32 | 36.80 |
|  | right | 14 | -0.118 | [-0.326 to 0.09] | 0.27 | 59.80 |
|  | left | 13 | -0.039 | [-0.206 to 0.128] | 0.64 | 18.40 |
| isMacula | both | 11 | 0.081 | [-0.101 to 0.262] | 0.39 | 62.40 |
| osMacula | both | 12 | 0.047 | [-0.216 to 0.31] | 0.73 | 82.58 |
| inMacula | both | 11 | 0.066 | [-0.148 to 0.279] | 0.55 | 73.13 |
| onMacula | both | 12 | 0.063 | [-0.145 to 0.271] | 0.55 | 72.24 |
| iiMacula | both | 11 | 0.097 | [-0.067 to 0.261] | 0.24 | 53.88 |
|  | both | 11 | 0.097 | [-0.067 to 0.261] | 0.24 | 53.88 |
| oiMacula | both | 12 | 0.129 | [-0.028 to 0.287] | 0.11 | 51.61 |
| itMacula | both | 11 | 0.077 | [-0.115 to 0.269] | 0.43 | 66.62 |
| otMacula | both | 11 | 0.026 | [-0.165 to 0.218] | 0.79 | 56.65 |
| cfMacula | both | 22 | 0.039 | [-0.183 to 0.262] | 0.73 | 85.35 |
|  | right | 17 | 0.018 | [-0.355 to 0.391] | 0.92 | 92.85 |
|  | left | 14 | 0.113 | [-0.013 to 0.239] | 0.08 | 28.80 |
| GCL-IPL | both | 10 | -0.156 | [-0.262 to -0.049] | **0.004** | 0.00 |
| ***Bipolar Disorder*** | | |  |  |  |  |
| pRNFL | both | 17 | -0.216 | [-0.45 to 0.018] | 0.07 | 84.20 |
| iRNFL | both | 13 | -0.102 | [-0.307 to 0.104] | 0.33 | 29.69 |
| sRNFL | both | 12 | -0.044 | [-0.253 to 0.165] | 0.68 | 32.69 |
| nRNFL | both | 13 | 0.035 | [-0.204 to 0.274] | 0.77 | 58.65 |
| tRNFL | both | 13 | -0.083 | [-0.283 to 0.117] | 0.41 | 43.29 |
| ***Major Depressive Disorder*** | | | |  |  |  |
| RNFL | both | 11 | -0.095 | [-0.238 to 0.048] | 0.19 | 26.75 |

**Regression Models for Duration of Disease**

*Table S 13 linear regression models between retinal thickness and duration of disease (in years). pRNFL: Peripapillary retinal nerve fiber layer; MV: Macular volume; GCL-IPL: Ganglion cell layer – inner plexiform layer; k: Number of samples included for the analysis; 95% CI: confidence interval; I^2^: residual heterogeneity.*

| **Measure** | **Eye** | **k** | **Coefficient** | **95% CI** | **p** | **I^2^** |
| --- | --- | --- | --- | --- | --- | --- |
| ***Schizophrenia Spectrum Disorders*** | | | | |  |  |
| pRNFL | both | 31 | -0.019 | [-0.037 to -0.001] | **0.042** | 42.56 |
|  | right | 25 | -0.024 | [-0.043 to -0.005] | **0.015** | 42.55 |
|  | left | 18 | -0.018 | [-0.039 to 0.003] | 0.10 | 41.72 |
| iRNFL | both | 19 | -0.033 | [-0.058 to -0.008] | **0.009** | 29.54 |
|  | right | 14 | -0.039 | [-0.07 to -0.009] | **0.012** | 43.25 |
| sRNFL | both | 19 | -0.030 | [-0.057 to -0.003] | **0.032** | 38.34 |
|  | right | 14 | -0.036 | [-0.062 to -0.01] | **0.007** | 27.56 |
| nRNFL | both | 20 | 0.000 | [-0.033 to 0.033] | 0.99 | 51.29 |
|  | right | 15 | -0.019 | [-0.053 to 0.016] | 0.29 | 47.02 |
| tRNFL | both | 19 | 0.011 | [-0.014 to 0.036] | 0.40 | 13.58 |
|  | right | 15 | 0.021 | [-0.006 to 0.047] | 0.13 | 8.75 |
| Macula | both | 10 | 0.020 | [-0.002 to 0.042] | 0.07 | 15.08 |
|  | right | 9 | 0.006 | [-0.022 to 0.033] | 0.69 | 36.36 |
|  | left | 8 | 0.018 | [-0.004 to 0.039] | 0.10 | 1.95 |
| MV | both | 15 | -0.012 | [-0.035 to 0.012] | 0.32 | 31.34 |
|  | right | 10 | -0.040 | [-0.067 to -0.014] | **0.003** | 29.41 |
|  | left | 10 | -0.004 | [-0.033 to 0.025] | 0.79 | 43.68 |
| isMacula | both | 7 | -0.014 | [-0.053 to 0.025] | 0.48 | 48.20 |
| osMacula | both | 7 | -0.030 | [-0.112 to 0.051] | 0.46 | 87.83 |
| inMacula | both | 7 | 0.048 | [-0.004 to 0.099] | 0.07 | 68.56 |
| onMacula | both | 7 | 0.019 | [-0.023 to 0.062] | 0.38 | 56.38 |
| iiMacula | both | 7 | 0.004 | [-0.035 to 0.044] | 0.83 | 50.18 |
|  | both | 7 | 0.004 | [-0.035 to 0.044] | 0.83 | 50.18 |
| oiMacula | both | 7 | 0.022 | [-0.023 to 0.066] | 0.34 | 58.59 |
| itMacula | both | 7 | 0.025 | [-0.019 to 0.069] | 0.27 | 59.00 |
| otMacula | both | 7 | 0.033 | [-0.012 to 0.078] | 0.15 | 58.95 |
| cfMacula | both | 14 | 0.046 | [-0.034 to 0.127] | 0.26 | 93.64 |
|  | right | 12 | 0.060 | [-0.037 to 0.157] | 0.22 | 95.30 |
|  | left | 11 | 0.001 | [-0.03 to 0.032] | 0.95 | 52.85 |
| GCL-IPL | both | 7 | 0.015 | [-0.03 to 0.06] | 0.52 | 26.64 |
| ***Bipolar Disorder*** | | |  |  |  |  |
| pRNFL | both | 14 | 0.026 | [-0.052 to 0.105] | 0.51 | 88.60 |
| iRNFL | both | 11 | 0.044 | [0.009 to 0.078] | **0.012** | 0.68 |
| sRNFL | both | 10 | 0.028 | [-0.011 to 0.068] | 0.16 | 11.13 |
| nRNFL | both | 11 | 0.029 | [-0.012 to 0.07] | 0.16 | 29.27 |
| tRNFL | both | 11 | 0.004 | [-0.041 to 0.049] | 0.86 | 42.83 |
| ***Major Depressive Disorder*** | | | |  |  |  |
| pRNFL | both | 6 | 0.036 | [-0.04 to 0.111] | 0.35 | 67.17 |

**Regression Models for Psychopathology**

*Table S 14 linear regression models between retinal thickness and Psychopathology: Positive and Negative Syndrome Scale for Schizophrenia (PANSS), Young Mania Rating Scale (YMRS) or Hamilton Depression Score (HAMD-D). pRNFL: Peripapillary retinal nerve fiber layer; MV: Macular volume; GCL-IPL: Ganglion cell layer – inner plexiform layer; k: Number of samples included for the analysis; 95% CI: confidence interval; I^2^: between study heterogeneity.*

| **Measure** | **Eye** | **k** | **Coefficient** | **95% CI** | **p** | **I^2^** |
| --- | --- | --- | --- | --- | --- | --- |
| ***Schizophrenia Spectrum Disorders - PANSS*** | | | | |  |  |
| pRNFL | both | 13 | -0.006 | [-0.013 to 0.001] | 0.09 | 28.05 |
|  | right | 5 | -0.007 | [-0.015 to 0.001] | 0.10 | 18.67 |
| iRNFL | both | 9 | -0.005 | [-0.012 to 0.003] | 0.23 | 40.66 |
|  | right | 6 | -0.006 | [-0.015 to 0.004] | 0.25 | 52.29 |
| sRNFL | both | 9 | -0.005 | [-0.011 to 0.002] | 0.14 | 9.48 |
|  | right | 6 | -0.006 | [-0.015 to 0.002] | 0.14 | 30.39 |
| **nRNFL** | both | 10 | -0.007 | [-0.013 to -0.001] | **0.037** | 1.08 |
|  | right | 6 | -0.009 | [-0.017 to -0.001] | **0.021** | 0.00 |
| tRNFL | both | 10 | 0.001 | [-0.007 to 0.009] | 0.81 | 48.83 |
|  | right | 6 | 0.002 | [-0.009 to 0.014] | 0.66 | 57.70 |
| **Macula** | both | 5 | -0.014 | [-0.027 to -0.001] | **0.041** | 24.34 |
| MV | both | 6 | -0.001 | [-0.013 to 0.011] | 0.87 | 6.89 |
| **cfMacula** | both | 8 | -0.008 | [-0.016 to -0.001] | **0.034** | 0.00 |
| ***Bipolar Disorder - YMRS*** | | |  |  |  |  |
| pRNFL | both | 8 | 0.007 | [-0.037 to 0.05] | 0.77 | 56.94 |
| ***Major Depressive Disorder - HAM-D*** | | | |  |  |  |
| pRNFL | both | 6 | -0.002 | [-0.07 to 0.065] | 0.95 | 95.43 |

**Regression Models for Chlorpromazine Equivalents**

*Table S 15 linear regression models between retinal thickness and chlorpromazine equivalents (in units of 100 mg). pRNFL: Peripapillary retinal nerve fiber layer; MV: Macular volume; GCL-IPL: Ganglion cell layer – inner plexiform layer; k: Number of samples included for the analysis; 95% CI: confidence interval; I^2^: between study heterogeneity.*

| **Measure** | **Eye** | **k** | **Coefficient** | **95% CI** | **p** | **I^2^** |
| --- | --- | --- | --- | --- | --- | --- |
| ***Schizophrenia Spectrum Disorders*** | | | | |  |  |
| **pRNFL** | both | 12 | 0.086 | [0.012 to 0.16] | **0.02** | 33.72 |
| iRNFL | both | 6 | -0.041 | [-0.145 to 0.063] | 0.44 | 0.01 |
| sRNFL | both | 6 | -0.055 | [-0.22 to 0.109] | 0.51 | 59.54 |
| nRNFL | both | 8 | -0.045 | [-0.128 to 0.038] | 0.29 | 0.00 |
| tRNFL | both | 8 | 0.017 | [-0.105 to 0.138] | 0.79 | 52.34 |
| Macula | both | 8 | -0.042 | [-0.103 to 0.019] | 0.18 | 0.96 |
| MV | both | 8 | 0.027 | [-0.076 to 0.131] | 0.606 | 0.00 |
| cfMacula | both | 11 | -0.010 | [-0.091 to 0.071] | 0.804 | 27.25 |

**Regression Models for Smoking Group Differences**

*Table S 16 linear regression models between retinal thickness and smoking group differences (percentage of smokers among patients – percentage of smokers among controls). pRNFL: Peripapillary retinal nerve fiber layer; MV: Macular volume; GCL-IPL: Ganglion cell layer – inner plexiform layer; k: Number of samples included for the analysis; 95% CI: confidence interval; I^2^: between study heterogeneity.*

| **Measure** | **Eye** | **k** | **Coefficient** | **95% CI** | **p** | **I^2^** |
| --- | --- | --- | --- | --- | --- | --- |
| ***Schizophrenia Spectrum Disorders*** | | | | |  |  |
| pRNFL | both | 10 | -0.006 | [-0.018 to 0.005] | 0.29 | 47.81 |
|  | right | 5 | -0.002 | [-0.018 to 0.014] | 0.78 | 45.06 |
| iRNFL | both | 5 | -0.010 | [-0.029 to 0.009] | 0.31 | 50.27 |
| sRNFL | both | 5 | -0.003 | [-0.019 to 0.012] | 0.66 | 11.64 |
| nRNFL | both | 5 | -0.006 | [-0.017 to 0.006] | 0.34 | 19.11 |
| Macula | both | 5 | -0.009 | [-0.021 to 0.004] | 0.17 | 0.00 |
|  | right | 5 | -0.009 | [-0.021 to 0.004] | 0.16 | 0.00 |
| cfMacula | both | 5 | -0.007 | [-0.021 to 0.007] | 0.35 | 43.77 |
| GCL-IPL | both | 5 | 0.011 | [-0.008 to 0.03] | 0.26 | 37.25 |
| ***Bipolar Disorder*** | | |  |  |  |  |
| pRNFL | both | 12 | -0.010 | [-0.024 to 0.004] | 0.15 | 55.45 |
| iRNFL | both | 9 | -0.011 | [-0.023 to 0.002] | 0.09 | 24.58 |
| sRNFL | both | 8 | -0.005 | [-0.017 to 0.008] | 0.47 | 28.54 |
| nRNFL | both | 10 | -0.007 | [-0.021 to 0.007] | 0.31 | 44.35 |
| tRNFL | both | 10 | -0.001 | [-0.013 to 0.011] | 0.88 | 29.40 |

**Supplementary Table 17 Grading of the Evidence**

*Table S 17 detailed GRADE assessment for the meta-analyses that yielded statistically significant results. Major depressive disorder was not assessed, as no outcomes reached statistical significance. pRNFL: Peripapillary retinal nerve fiber layer; MV: Macular volume; GCL-IPL: Ganglion cell layer – inner plexiform layer; k: Number of samples included for the analysis.*

| **Measure** | **Eye** | **k** | **Magnitude** | **Dose Response** | **Risk of Bias** | **Precision** | **Consistency** | **Publication Bias** | **GRADE** |
| --- | --- | --- | --- | --- | --- | --- | --- | --- | --- |
| ***Schizophrenia Spectrum Disorder*** | | | | | | | | | |
| pRNFL | both | 42 | = | **↑**^b^ | = | = | = | = | MODERATE |
|  | right | 29 | = | **↑**^b^ | = | **↓**^c^ | = | = | LOW |
|  | left | 20 | = | = | = | **↓**^c^ | = | = | VERY LOW |
| iRNFL | both | 26 | = | **↑**^b^ | = | **↓**^c^ | = | = | LOW |
|  | right | 17 | **↓**^a^ | **↑**^b^ | = | **↓**^c^ | = | = | VERY LOW |
|  | left | 8 | = | = | = | **↓**^c^ | = | **↓**^g^ | VERY LOW |
| sRNFL | both | 26 | = | **↑**^b^ | = | **↓**^c^ | = | = | LOW |
|  | right | 17 | = | **↑**^b^ | = | **↓**^c^ | = | = | LOW |
|  | left | 8 | = | = | = | **↓**^c^ | = | **↓**^g^ | VERY LOW |
| nRNFL | both | 27 | **↓**^a^ | = | = | **↓**^c^ | = | = | VERY LOW |
| tRNFL | both | 26 | **↓**^a^ | = | = | **↓**^c^ | = | = | VERY LOW |
|  | right | 18 | **↓**^a^ | = | = | **↓**^c^ | = | = | VERY LOW |
| Macula | both | 16 | = | = | = | = | = | = | LOW |
|  | right | 13 | = | = | = | = | = | = | LOW |
|  | left | 10 | = | = | = | = | = | = | LOW |
| MV | both | 21 | = | = | = | = | = | = | LOW |
|  | right | 14 | = | **↑**^b^ | = | = | = | = | MODERATE |
|  | left | 13 | = | = | = | = | = | = | LOW |
| isMacula | both | 11 | = | = | = | = | = | = | LOW |
|  | right | 8 | = | = | = | = | **↓**^f^ | **↓**^g^ | VERY LOW |
|  | left | 5 | = | = | = | **↓**^c^ | **↓**^f^ | **↓**^g^ | VERY LOW |
| osMacula | both | 12 | = | = | = | = | = | = | LOW |
|  | right | 9 | = | = | = | = | **↓**^f^ | **↓**^g^ | VERY LOW |
| inMacula | both | 11 | = | = | = | = | = | = | LOW |
|  | right | 8 | = | = | = | = | **↓**^f^ | **↓**^g^ | VERY LOW |
| onMacula | both | 12 | = | = | = | = | = | = | LOW |
|  | right | 9 | = | = | = | **↓**^c^ | **↓**^f^ | **↓**^g^ | VERY LOW |
|  | left | 6 | = | = | = | **↓**^c^ | = | **↓**^g^ | VERY LOW |
| iiMacula | both | 11 | = | = | = | = | = | = | LOW |
|  | right | 8 | = | = | = | = | **↓**^f^ | **↓**^g^ | VERY LOW |
|  | left | 5 | = | = | = | **↓**^c^ | **↓**^f^ | **↓**^g^ | VERY LOW |
| oiMacula | both | 12 | = | = | = | = | = | = | LOW |
|  | right | 9 | = | = | = | = | **↓**^f^ | **↓**^g^ | VERY LOW |
|  | left | 6 | = | = | = | **↓**^c^ | = | **↓**^g^ | VERY LOW |
| itMacula | both | 11 | = | = | = | = | = | = | LOW |
|  | right | 8 | = | = | = | **↓**^c^ | **↓**^f^ | **↓**^g^ | VERY LOW |
| otMacula | both | 11 | = | = | = | = | = | = | LOW |
|  | right | 8 | = | = | = | **↓**^c^ | **↓**^f^ | **↓**^g^ | VERY LOW |
|  | left | 6 | **↓**^a^ | = | = | **↓**^c^ | = | **↓**^g^ | VERY LOW |
| cfMacula | both | 22 | = | = | = | **↓**^c^ | = | = | VERY LOW |
|  | right | 17 | = | = | = | **↓**^c^ | = | = | VERY LOW |
|  | left | 14 | = | = | = | **↓**^c^ | = | = | VERY LOW |
| IPL | both | 3 | = | = | = | **↓**^d^ | = | **↓**^g^ | VERY LOW |
|  | right | 3 | = | = | = | **↓**^e^ | = | **↓**^g^ | VERY LOW |
| GCL-IPL | both | 10 | = | = | = ^h^ | = | = | = | LOW |
|  | right | 9 | = | = | = | = | = | **↓**^g^ | VERY LOW |
|  | left | 9 | = | = | = | = | = | **↓**^g^ | VERY LOW |
| GCC | both | 8 | = | = | = | **↓**^c^ | **↓**^f^ | **↓**^g^ | VERY LOW |
|  | right | 6 | = | = | = | **↓**^c^ | **↓**^f^ | **↓**^g^ | VERY LOW |
|  | left | 4 | = | = | = | **↓**^e^ | = | **↓**^g^ | VERY LOW |
| ***Bipolar Disorder*** | | | | | | | | | |
| pRNFL | both | 17 | = | = | = | = | = | = | LOW |
|  | left | 6 | = | = | = | **↓**^c^ | **↓**^f^ | **↓**^g^ | VERY LOW |
| iRNFL | both | 13 | = | = | = | = | = | = | LOW |
|  | right | 4 | = | = | = | = | = | **↓**^g^ | VERY LOW |
|  | left | 3 | = | = | = | = | = | **↓**^g^ | VERY LOW |
| sRNFL | both | 12 | = | = | = | = | = | = | LOW |
|  | right | 4 | = | = | = | = | = | **↓**^g^ | VERY LOW |
|  | left | 3 | = | = | = | = | = | **↓**^g^ | VERY LOW |
| nRNFL | both | 13 | **↓**^a^ | = | = | **↓**^c^ | = | = | VERY LOW |
| isMacula | both | 3 | = | = | = | **↓**^c^ | **↓**^f^ | **↓**^g^ | VERY LOW |
| osMacula | both | 3 | = | = | = | = | = | **↓**^g^ | VERY LOW |
| inMacula | both | 3 | = | = | = | = | = | **↓**^g^ | VERY LOW |
| itMacula | both | 3 | = | = | = | = | = | **↓**^g^ | VERY LOW |
| cfMacula | both | 6 | = | = | = | **↓**^c^ | **↓**^f^ | **↓**^g^ | VERY LOW |
|  | right | 4 | = | = | = | **↓**^c^ | **↓**^f^ | **↓**^g^ | VERY LOW |
| ***Obsessive Compulsive Disorder*** | | | | | | | | | |
| pRNFL | both | 3 | = | = | = | **↓**^d^ | = | **↓**^g^ | VERY LOW |
| ***Attention Deficit and Hyperactivity Disorder*** | | | | | | | | | |
| pRNFL | both | 3 | = | = | = | **↓**^d^ | **↓**^f^ | **↓**^g^ | VERY LOW |
| ***Alcohol Use Disorder*** | | | | | | | | | |
| pRNFL | both | 3 | = | = | = | **↓**^d^ | = | **↓**^g^ | VERY LOW |
| ***Opiate Use Disorder*** | | | | | | | | | |
| pRNFL | both | 3 | = | = | = | **↓**^d^ | = | **↓**^g^ | VERY LOW |

a: downgrade due to small magnitude of the effect (SMD < 0.2);

b: upgrade due to presence of a significant negative meta-regression with duration of disease;

c: downgrade due to imprecision of the confidence intervals (UCI > -0.20);

d: downgrade due to imprecision of the confidence intervals (UCI > -0.20) with sample size smaller than 50% of the optimal information size (N < 400);

e: downgrade due to imprecision arising from sample size smaller than 50% of the optimal information size (N < 400);

f: downgrade due to heterogeneity between studies (I^2^ > 50%) and low number of samples (k < 10);

g: downgrade due low number of samples (k < 10);

h: downgrade due to a significant negative correlation with age group differences was not applied, as the controlled meta-analysis yielded consistent results.

**Supplementary Figures 2 –21**

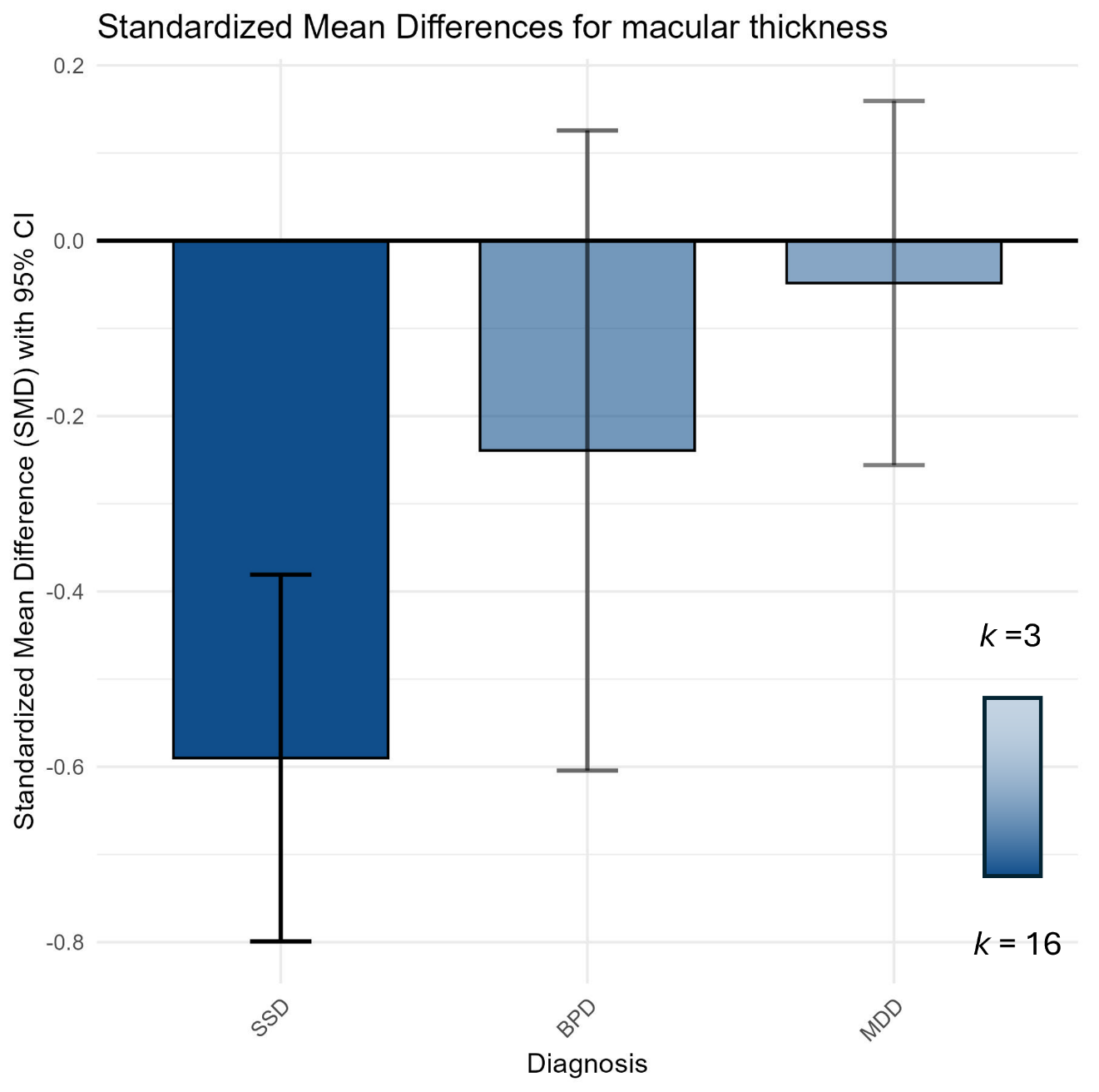

*Figure S 2 Random effects model results of the meta-analysis for macular thickness for Schizophrenia Spectrum disorders (SSD), bipolar disorder (BPD), and Major Depression (MDD). Each bar plot shows the result of a meta-analysis of multiple studies with the estimated effect size as standardized mean differences and corresponding confidence intervals on the y-axis. Color transparency of bar plots and confidence intervals corresponds to number of studies available for final analysis with higher color saturation meaning higher number of studies: SSD: 16 studies, BD 6 Studies, MDD 3 studies.*

**Peripapillary RNFL**

**Schizophrenia Spectrum Disorders**

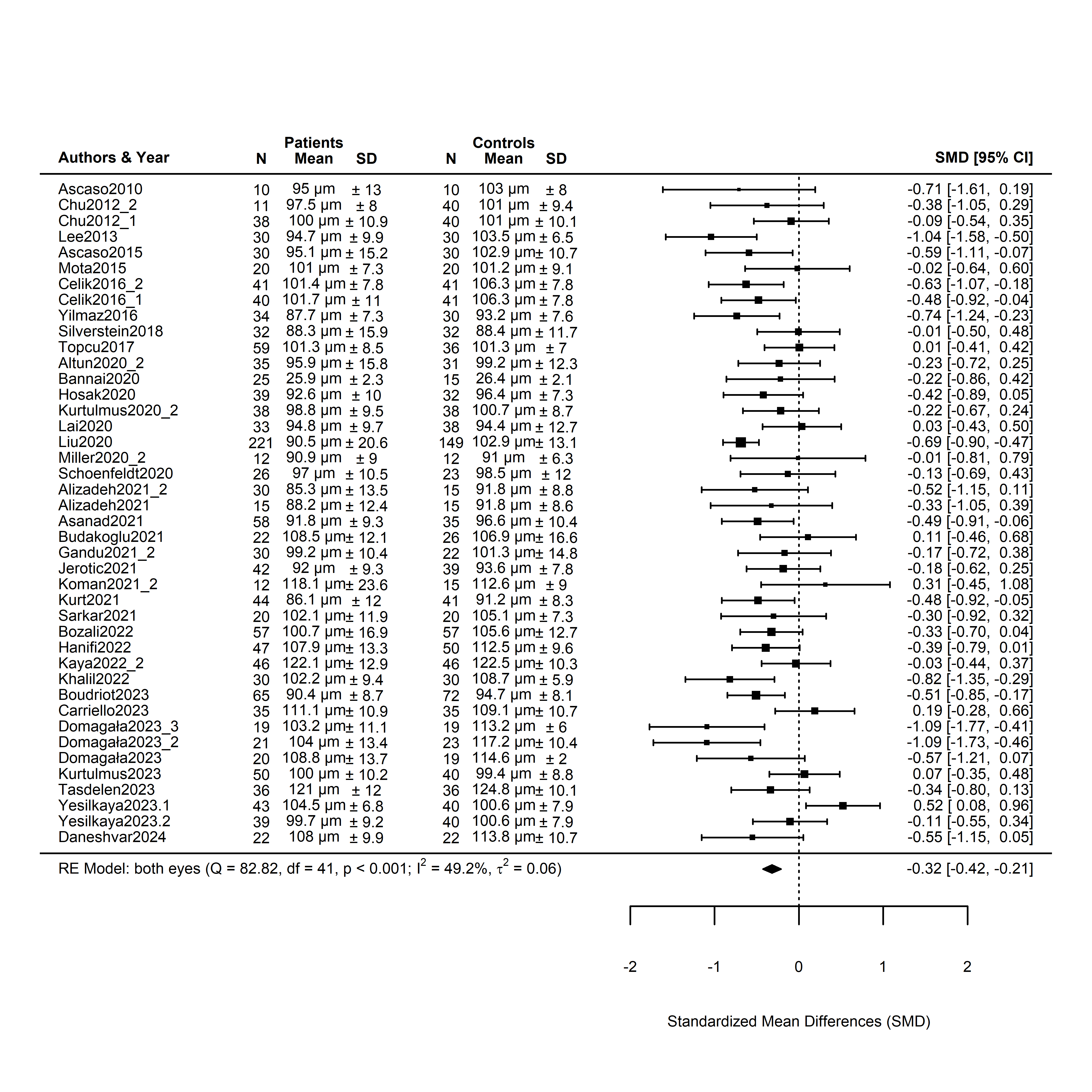

*Figure S 3 Forest plot of a meta-analysis comparing pRNFL thickness in patients with schizophrenia spectrum disorders (SSD) to controls. This forest plot illustrates the standardized mean differences (SMD) in pRNFL thickness between SSD patients and control subjects across multiple studies. Each horizontal line represents an individual study, with the center of the box indicating the SMD and the horizontal line representing the 95% confidence interval (CI). Studies are identified by the first author's last name and year of publication. 42 individual samples from a total of 36 clinical studies were included. 5 studies compared independently sampled subgroups: Chu2012 compared 39 patients with schizophrenia (Chu2012.2) and 11 patients with schizoaffective disorder (Chu2012.1) to 40 controls. Celik20216 compared 41 patients with treatment resistant schizophrenia (Celik2016.1) and 40 patients with Schizophrenia (Celik2016.2) to 41 controls. Alizadeh 2021 compared 30 patients with chronic SSD (Alizadeh2021.1) with 15 patients with acute SSD (Alizadeh2021.2). Domagala 2023 compared three different age groups: 20-30 years (Domagala2023.1), 32-45 years (Domagala2023.2), and 45-60 years (Domagala2023.3)*

**Bipolar Disorder**

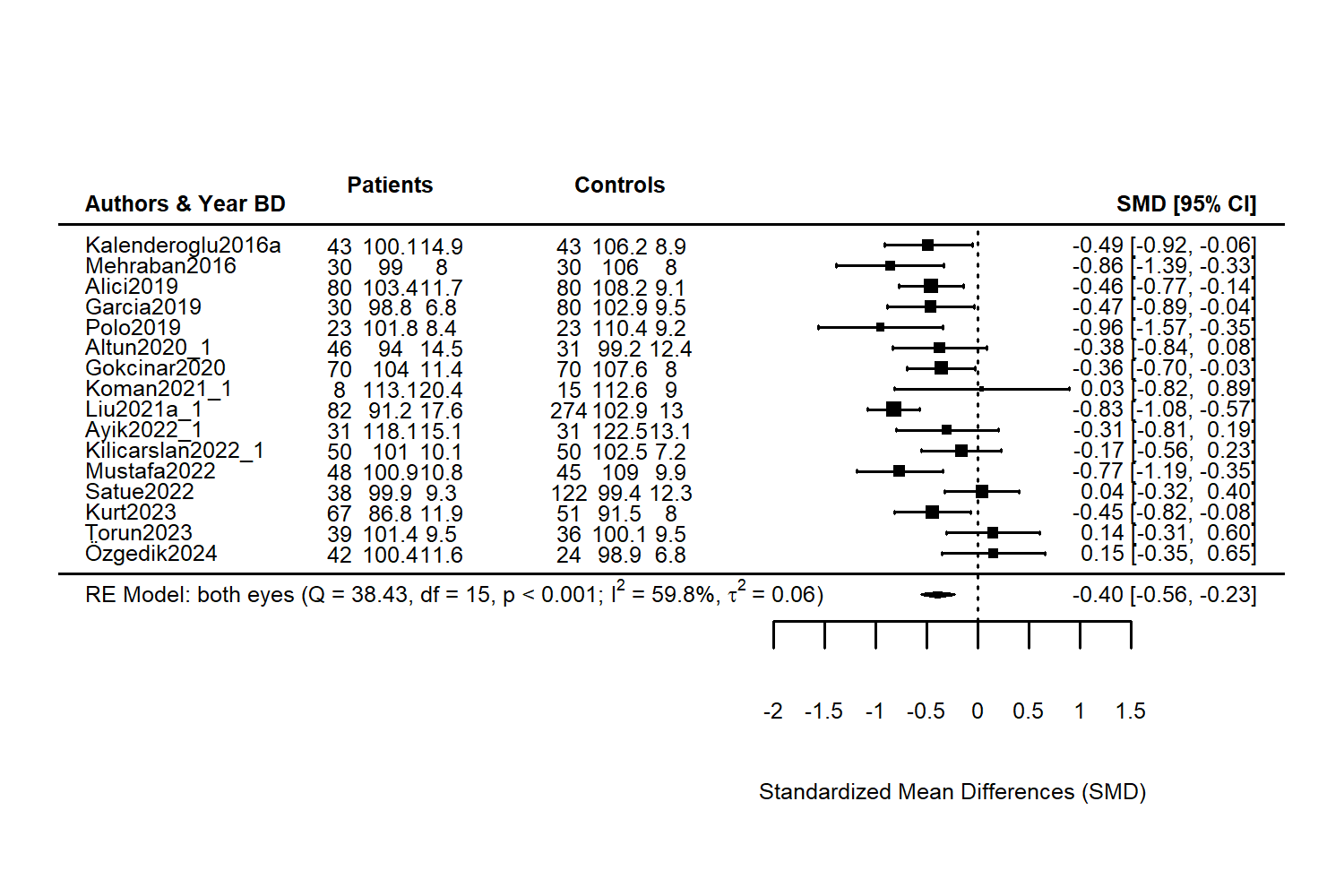

*Figure S 4 Forest plot of a meta-analysis comparing pRNFL thickness in patients with bipolar disorder (BPD) to controls. This forest plot illustrates the standardized mean differences (SMD) in RNFL thickness between BPD patients and control subjects across multiple studies. Each horizontal line represents an individual study, with the center of the box indicating the SMD and the horizontal line representing the 95% confidence interval (CI). Studies are identified by the first author's last name and year of publication.*

**Major Depression**

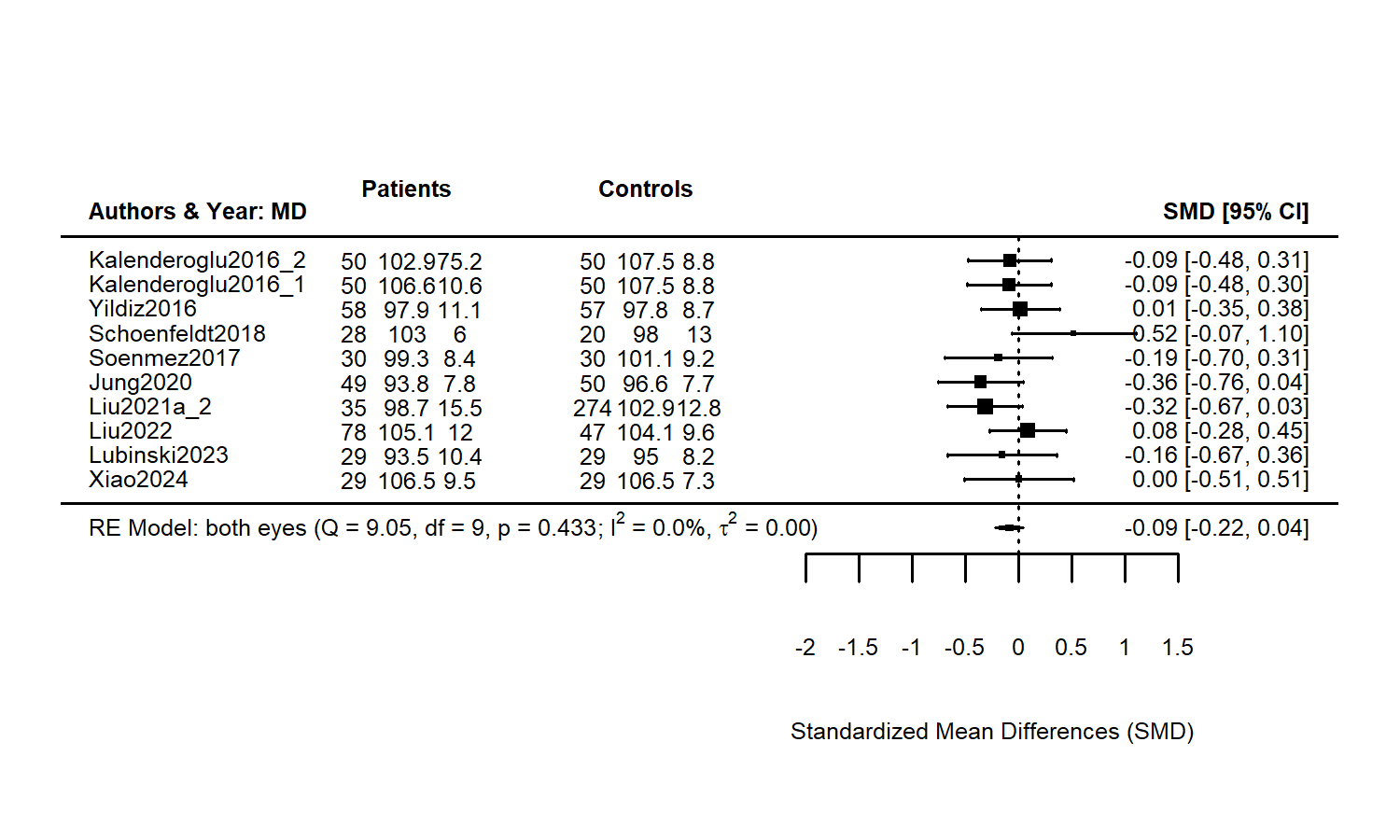

*Figure S 5 Forest plot of a meta-analysis comparing RNFL thickness in patients with major depressive disorder (MDD) to controls across multiple studies. Each horizontal line represents an individual study, with the center of the box indicating the SMD and the horizontal line representing the 95% confidence interval (CI). Bottom line: Results for RE Model with residual heterogeneity (QE), degrees of freedom (df), corresponding p-value (p), I^2 and t^2 statistics. Diamond shape at the bottom shows the model result as SMD. Numbers to the right of the study ID signify the number of participants, mean, and SD of pRNFL thickness per group. For pRNFL thickness in MDD, we included 10 samples from 9 clinical studies, Kalenderoglu2016 compared two independently sampled subgroups: patients with a first episode of major depression (Kalenderoglu2016_1) and patients with recurrent depressive episodes (Kalenderoglu2016_2).*

**Attention Deficit and Hyperactivity Disorder**

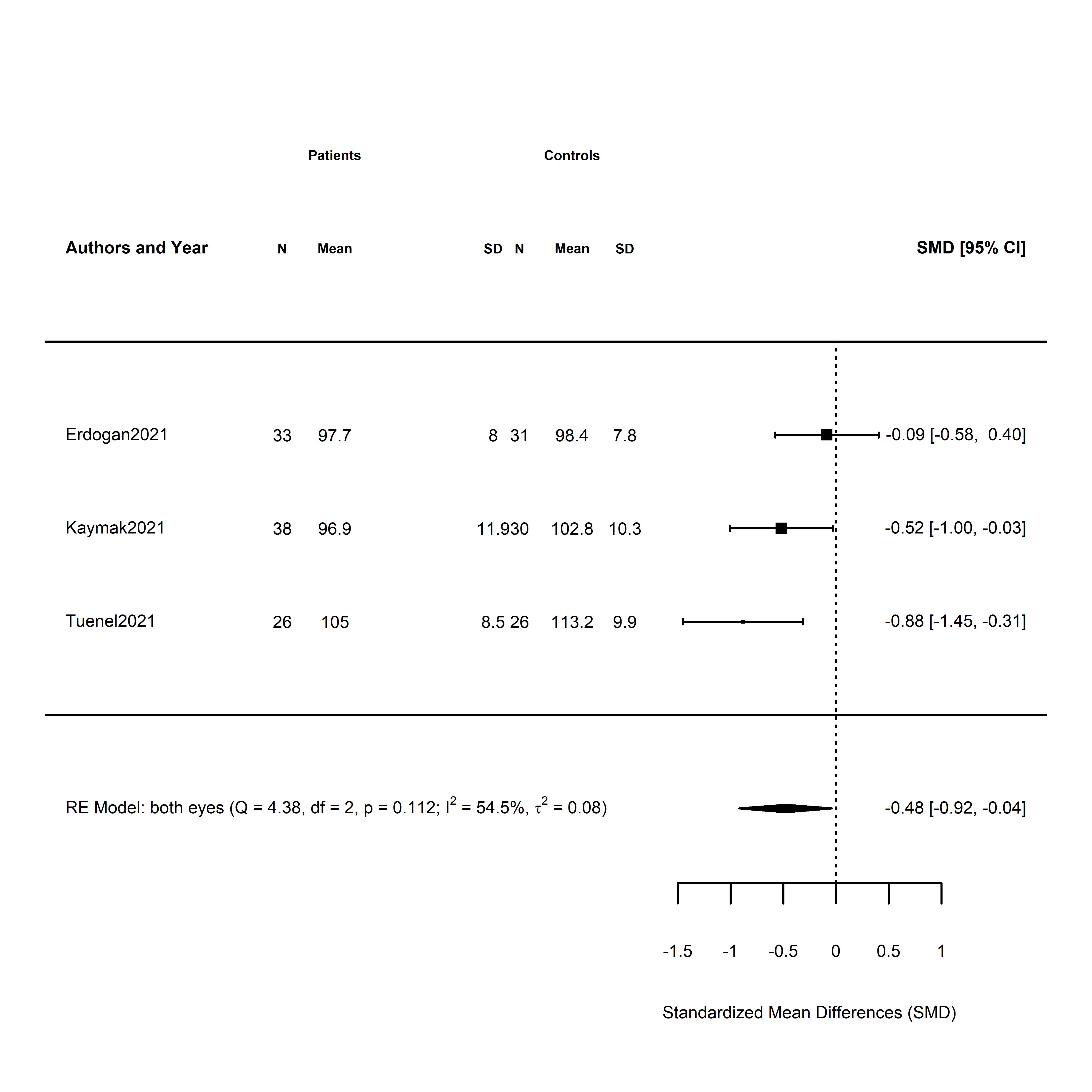

*Figure S 6 Forest plot of a meta-analysis comparing pRNFL thickness in patients with attention deficit/hyperactivity disorder (ADHD) to controls. This forest plot illustrates the standardized mean differences (SMD) in RNFL thickness between ADHD patients and control subjects across multiple studies. Each horizontal line represents an individual study, with the center of the box indicating the SMD and the horizontal line representing the 95% confidence interval (CI). Bottom line: Results for RE Model with residual heterogeneity (QE), degrees of freedom (df), corresponding p-value (p), I^2 and t^2 statistics. Diamond shape at the bottom shows result model result as SMD. Numbers to the right of the study ID signify the number of participants, mean, and SD of pRNFL thickness per group.*

**Alcohol Use Disorder**

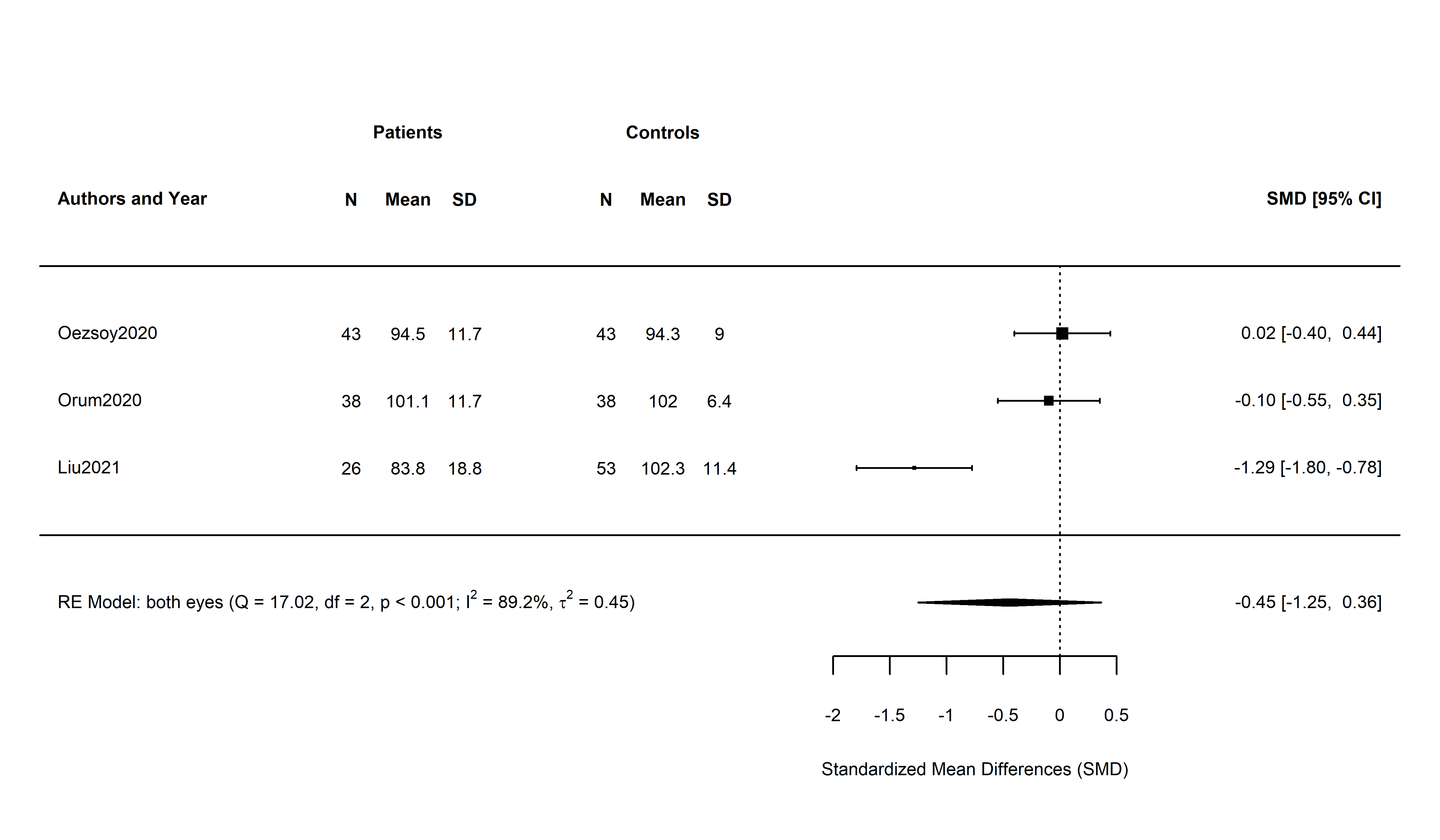

*Figure S 7 Forest plot of a meta-analysis comparing pRNFL thickness in patients with alcohol use disorder (AUD) to controls. This forest plot illustrates the standardized mean differences (SMD) in RNFL thickness between alcohol use disorder patients and control subjects across multiple studies. Each horizontal line represents an individual study, with the center of the box indicating the SMD and the horizontal line representing the 95% confidence interval (CI). Bottom line: Results for RE Model with residual heterogeneity (QE), degrees of freedom (df), corresponding p-value (p), I^2 and t^2 statistics. Diamond shape at the bottom shows result model result as SMD. Numbers to the right of the study ID signifies the number of participants, mean and SD of pRNFL thickness per group.*

**Opiate Use Disorder**

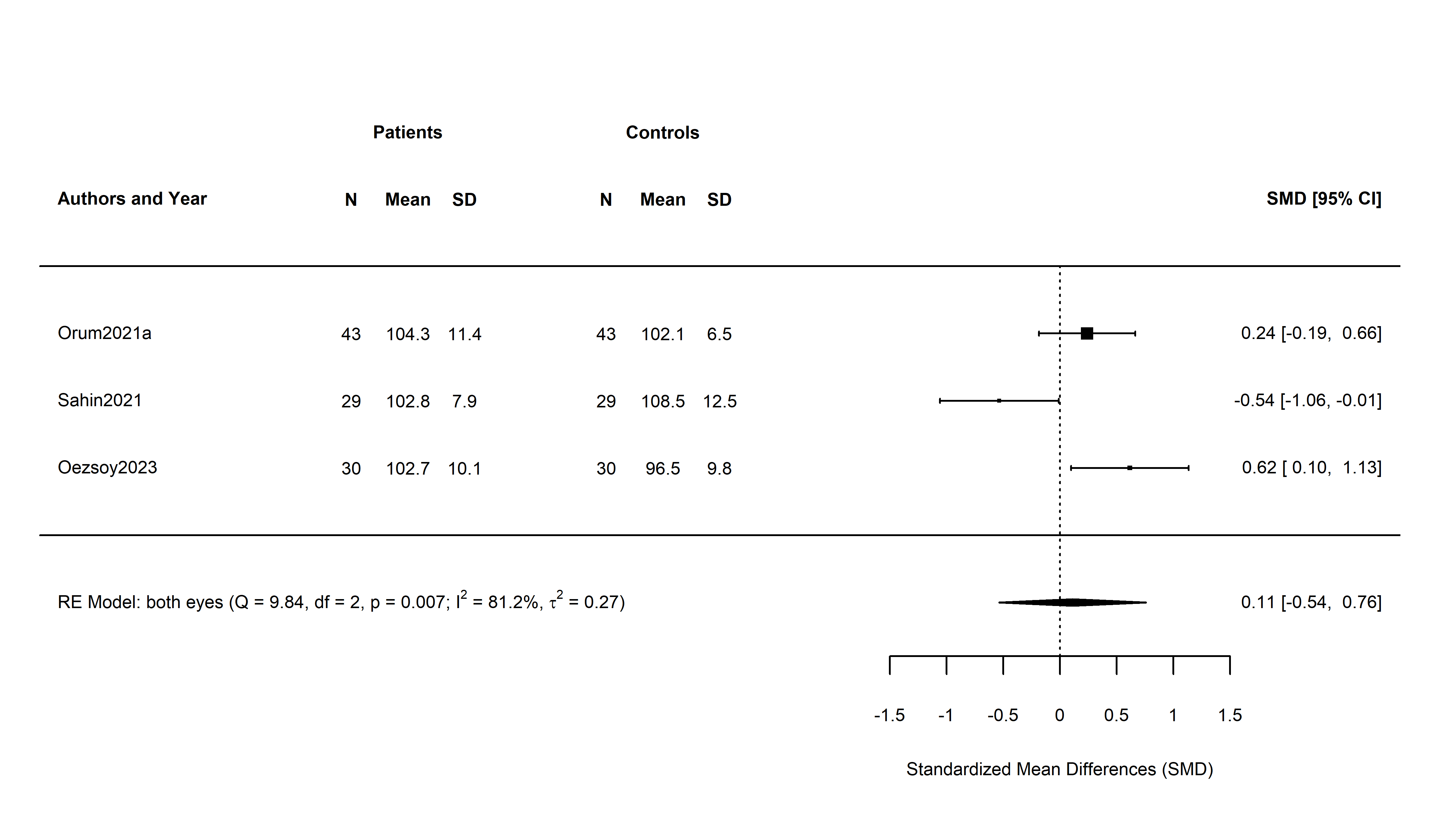

*Figure S 8 Forest plot of a meta-analysis comparing pRNFL in patients with opiate use disorder (OUD) to controls. This forest plot illustrates the standardized mean differences (SMD) in RNFL thickness between opiate use disorder patients and control subjects across multiple studies. Each horizontal line represents an individual study, with the center of the box indicating the SMD and the horizontal line representing the 95% confidence interval (CI). Bottom line: Results for RE Model with residual heterogeneity (QE), degrees of freedom (df), corresponding p-value (p), I^2 and t^2 statistics. Diamond shape at the bottom shows result model result as SMD. Numbers to the right of the study ID signifies the number of participants, mean and SD of pRNFL thickness per group.*

**Obsessive Compulsive Disorder**

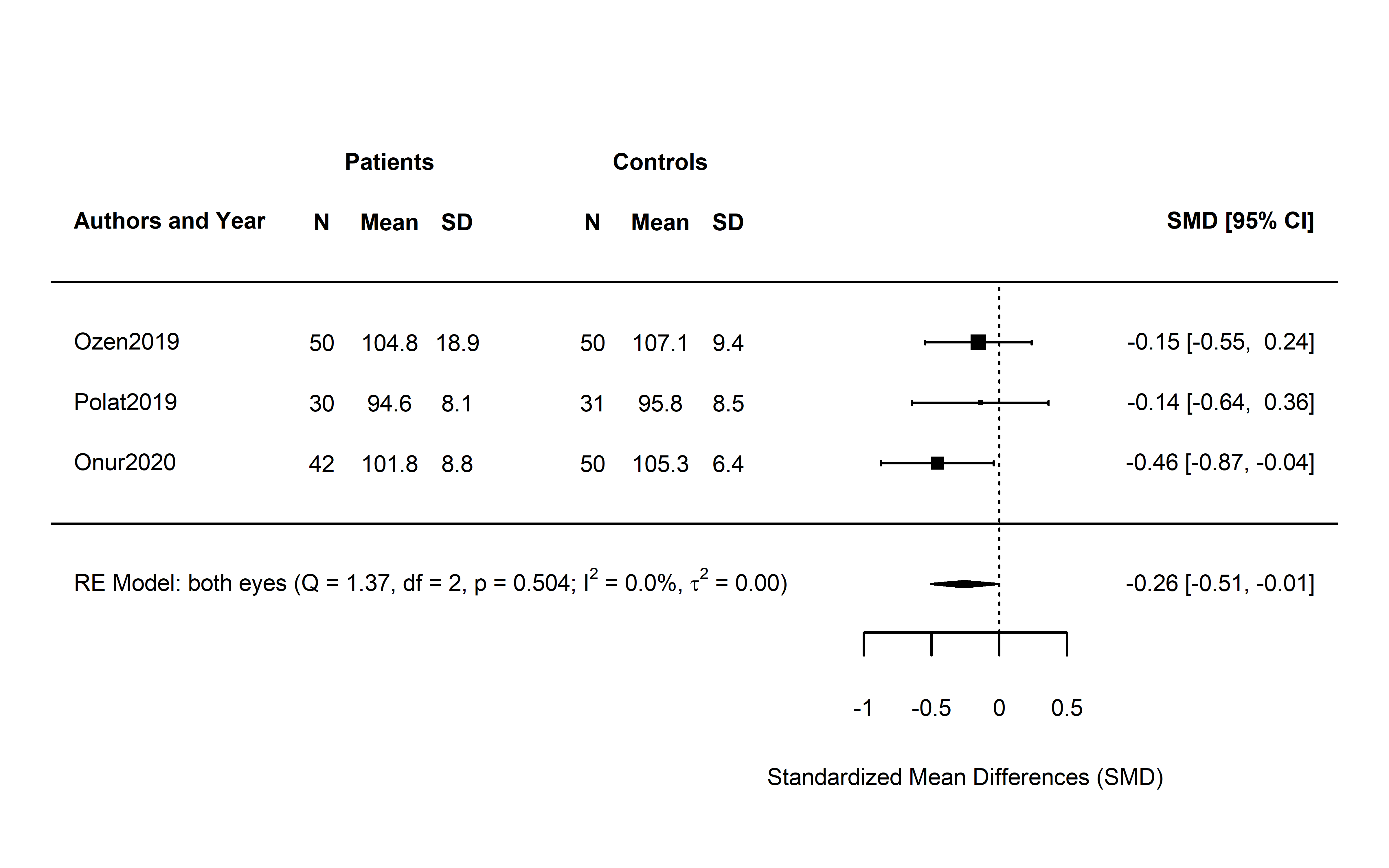

*Figure S 9 Forest plot of a meta-analysis comparing pRNFL thickness in patients with obsessive compulsive disorder (OCD) to controls. This forest plot illustrates the standardized mean differences (SMD) in RNFL thickness between opiate use disorder patients and control subjects across multiple studies. Each horizontal line represents an individual study, with the center of the box indicating the SMD and the horizontal line representing the 95% confidence interval (CI). Bottom line: Results for RE Model with residual heterogeneity (QE), degrees of freedom (df), corresponding p-value (p), I^2 and t^2 statistics. Diamond shape at the bottom shows result model result as SMD. Studies are identified by the first author's last name and year of publication. N Numbers to the right of the study ID signifies the number of participants, mean and SD of pRNLF thickness per group.*

**pRNFL, all quadrants – individual forest plots**

**Schizophrenia Spectrum Disorder**

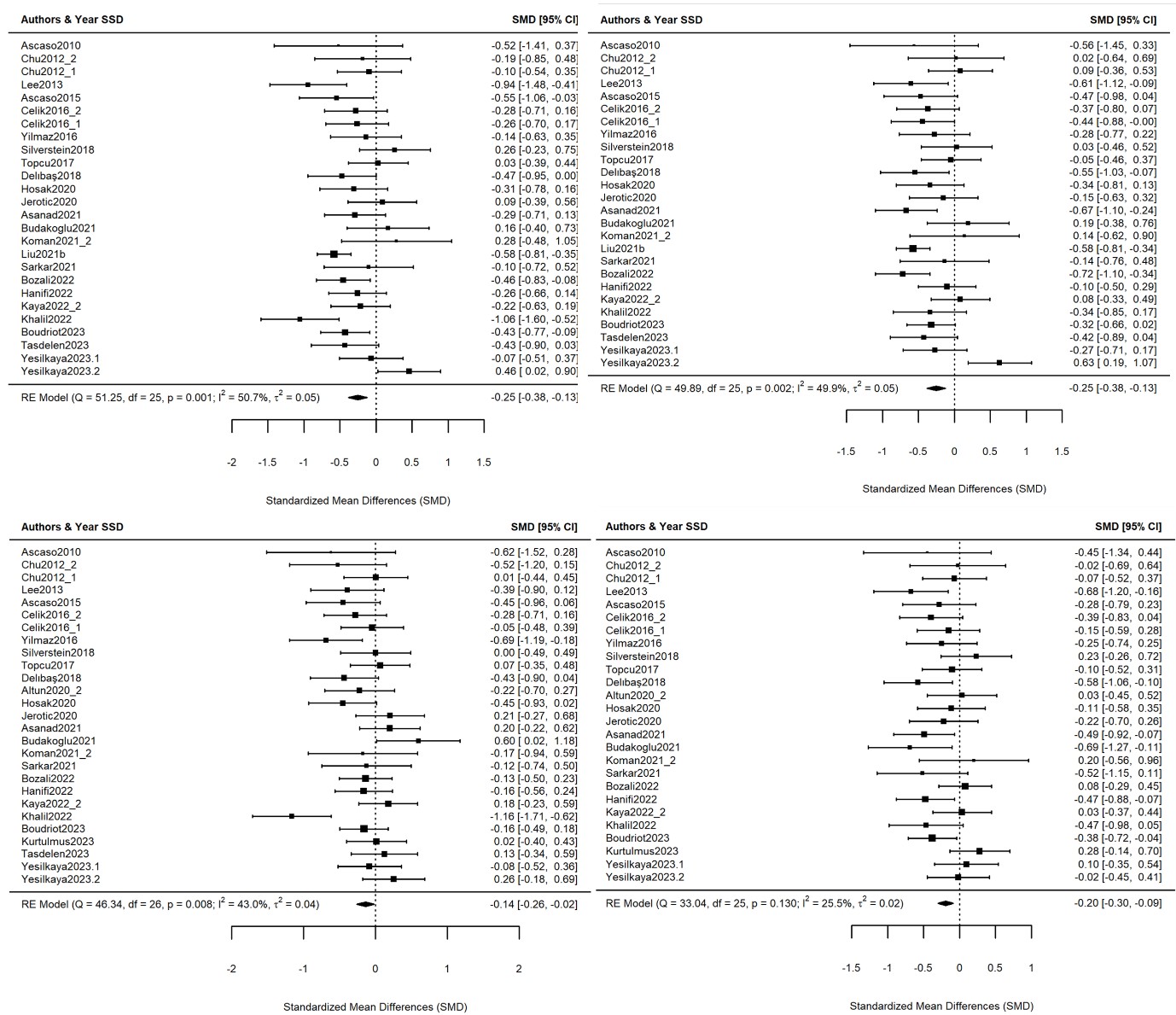

*Figure S 10 Forest plots for each pRNFL quadrant for schizophrenia spectrum disorder. Superior quadrant (upper left), Inferior quadrant (upper right), nasal quadrant (lower left) and temporal quadrant (lower right). Bottom line: Results for RE Model with residual heterogeneity (QE), degrees of freedom (df), corresponding p-value (p), I^2 and t^2 statistics. Diamond shape at the bottom shows result model result as SMD. Studies are identified by the first author's last name and year of publication. Numbers to the right of the study ID signifies the number of participants, mean and SD of pRNFL quadrant thickness per group.*

**Bipolar Disorder**

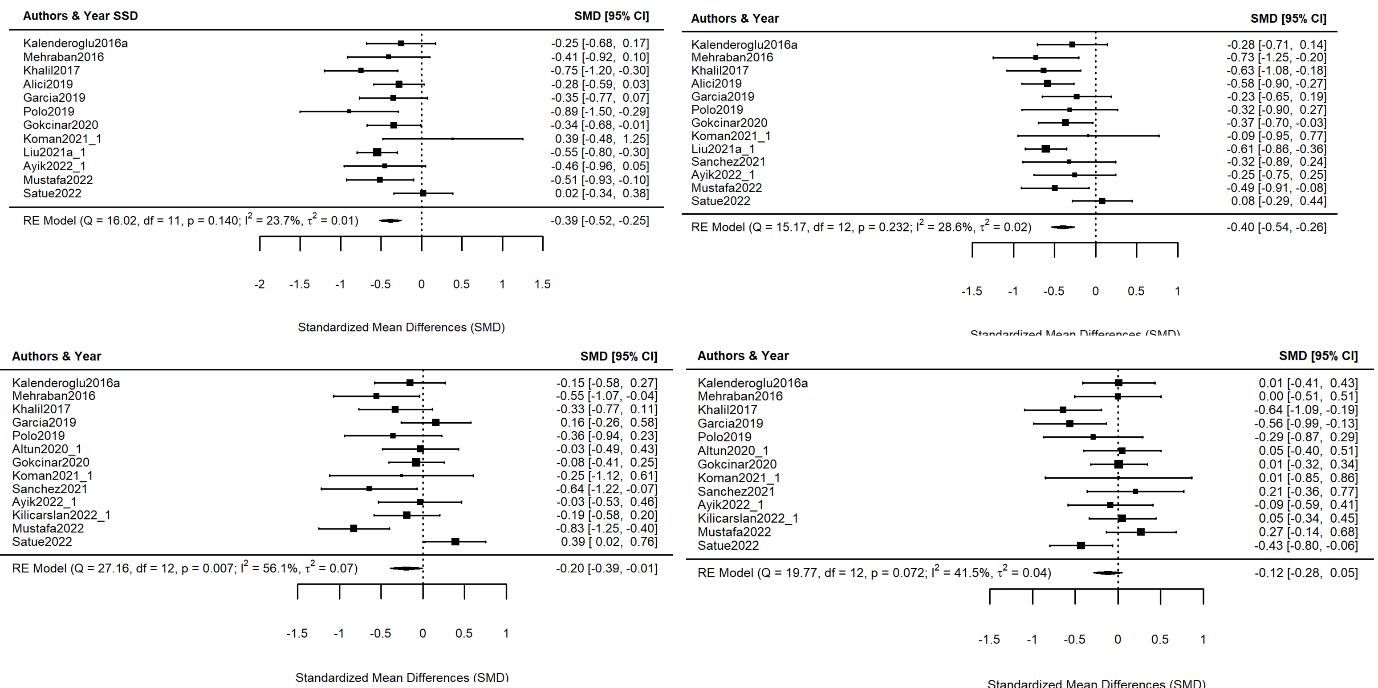

*Figure S 11 Forest plot for each pRNFL quadrant for bipolar disorder: superior (upper left), inferior (upper right), nasal (lower left) and temporal (lower right). Bottom line: Results for RE Model with residual heterogeneity (QE), degrees of freedom (df), corresponding p-value (p), I^2 and t^2 statistics. Diamond shape at the bottom shows result model result as SMD. Studies are identified by the first author's last name and year of publication. Numbers to the right of the study ID signifies the number of participants, mean and SD of pRNFL per group.*

**Major Depression**

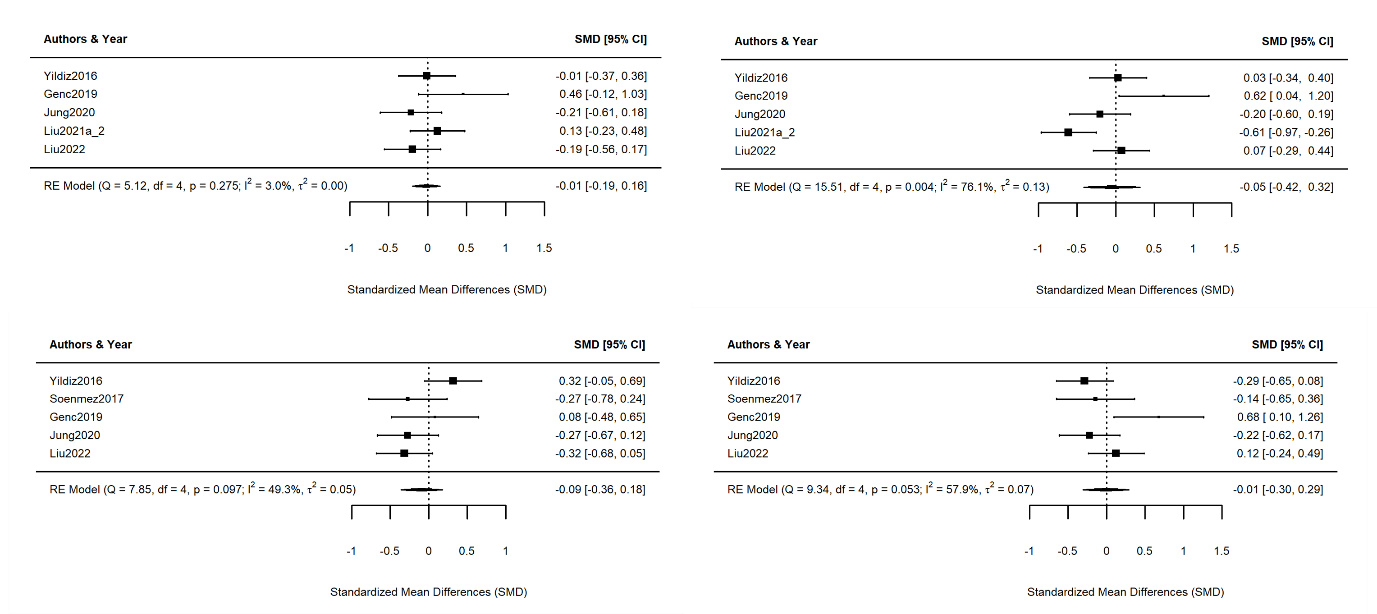

*Figure S 12 Forest plot for each pRNFL quadrant for major depressive disorder: superior (upper left), inferior (upper right), nasal (lower left) and temporal (lower right). Bottom line of each quadrant shows results for RE Model with residual heterogeneity (QE), degrees of freedom (df), corresponding p-value (p), I^2 and t^2 statistics. Diamond shape at the bottom shows result model result as SMD. Studies are identified by the first author's last name and year of publication. Numbers to the right of the study ID signifies the number of participants, mean and SD of pRNFL thickness per group.*

**Macular Thickness**

**Schizophrenia Spectrum Disorder**

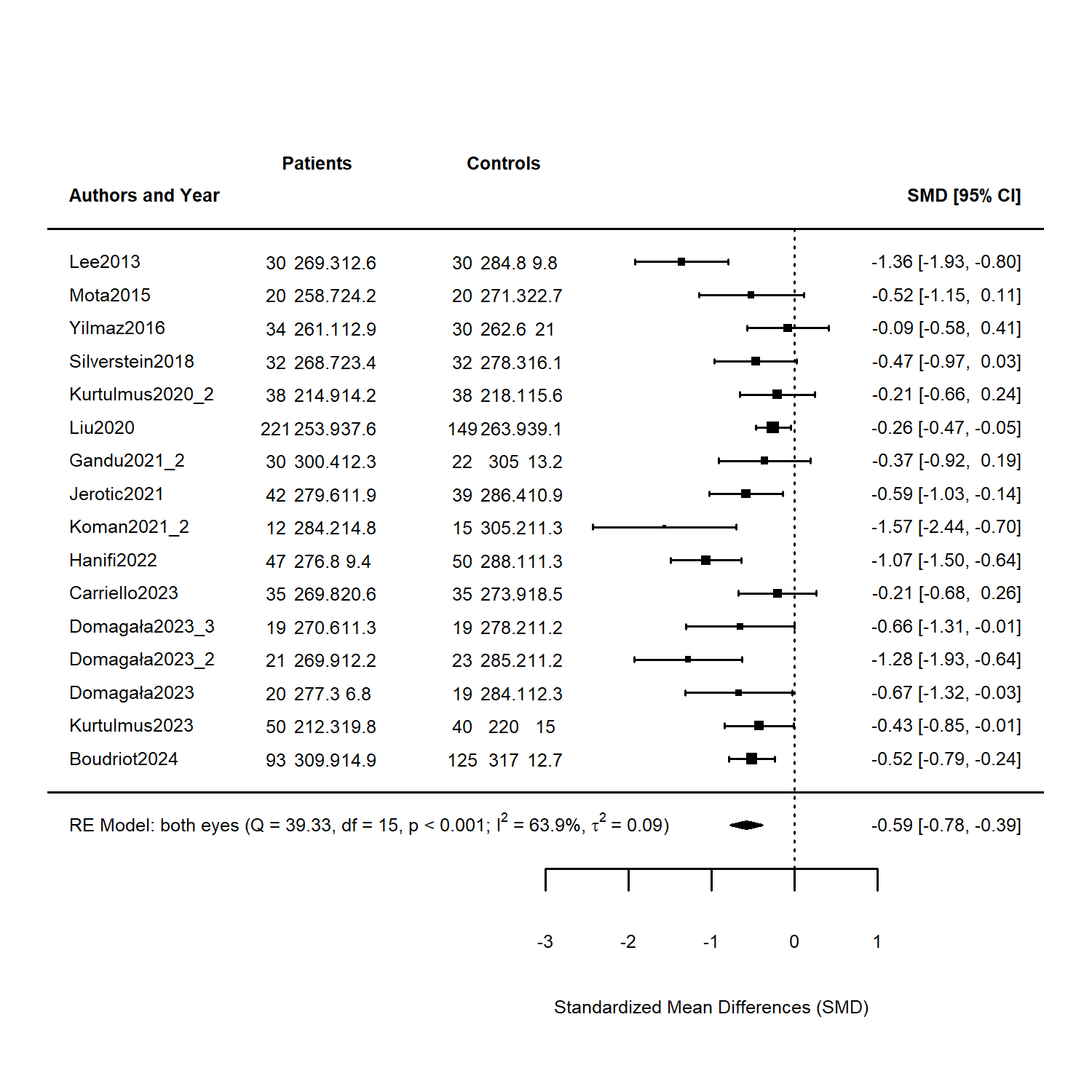

*Figure S 13 macular thickness – SSD: Forest plot for macular thickness in schizophrenia spectrum disorder. 16 samples from a total of 14 clinical studies were included. Bottom line: Results for RE Model with residual heterogeneity (QE), degrees of freedom (df), corresponding p-value (p), I^2 and t^2 statistics. Diamond shape at the bottom shows result model result as SMD. Studies are identified by the first author's last name and year of publication. Numbers to the right of the study ID signifies the number of participants, mean and SD of macular thickness per group. Domagala 2023 compared three independently sampled age groups with corresponding controls: 20-30 years (Domagala2023.1), 32-45 years (Domagala2023.2) and 45-60 years (Domagala2023.3)*

**Bipolar Disorder**

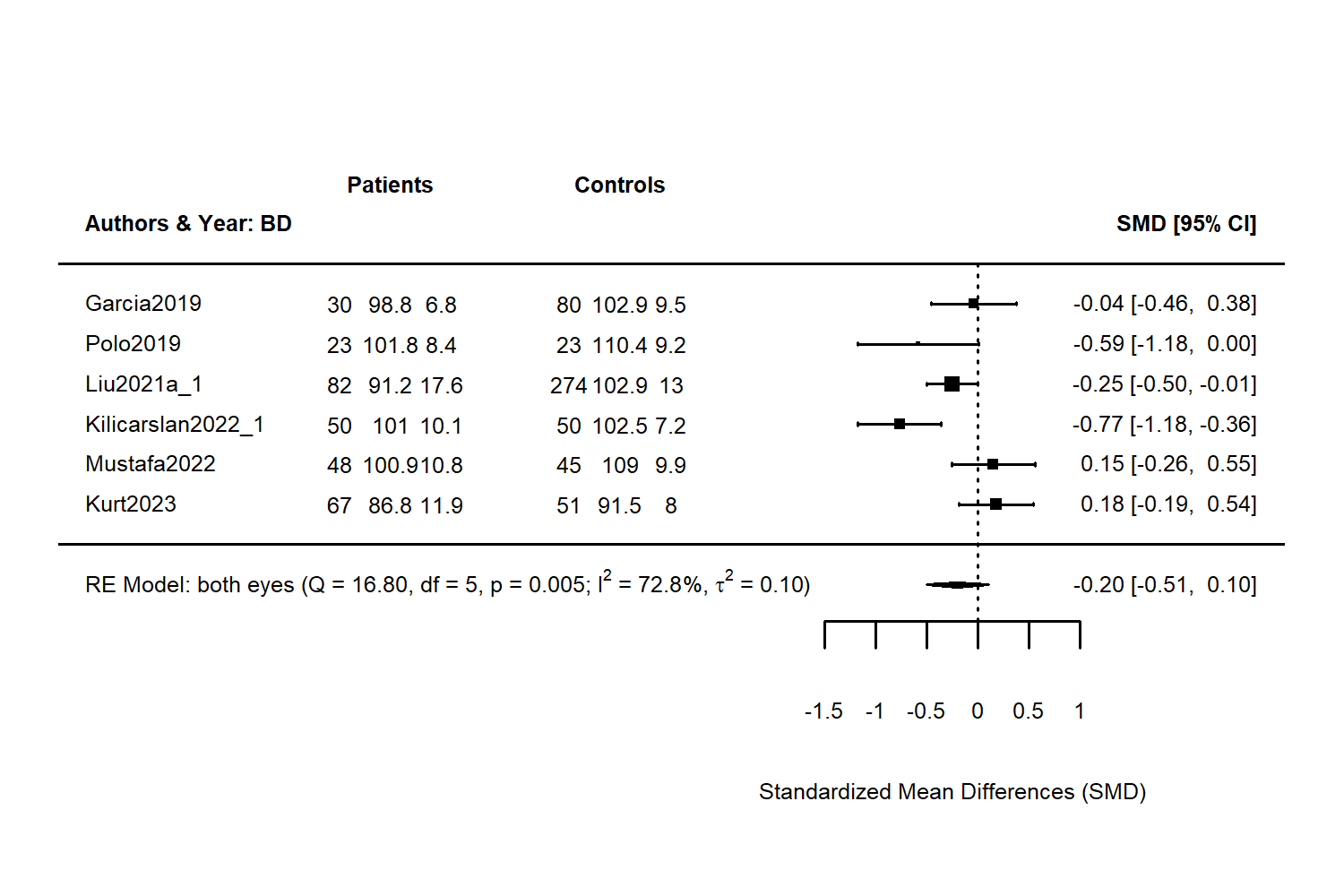

*Figure S 14 macular thickness – bipolar disorder: results of meta-analysis for bipolar disorder for macular thickness. Each layer corresponds to a study, the final result is based on a random effects model. Second row is the number of patients included and corresponding mean macular thickness and SD for Patients, third row shows number of controls, the mean and SD for controls. Fourth row shows a visual representation of outcomes of each study with corresponding 95% CI. Bottom line: Results for RE Model with residual heterogeneity (QE), degrees of freedom (df), corresponding p-value (p), I^2 and t^2 statistics. Diamond shape at the bottom shows result model result as SMD. Studies are identified by the first author's last name and year of publication. Numbers to the right of the study ID signifies the number of participants, mean and SD of macular thickness per group.*

**Major Depressive Disorder**

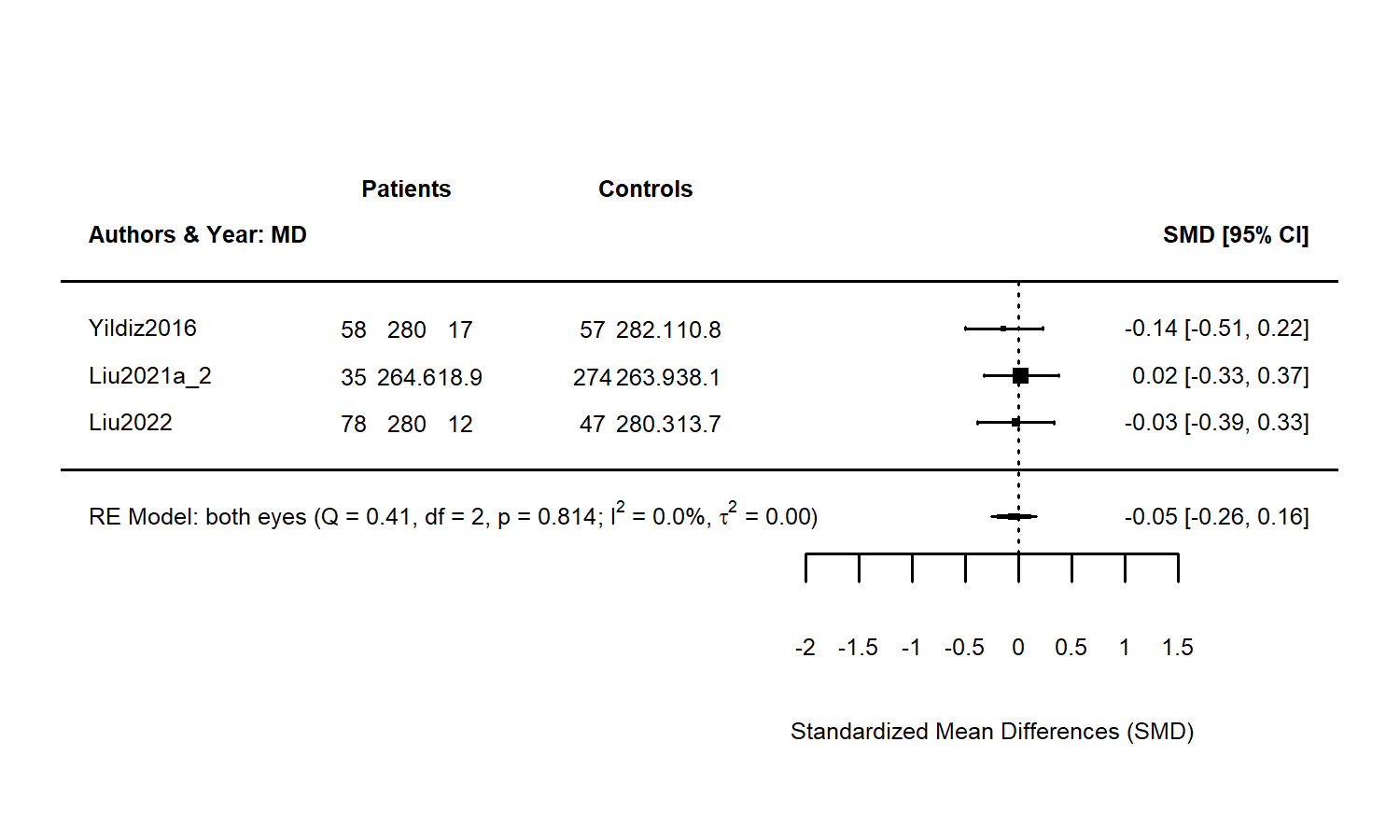

*Figure S 15 Macular thickness – major depressive disorder (MDD). Forest plot of a meta-analysis comparing macula thickness in patients with MDD to controls. This forest plot illustrates the standardized mean differences (SMD) in RNFL thickness between MDD patients and control subjects across multiple studies. Bottom line: Results for RE Model with residual heterogeneity (QE), degrees of freedom (df), corresponding p-value (p), I^2 and t^2 statistics. Diamond shape at the bottom shows result model result as SMD, Each horizontal line represents an individual study, with the center of the box indicating the SMD and the horizontal line representing the 95% confidence interval (CI). Studies are identified by the first author's last name and year of publication. Numbers to the right of the study ID signifies the number of participants, mean and SD of macular thickness per group.*

**Macular Subfields**

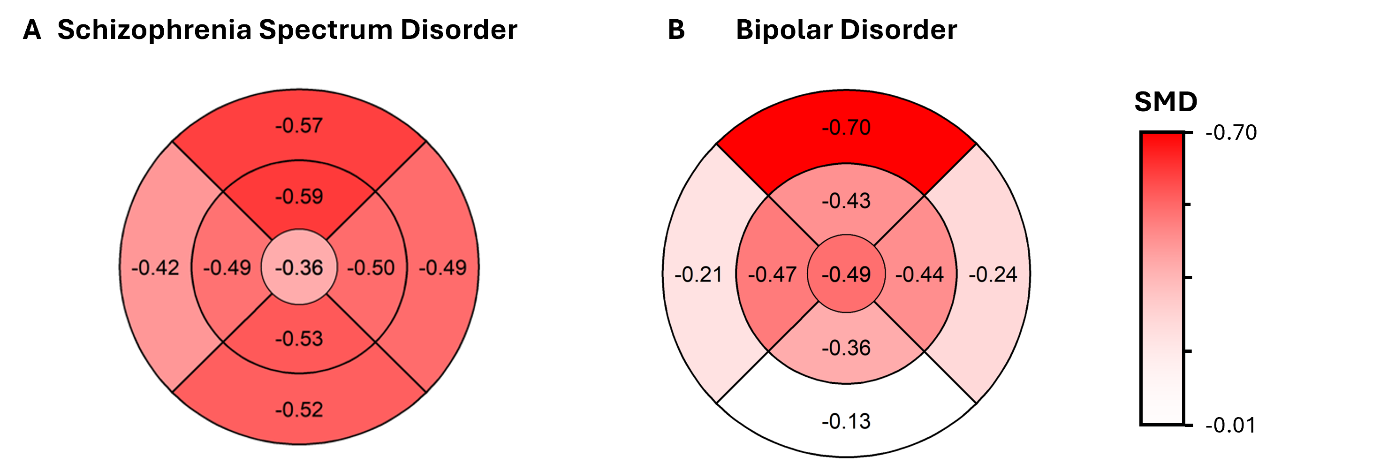

*Figure S 16 Overview of standardized mean differences (SMD) in macula subfields across all studies investigating Schizophrenia Spectrum disorder (left) and bipolar disorder (right) respectively. Starting on the top sector and going clockwise: outer superior and inner superior (top), outer temporal and inner temporal (right), outer inferior and inner inferior (bottom) and outer nasal and inner nasal (left) with central foveal subfield in the center.*

**Schizophrenia Spectrum Disorder – Inner Ring**

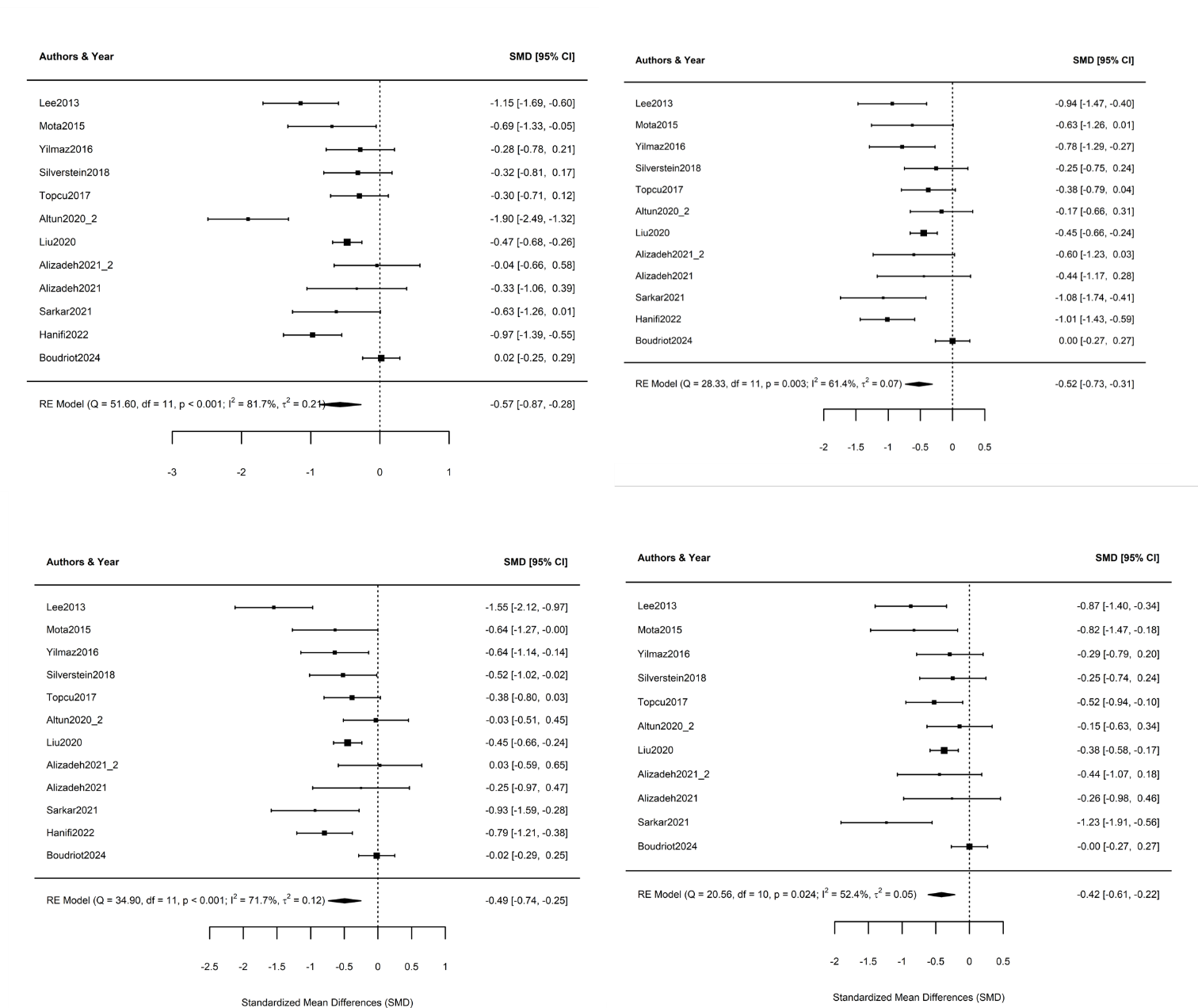

*Figure S 17 Forest plots for macular subfields in schizophrenia spectrum disorder for the inner ring: inner superior subfield (upper left), inner inferior subfield (upper right), inner nasal subfield (lower left), and inner temporal subfield (lower right).*

**Schizophrenia Spectrum Disorders – Outer Ring**

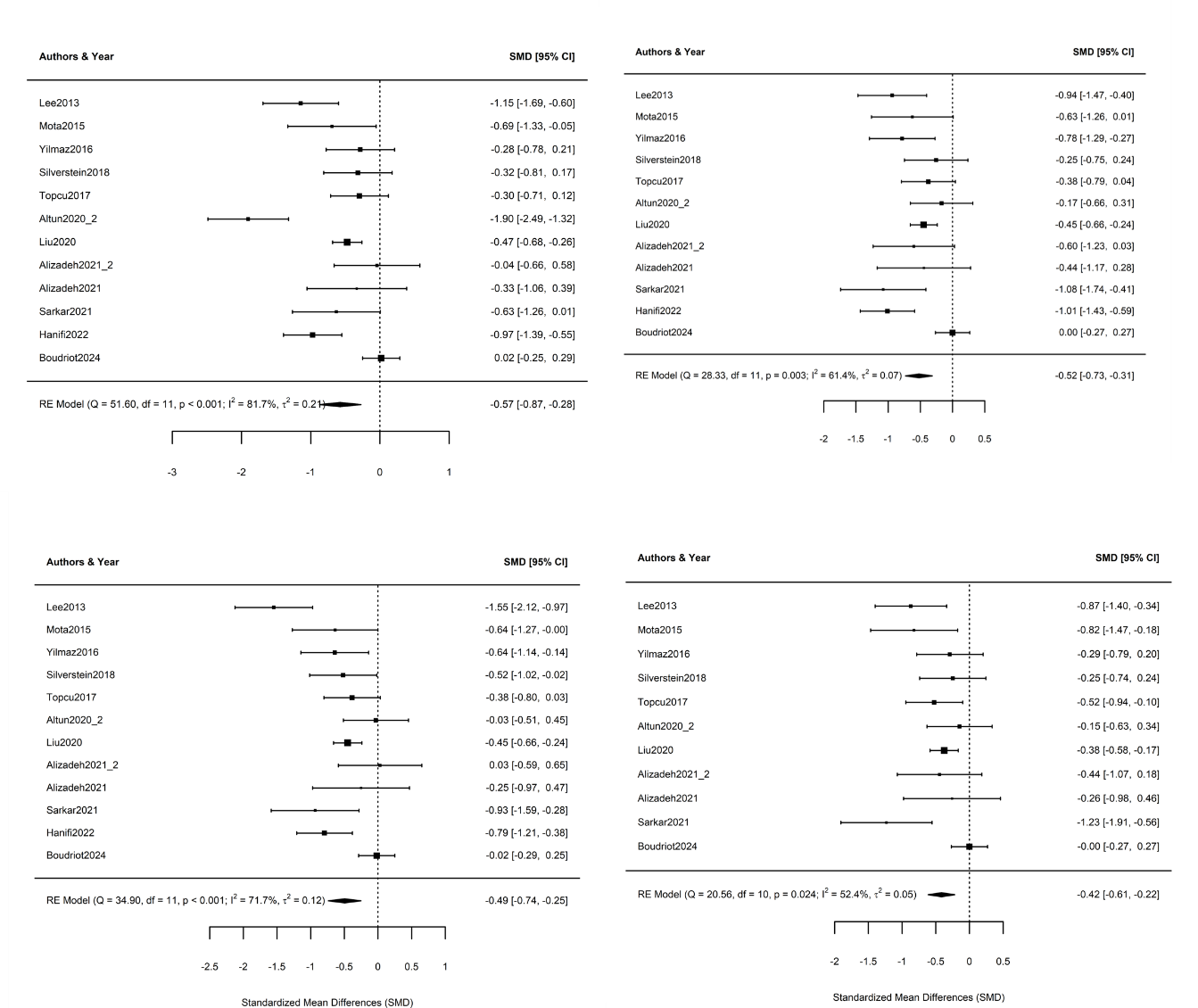

*Figure S 18 Forest plots for macular subfields in schizophrenia spectrum disorder for the outer ring: outer superior subfield (upper left), outer inferior subfield (upper right), outer nasal subfield (lower left), and outer temporal subfield (lower right).*

**Bipolar Disorder – Inner Ring**

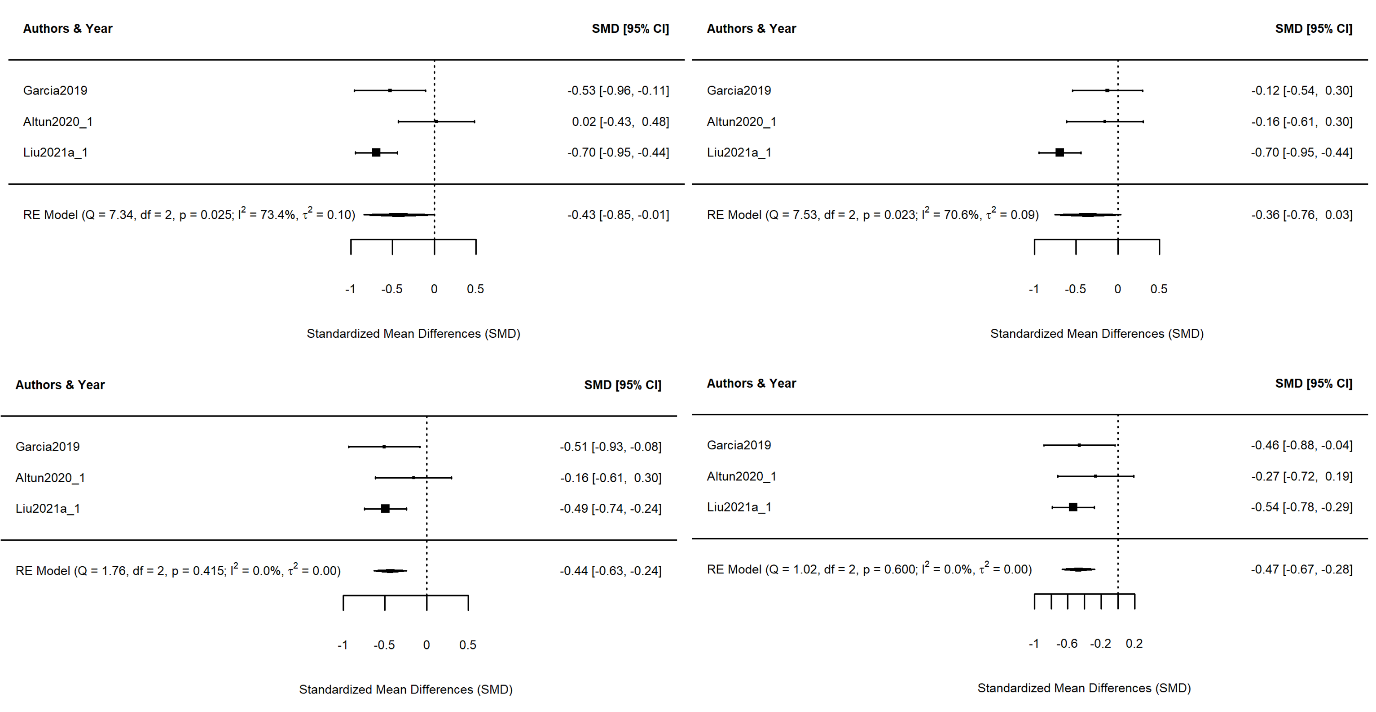

*Figure S 19 Forest plots for macular subfields in bipolar disorder for the inner ring: inner superior subfield (upper left), inner inferior subfield (upper right), inner nasal subfield (lower left), and inner temporal subfield (lower right).*

**Bipolar Disorder – Outer Ring**

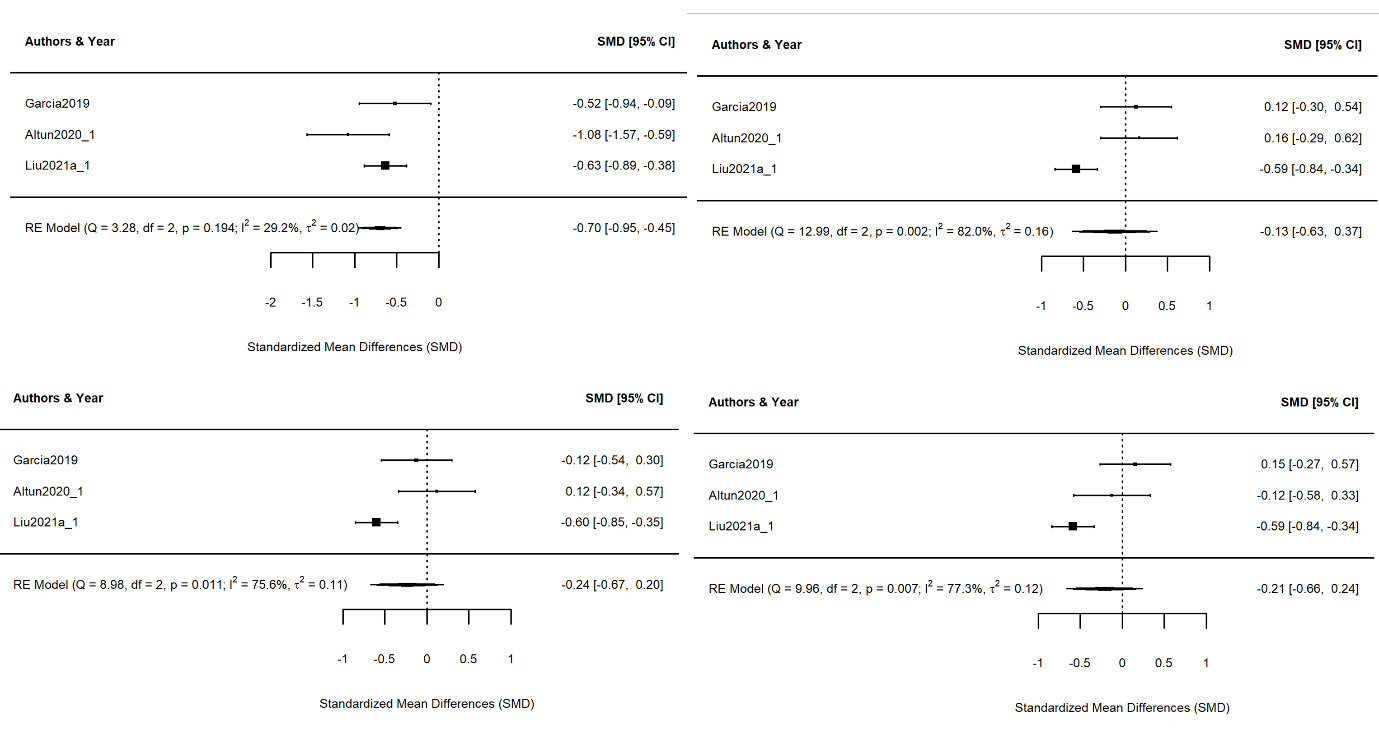

*Figure S 20 Forest plots for macular subfields in bipolar disorder for the outer ring: outer superior subfield (upper left), outer inferior subfield (upper right), outer nasal subfield (lower left), and outer temporal subfield (lower right).*

**Macula Volume**

**Schizophrenia Spectrum Disorder**

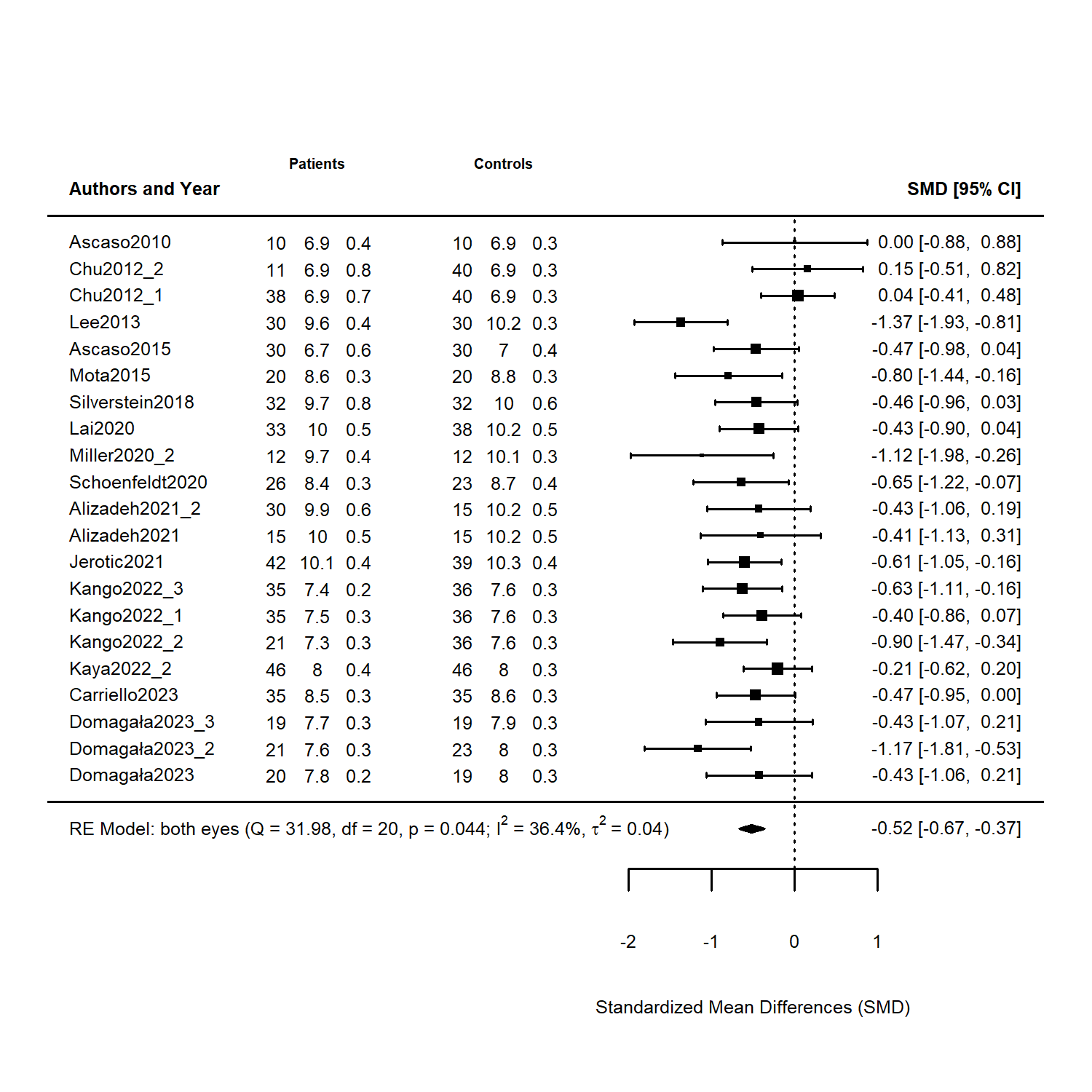

*Figure S 21 Macula Volume in schizophrenia spectrum disorder. Forest plot of a meta-analysis comparing the macular volume in schizophrenia spectrum disorder patients and healthy controls. Bottom line: Results for RE Model with residual heterogeneity (QE), degrees of freedom (df), corresponding p-value (p), I^2, and t^2 statistics. Diamond shape at the bottom shows result model result as SMD. Chu2012 compared 39 patients with schizophrenia (Chu2012.2) and 11 patients with schizoaffective disorder (Chu2012.1) to 40 controls. Domagala 2023 compared three different age groups: 20-30 years (Domagala2023.1), 32-45 years (Domagala2023.2) and 45-60 years (Domagala2023.3). Kango 2022 compared 35 patients with schizophrenia (Kango2022.1), 21 patients with first episode psychosis patients (Kango2022.2) and 35 patients with treatment resistant Schizophrenia to 36 controls.*

**GCL-IPL Schizophrenia Spectrum Disorder**

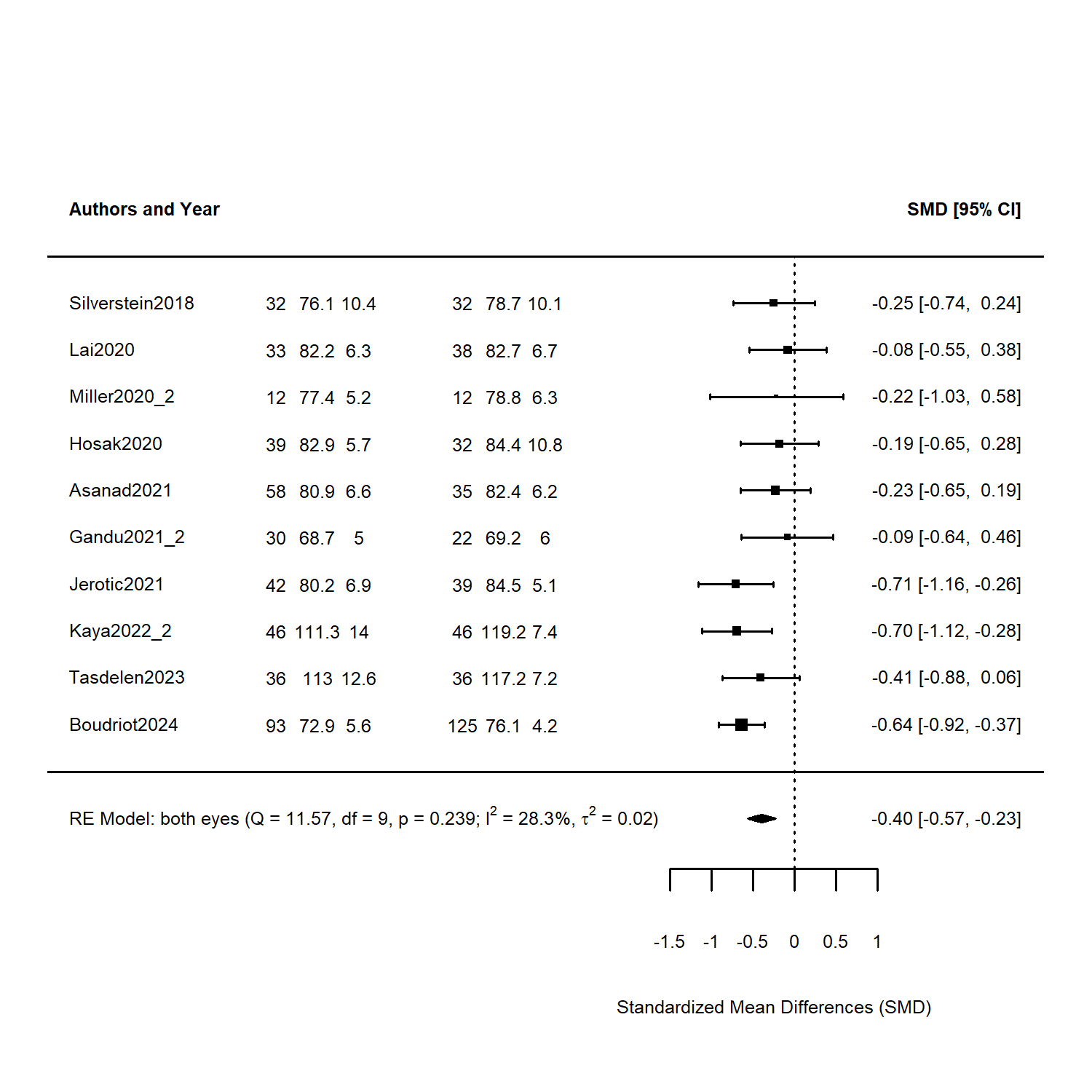

*Figure S 22 GCL-IPL in SSD. Forest plot of a meta-analysis comparing the GPL-IPL thickness in schizophrenia spectrum disorder to healthy controls. Bottom line: Results for RE Model with residual heterogeneity (QE), degrees of freedom (df), corresponding p-value (p), I^2, and t^2 statistics. Diamond shape at the bottom shows result model result as SMD. Studies are identified by the first author's last name and year of publication. Numbers to the right of the study ID signifies the number of participants, mean and SD of GCL-IPL-thickness per group.*

**Supplementary Figures 23 – 34: Funnel Plots**

**Schizophrenia Spectrum Disorder: pRNFL**

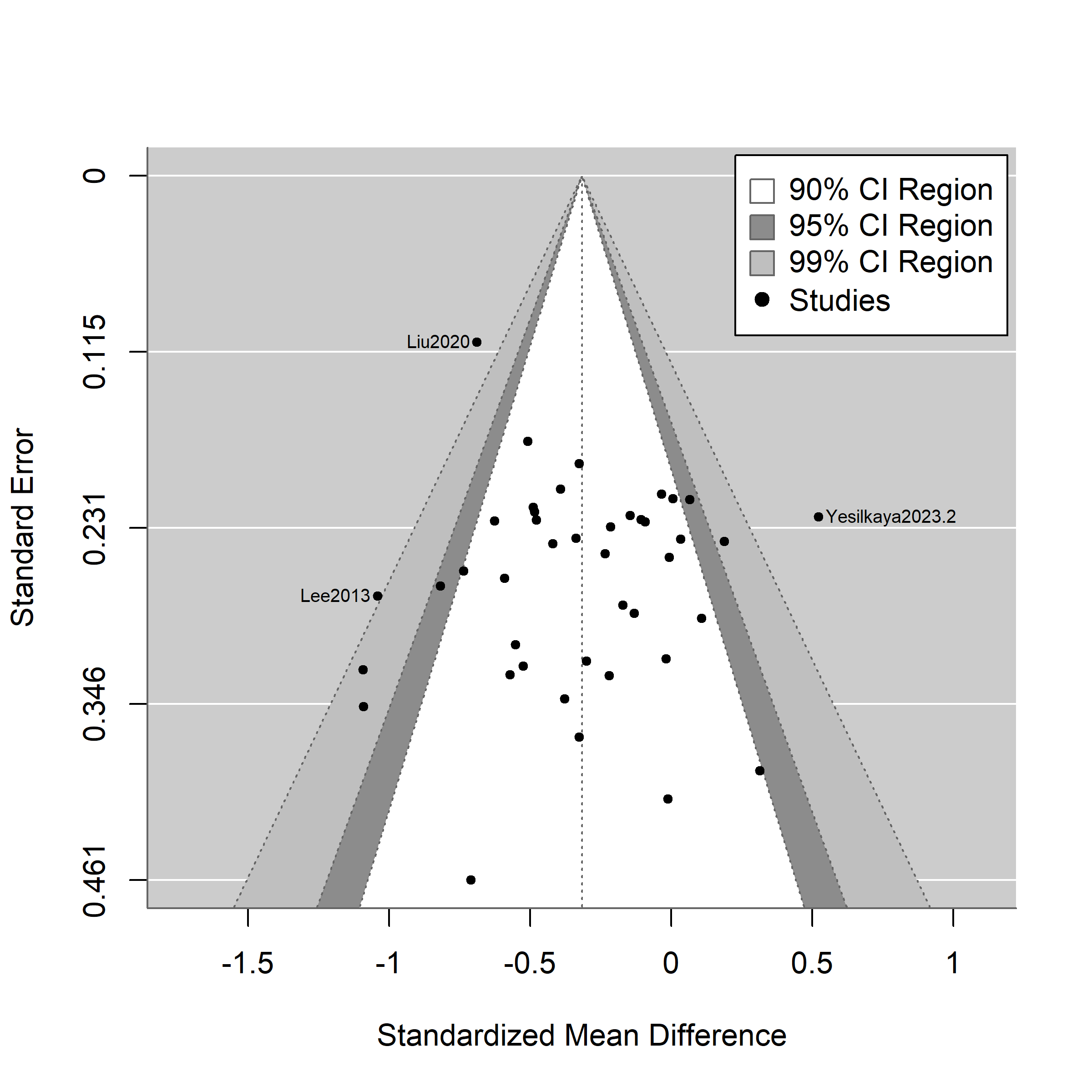

*Figure S 23 contour-enhanced funnel plot for schizophrenia spectrum disorder, RNFL. Every dot is a study, with studies outside the pseudo-confidence-region identified by Name of the first author and year of publication*

**Schizophrenia Spectrum Disorder: pRNFL Quadrants**

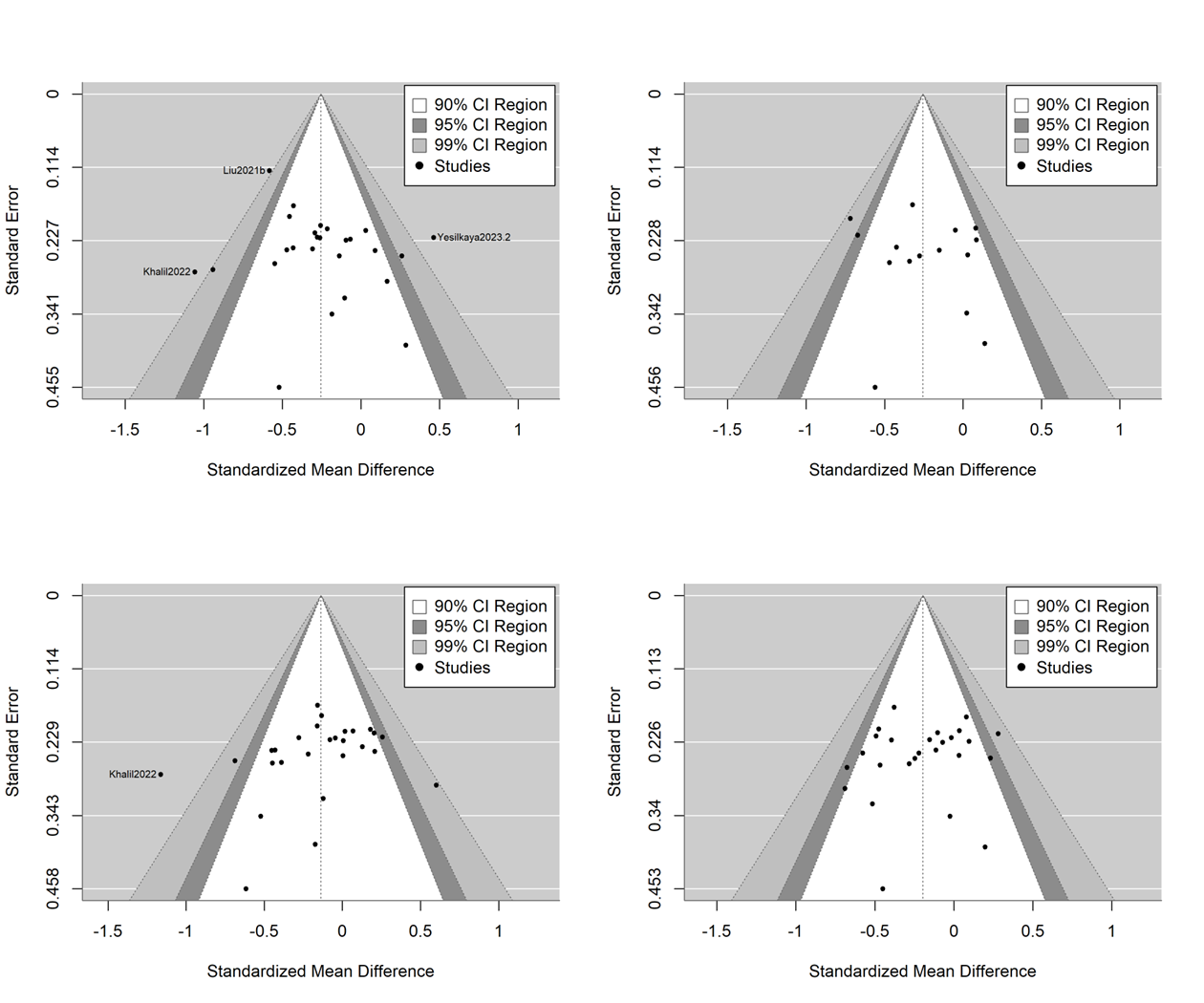

*Figure S 24* *contour-enhanced Funnel plots for schizophrenia spectrum disorder, pRNFL quadrants: Superior (upper left), inferior (upper right), nasal (lower left) and temporal (lower right)*

**Schizophrenia Spectrum Disorder: Macula Thickness**

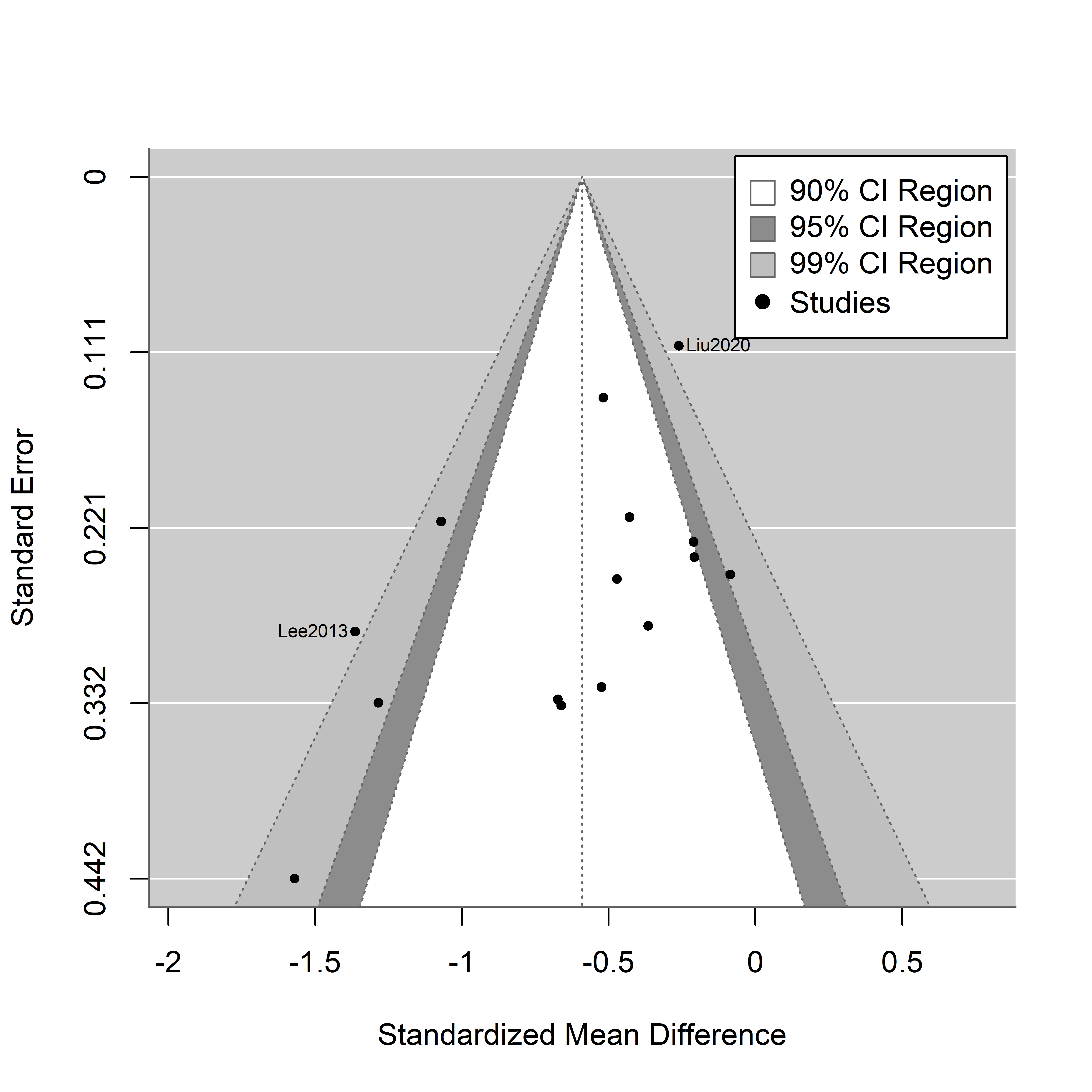

*Figure S 25 contour-enhanced funnel plot schizophrenia spectrum disorder, macula thickness A funnel plot shows the observed effect sizes or outcomes on the x-axis against some measure of precision of the observed effect sizes or outcomes on the y-axis, based on Sterne and Egger (2001). In the absence of publication bias and heterogeneity, one would then expect to see the points forming a funnel shape, with the majority of the points falling inside of the pseudo-confidence region (white). Every dot is a study, with studies outside the pseudo-confidence-region identified by Name of the first author and year of publication*

**Schizophrenia spectrum Disorder: Central Foveal Thickness**

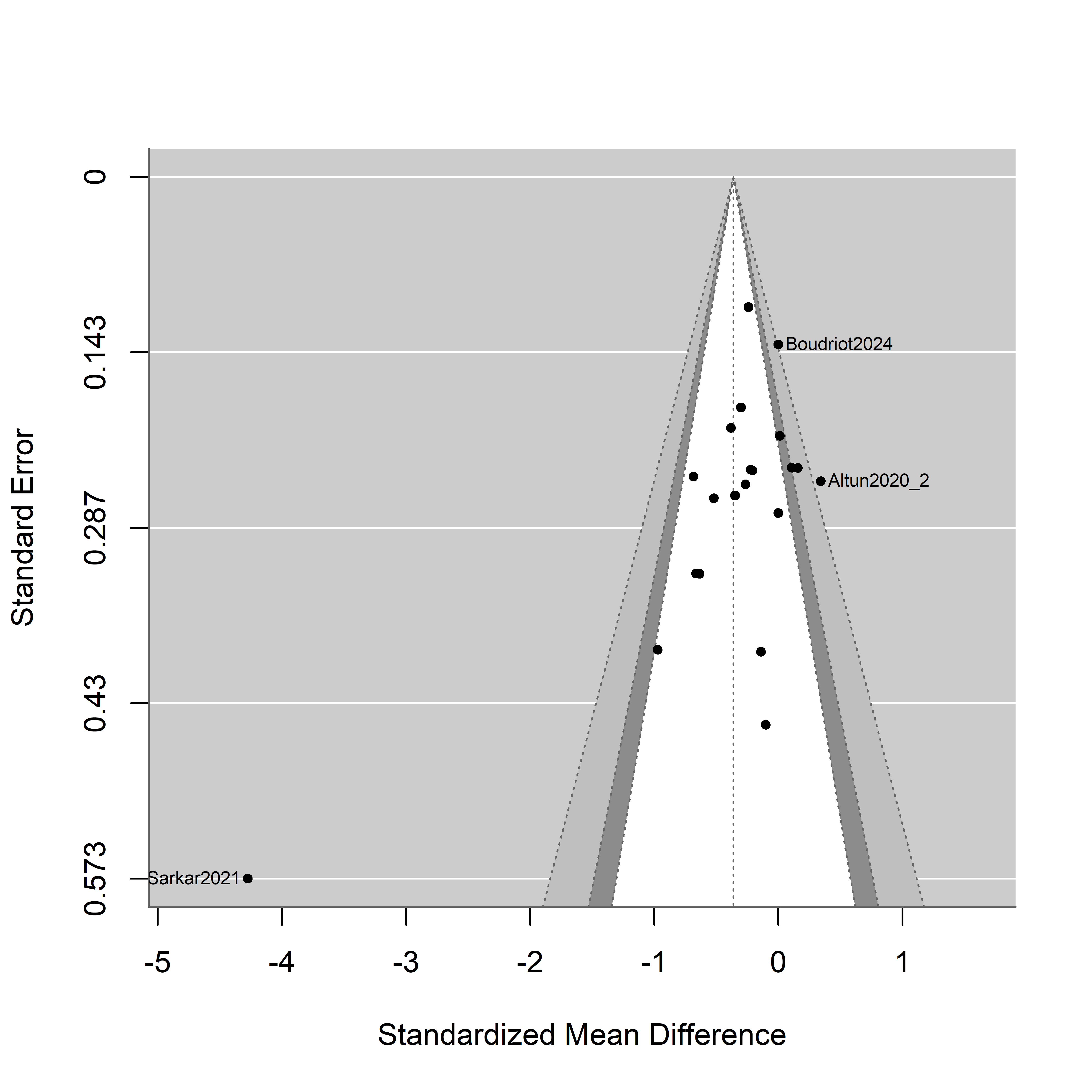

*Figure S 26 contour-enhanced funnel plot for schizophrenia spectrum disorder, central foveal macular thickness. A funnel plot shows the observed effect sizes or outcomes on the x-axis against some measure of precision of the observed effect sizes or outcomes on the y-axis, based on Sterne and Egger (2001). In the absence of publication bias and heterogeneity, one would then expect to see the points forming a funnel shape, with the majority of the points falling inside of the pseudo-confidence region (white). Every dot is a study, with studies outside the pseudo-confidence-region identified by Name of the first author and year of publication*

**Schizophrenia spectrum Disorder: Macular Subfields – Inner Ring**

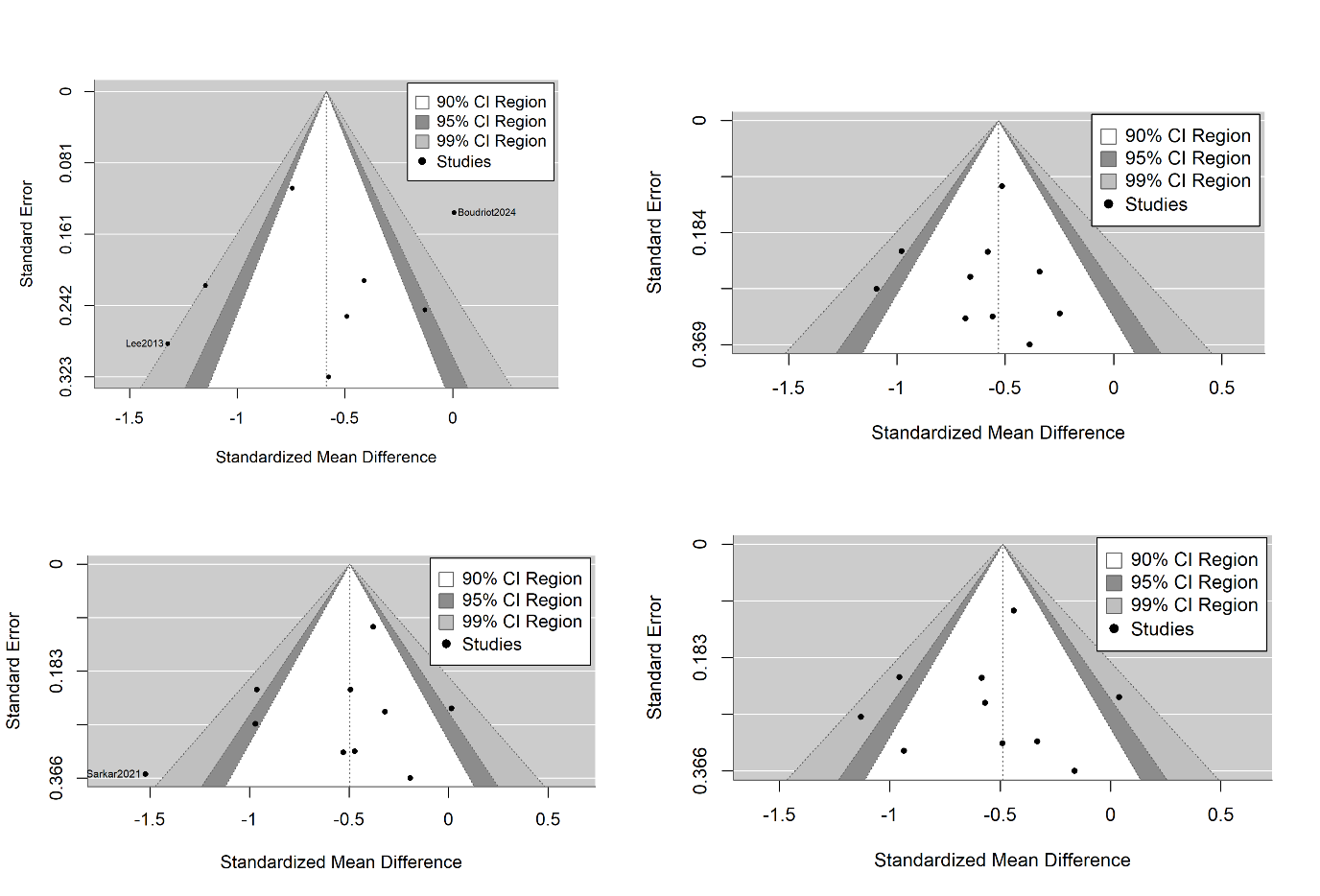

*Figure S 27 contour-enhanced Funnel plots for schizophrenia spectrum disorder, macula subfields, inner ring: Superior (upper left), inferior (upper right), nasal (lower left) and temporal (lower right)*

**Schizophrenia Spectrum Disorder: Macular Subfields – Outer Ring**

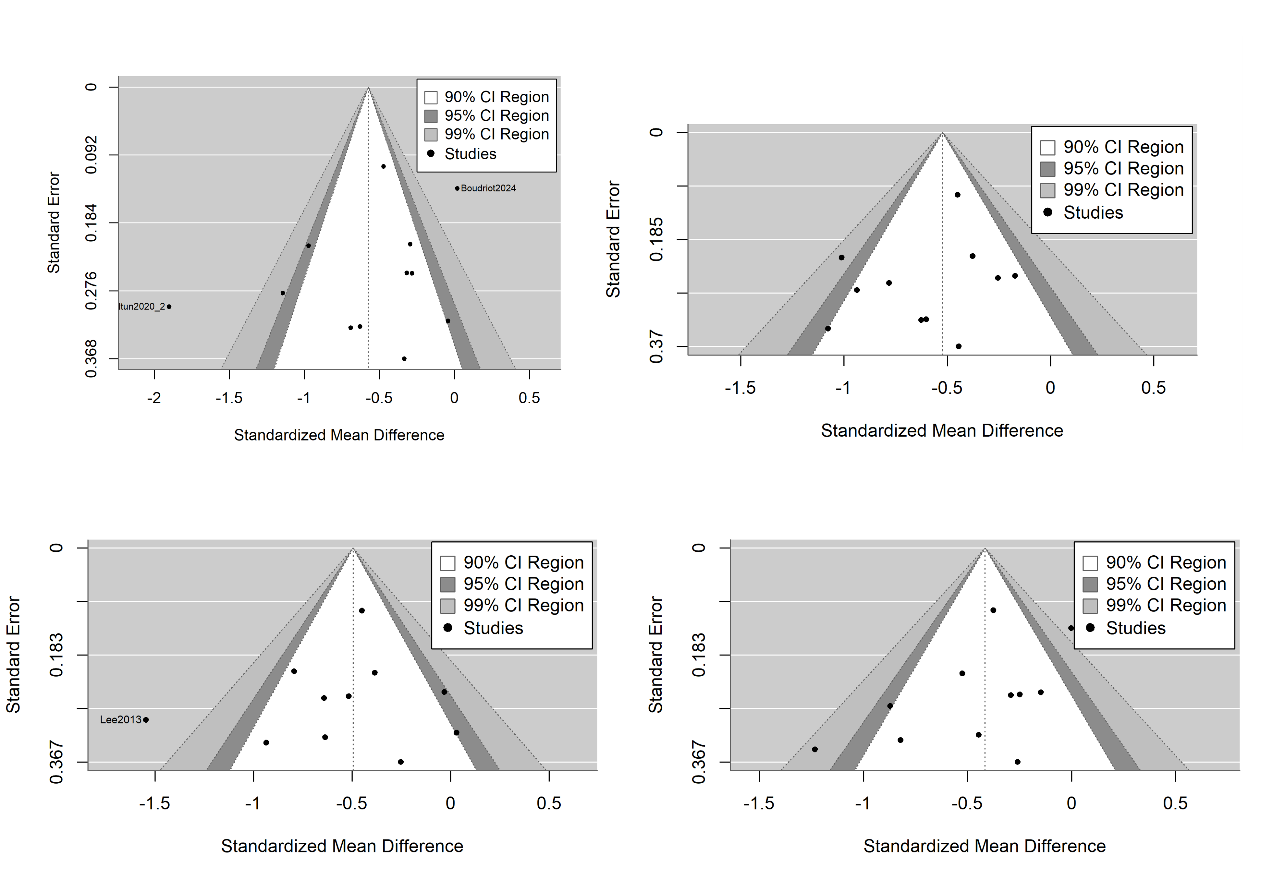

*Figure S 28 contour-contour-enhanced funnel plots for schizophrenia spectrum disorder, macula subfields, outer ring: Superior (upper left), inferior (upper right), nasal (lower left) and temporal (lower right)*

**Schizophrenia Spectrum Disorders: Macula volume**

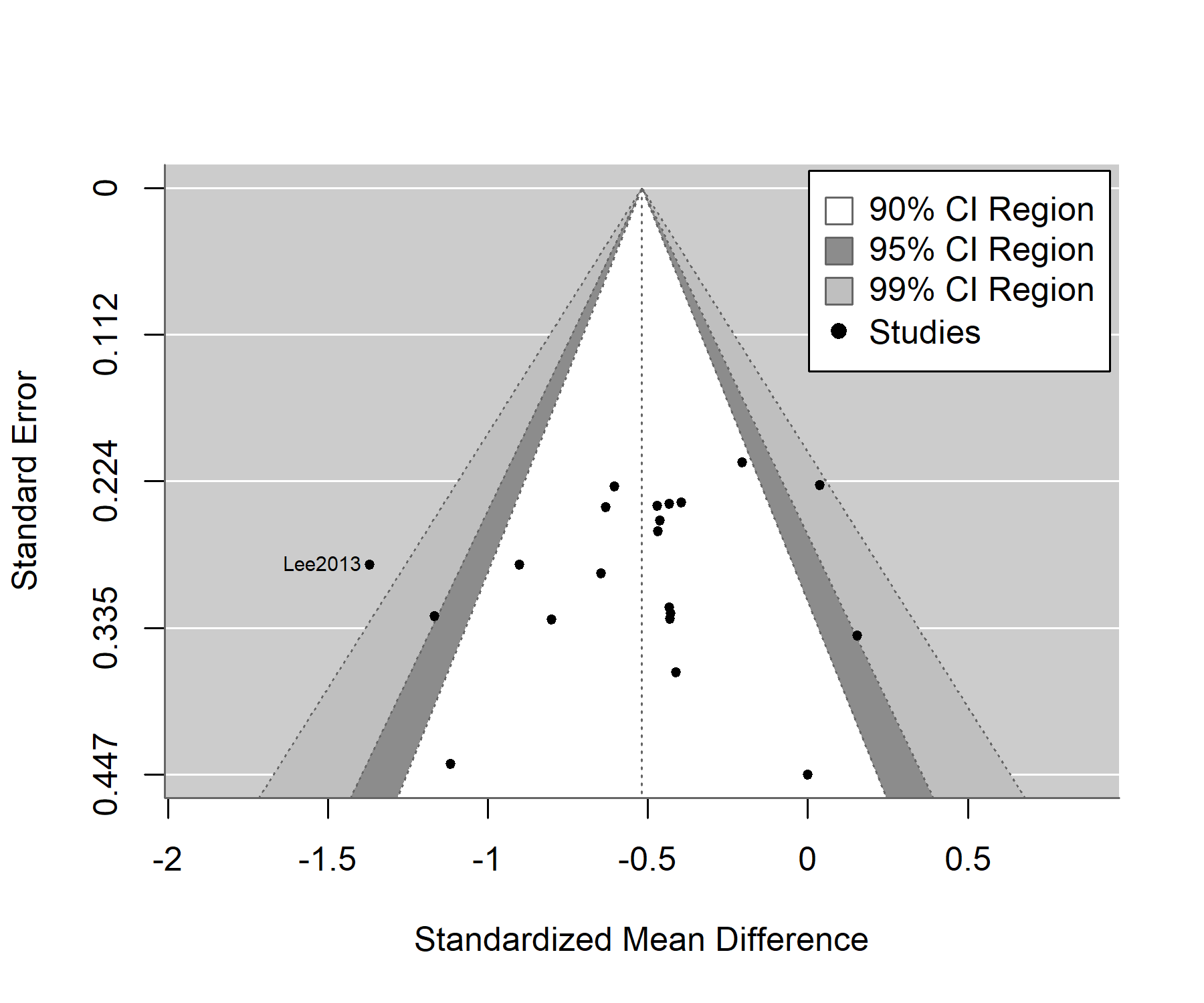

*Figure S29 contour-enhanced funnel plot for schizophrenia spectrum disorders, macular volume. A funnel plot shows the observed effect sizes or outcomes on the x-axis against some measure of precision of the observed effect sizes or outcomes on the y-axis, based on Sterne and Egger (2001). In the absence of publication bias and heterogeneity, one would then expect to see the points forming a funnel shape, with the majority of the points falling inside of the pseudo-confidence region (white). Every dot is a study, with studies outside the pseudo-confidence-region identified by Name of the first author and year of publication*

**Schizophrenia Spectrum Disorders: GCL-IPL**

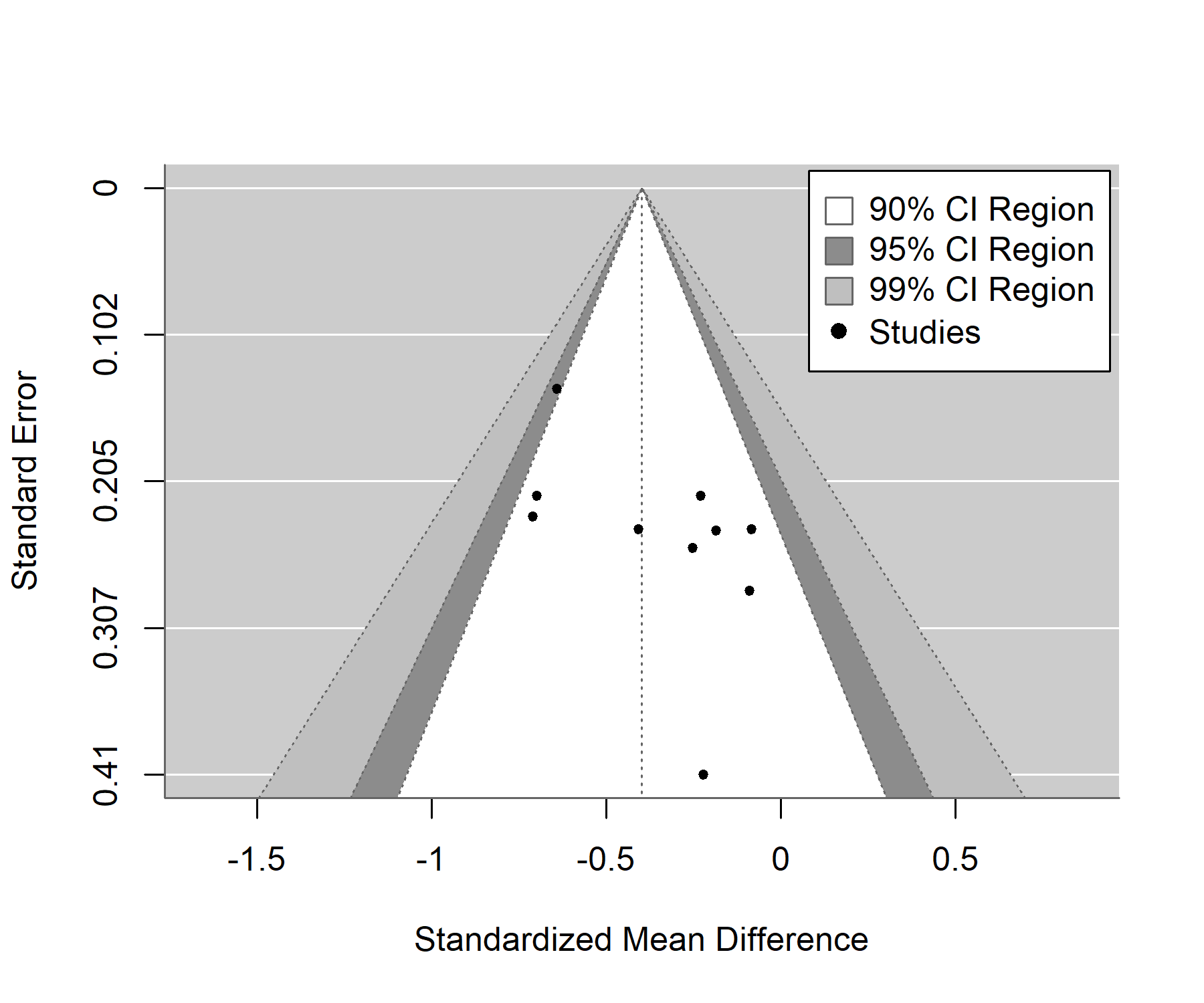

*Figure S 30 contour-enhanced funnel-plot for schizophrenia spectrum disorder, GCL-IPL A funnel plot shows the observed effect sizes or outcomes on the x-axis against some measure of precision of the observed effect sizes or outcomes on the y-axis, based on Sterne and Egger (2001). In the absence of publication bias and heterogeneity, one would then expect to see the points forming a funnel shape, with the majority of the points falling inside of the pseudo-confidence region (white). Every dot is a study, with studies outside the pseudo-confidence-region identified by Name of the first author and year of publication*

**Bipolar Disorder: pRNFL**

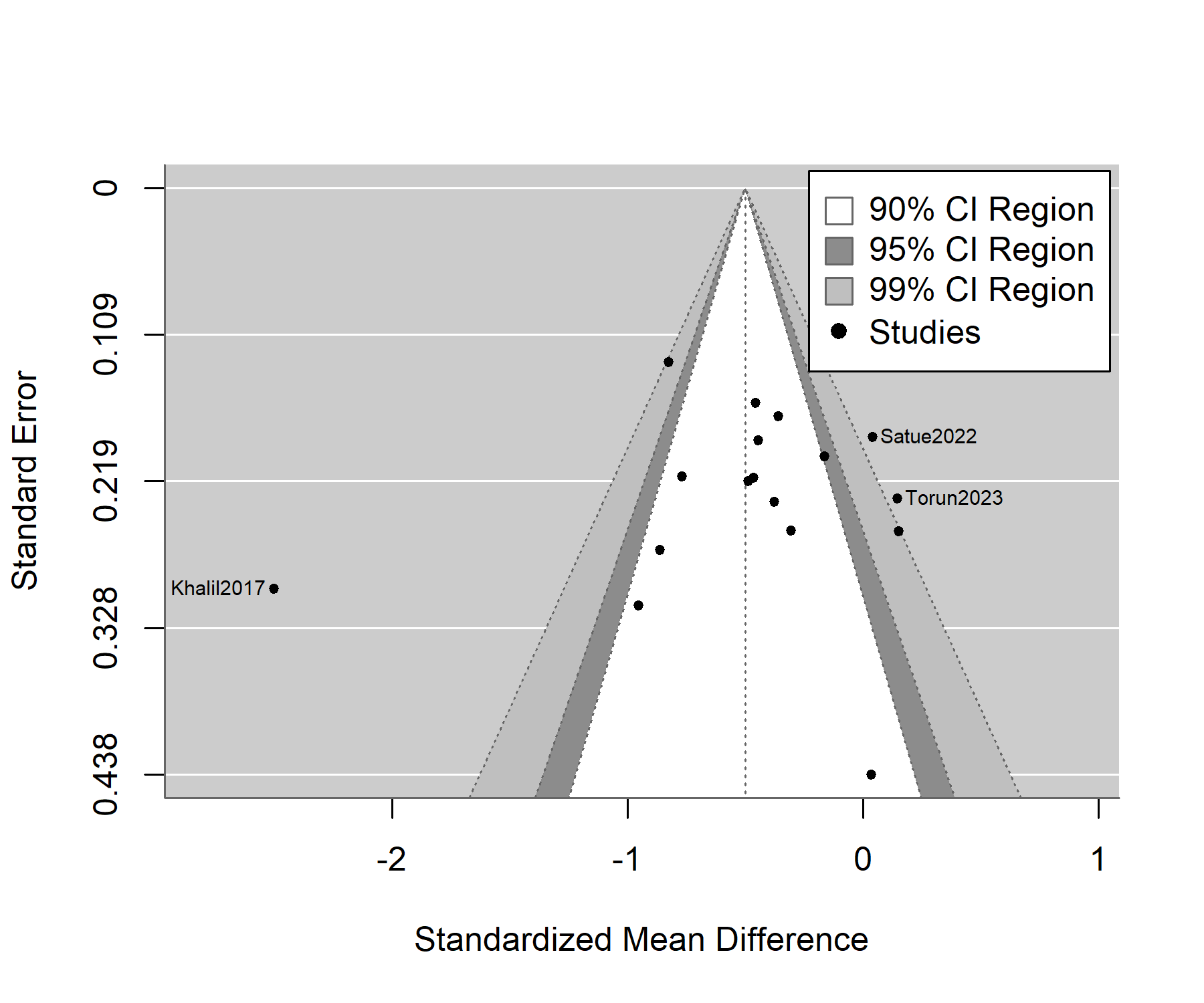

*Figure S 31 contour-enhanced funnel plot for bipolar disorder, pRNFL. A funnel plot shows the observed effect sizes or outcomes on the x-axis against some measure of precision of the observed effect sizes or outcomes on the y-axis, based on Sterne and Egger (2001). In the absence of publication bias and heterogeneity, one would then expect to see the points forming a funnel shape, with the majority of the points falling inside of the pseudo-confidence region (white). With publication bias present, second, empty and dashed triangle shows region one would expect studies to be if the null hypothesis was true (no difference between groups). Every dot is a study, with studies outside the pseudo-confidence-region identified by Name of the first author and year of publication.*

**Bipolar Disorder: pRNFL Quadrants**

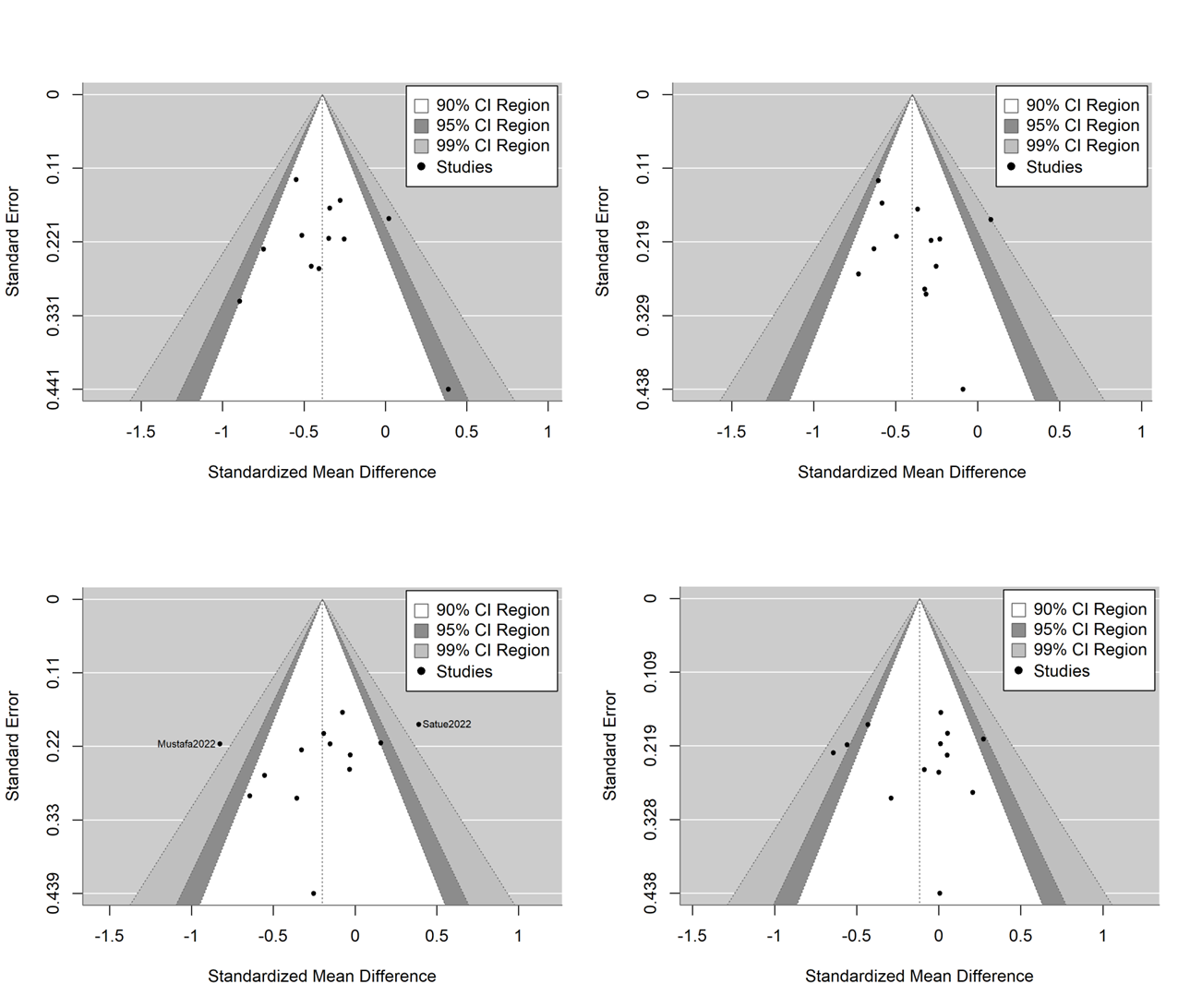

*Figure S 32 contour-enhanced funnel plots for bipolar disorder, pRNFL quadrants: Superior (upper left), inferior (upper right), nasal (lower left) and temporal (lower right).*

**Bipolar Disorder: Macula Thickness**

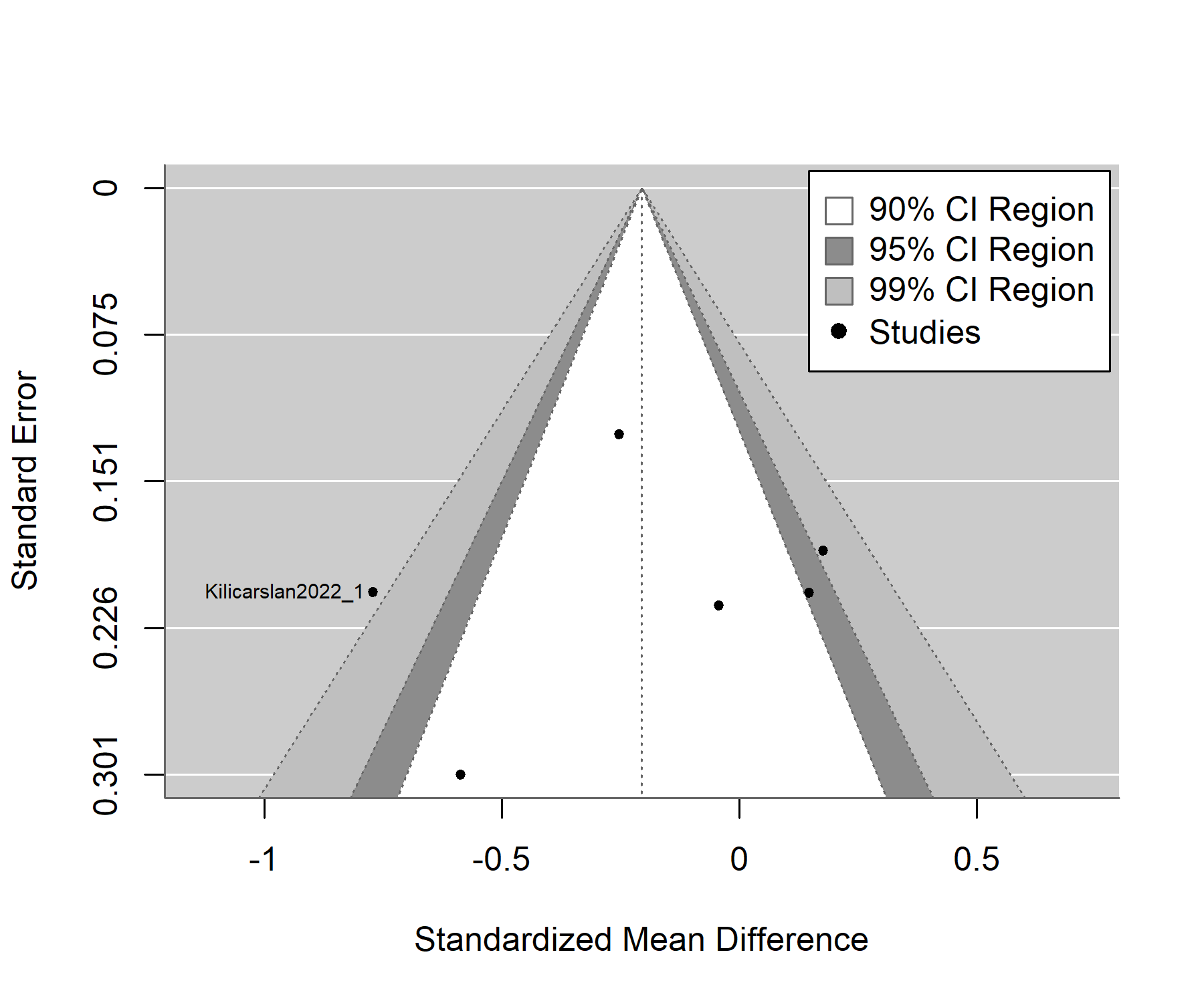

*Figure S 33 contour-enhanced funnel plot for bipolar disorder, macular thickness*

**Bipolar Disorder: Macula Subfields – Inner Ring**

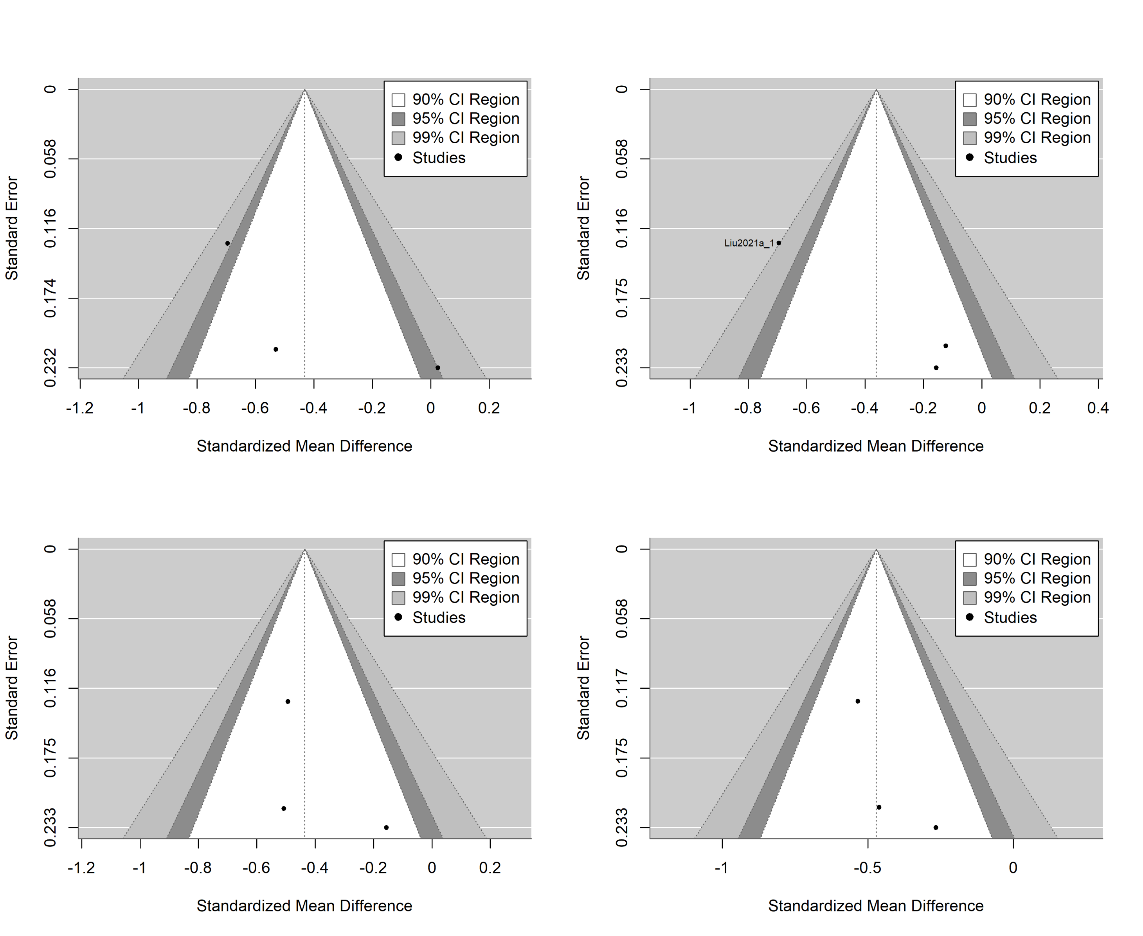

*Figure S 34 contour-enhanced funnel plots for bipolar disorder, macula subfields, inner ring: Superior (upper left), inferior (upper right), nasal (lower left) and temporal (lower right)*

**Bipolar Disorder: Macula Subfields – Outer Ring**

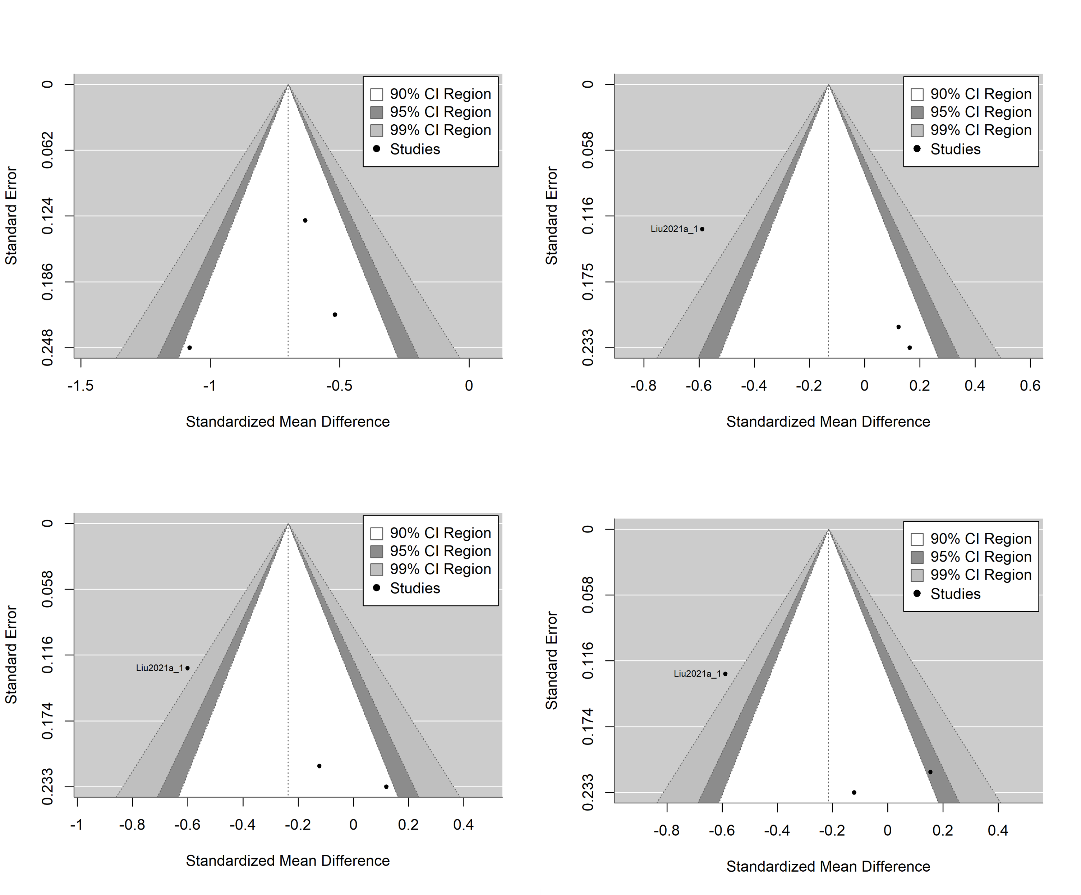

*Figure S 35 contour-enhanced funnel plots for bipolar disorder, macula subfields, outer ring: Superior (upper left), inferior (upper right), nasal (lower left) and temporal (lower right)*

**Major Depressive Disorder: pRNFL**

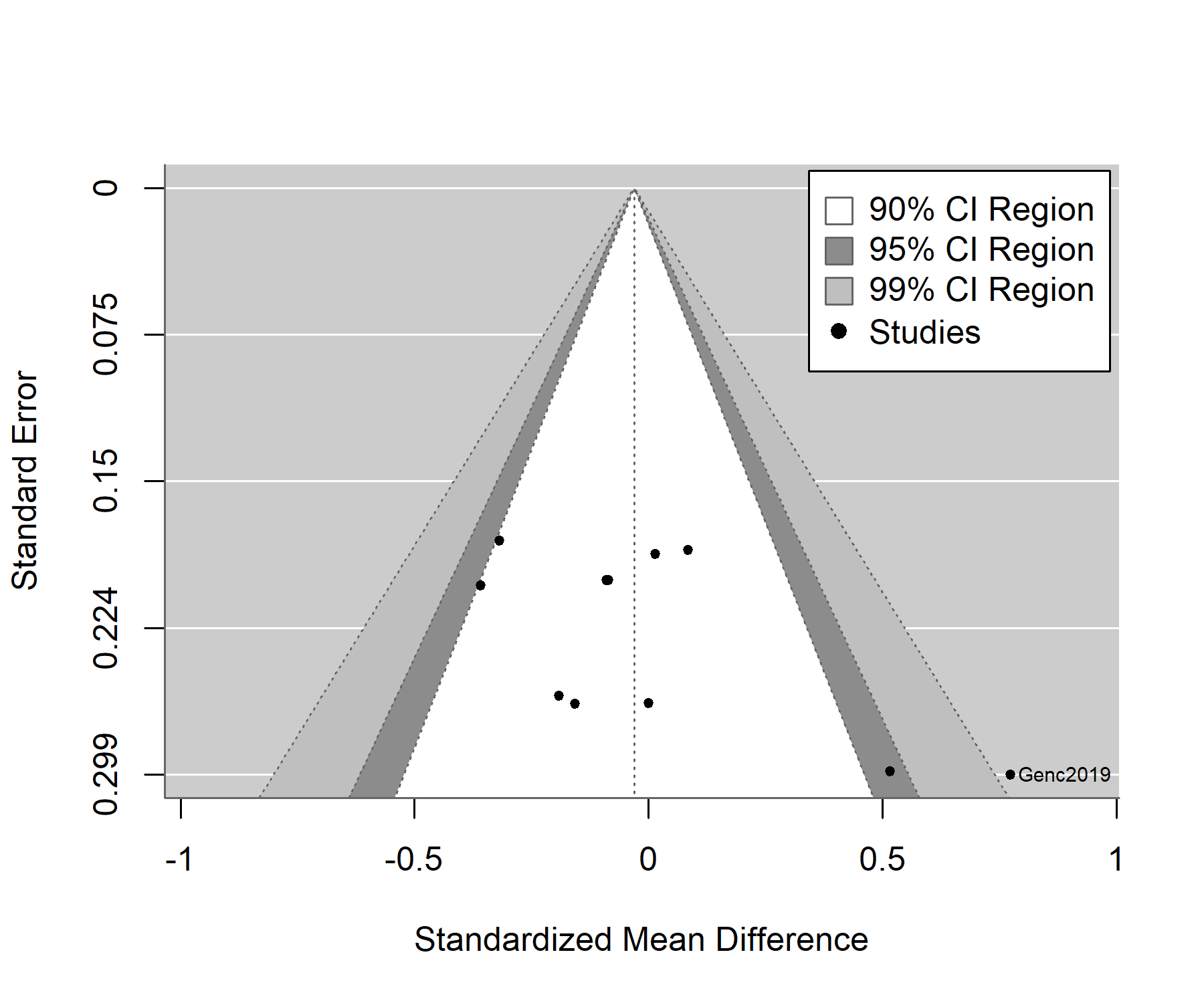

*Figure S 36 contour-enhanced funnel plot for major depressive disorder, RNFL. Only low-precision studies published.*

**Major Depressive Disorder RNFL quadrants**

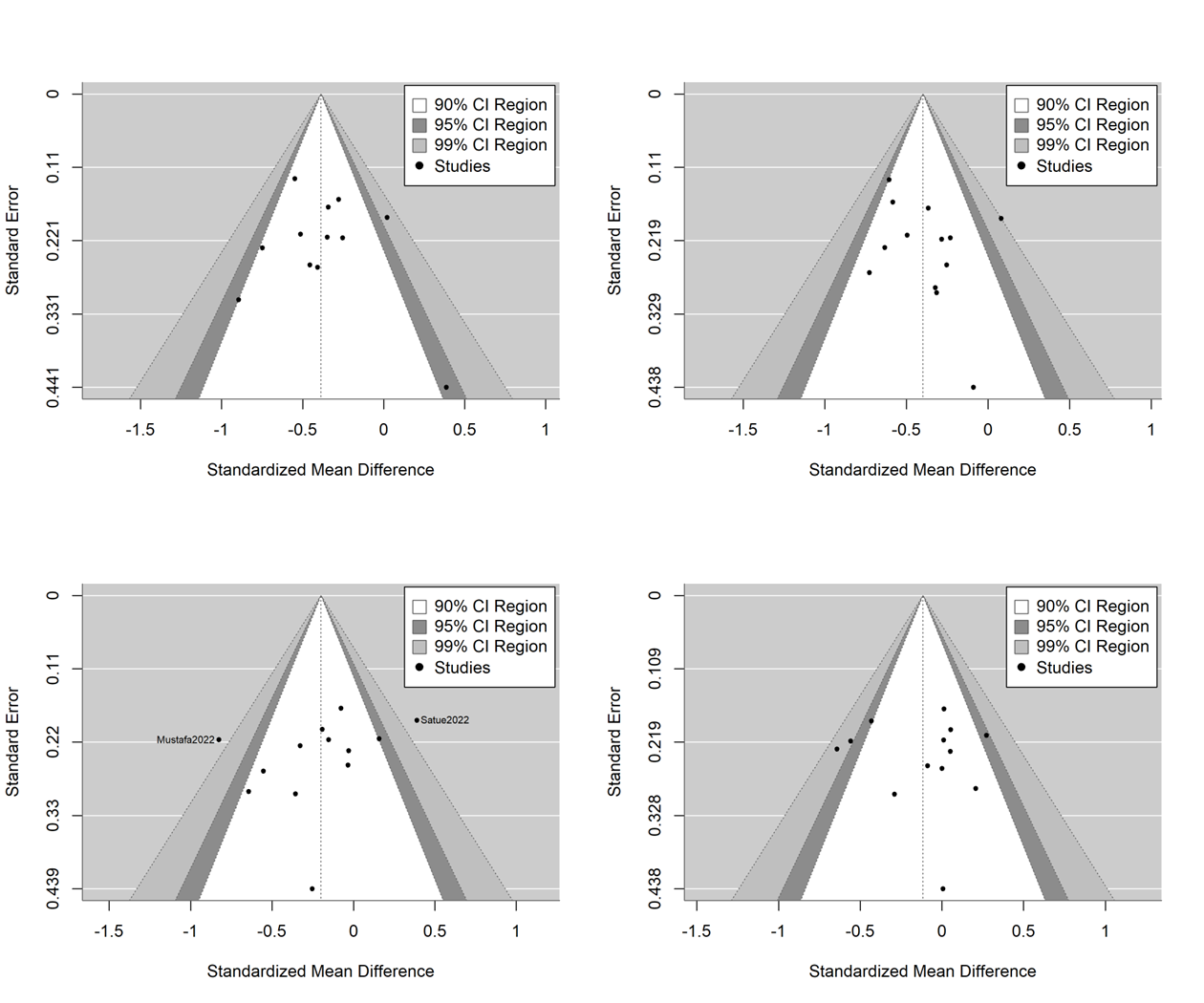

*Figure S 37 contour-enhanced funnel plots for major depressive disorder, pRNFL quadrants: Superior (upper left), inferior (upper right), nasal (lower left) and temporal (lower right)*

**Supplementary Tables 18 – 20: Data availability for Schizophrenia Spectrum Disorder, Biploar Disorder, and Major Depressive Disorder**

**Schizophrenia Spectrum Disorders**

*Table S 18 Data availability for retinal layers in Schizophrenia Spectrum Disorders. pRNFL = peripheral retinal nerve fiber layer. pRNFL quad. = pRNFL quadrants. MV = Macula volume. Macula sbfld = Macula subfield. Cf-Macula = central foveal Macula. GCL = Ganglion Cell Layer. IPL = inner plexiform layer. GCL-IPL = combined Ganglion Cell and Inner Plexiform Layer. GCC = Ganglion Cell Complex*

| **Key** | **eye** | **pRNFL** | **pRNFL quadrants** | **Macula** | **MV** | **Macular subfield** | **cfMacula** | **GCL** | **IPL** | **GCL-IPL** | **GCC** |
| --- | --- | --- | --- | --- | --- | --- | --- | --- | --- | --- | --- |
| Alizadeh2021 | pooled | x |  |  | x | x | x |  |  |  |  |
| Altun2020 | both | x | x | x |  | x | x |  |  |  |  |
| Asanad2021 | pooled | x | x |  |  | x | x |  |  | x |  |
| Ascaso2010 | pooled | x | x |  | x |  | x |  |  |  |  |
| Ascaso2015 | both | x | x |  | x |  | x |  |  |  |  |
| Bannai2020 | both | x |  |  |  |  |  | x | x |  |  |
| Boudriot2023 | pooled | x | x |  |  |  |  |  |  |  |  |
| Boudriot2024 | both |  |  | x |  | x | x | x |  | x |  |
| Bozali20222 | pooled | x | x |  |  |  | x |  |  |  |  |
| Budakoglu2021 | pooled | x | x |  |  |  |  |  |  |  |  |
| Carriello2023 | both | x |  | x | x |  | x |  |  |  |  |
| Celik2016 | pooled | x | x |  |  |  |  |  |  |  |  |
| Chu2012 | both | x | x |  | x |  |  |  |  |  |  |
| Daneshvar2024 | random | x |  |  |  |  |  |  |  |  |  |
| Delibas2018 | right |  | x |  |  |  |  |  |  |  |  |
| Domagala2023 | both | x |  | x | x |  |  |  |  |  | x |
| Gandu2021 | both | x |  | x |  |  |  |  |  | x | x |
| Hanifi2022 | right | x | x | x |  | x | x |  |  |  |  |
| Hosak2020 | both | x | x |  |  |  | x |  |  | x |  |
| Jerotic2020 | both |  | x |  |  |  |  |  |  |  |  |
| Jerotic2021 | both | x |  | x | x |  | x |  |  | x |  |
| Kango2022 | both |  |  |  | x |  | x | x |  |  |  |
| Kaya2022 | both | x | x |  | x |  |  |  |  | x |  |
| Khalil2022 | pooled | x | x |  |  |  |  |  |  |  | x |
| Koman2021 | pooled | x | x | x |  |  | x |  |  |  |  |
| Kurt2021 | both | x |  |  |  |  |  |  |  |  |  |
| Kurtulmus2020 | right | x |  | x |  |  |  | x | x |  |  |
| Kurtulmus2023 | both | x |  | x |  |  |  | x | x |  |  |
| Lai2020 | both | x |  |  | x |  | x |  |  | x |  |
| Lee2013 | right | x | x | x | x | x | x |  |  |  |  |
| Liu2020 | both | x |  | x |  | x | x |  |  |  |  |
| Liu2021b | both |  | x |  |  |  |  |  |  |  |  |
| Miller2020 | both | x |  |  | x |  |  |  |  | x |  |
| Mota2015 | pooled | x |  | x | x | x | x |  |  |  |  |
| Sarkar2021 | right | x | x |  |  | x | x |  |  |  |  |
| Schoenfeldt2020 | pooled | x |  |  | x |  |  |  |  |  |  |
| Silverstein2017 | both | x | x | x | x |  | x |  |  | x |  |
| Tasdelen2023 | both | x | x |  | x |  |  |  |  | x |  |
| Topcu2017 | na | x | x | x |  | x | x |  |  |  |  |
| Yesilkaya2023 | right | x | x |  |  |  |  |  |  |  | x |
| Yilmaz2016 | pooled | x | x | x |  | x |  |  |  |  |  |

**Bipolar Disorder**

*Table S 19 Data availability for retinal layers in Bipolar Disorder. pRNFL = peripheral retinal nerve fiber layer. pRNFL quad. = pRNFL quadrants. MV = Macula volume. Macula sbfld = Macula subfield. Cf-Macula = central foveal Macula. GCL = Ganglion Cell Layer. IPL = inner plexiform layer. GCL-IPL = combined Ganglion Cell and Inner Plexiform Layer. GCC = Ganglion Cell Complex.*

| **Key** | **eye** | **pRNFL** | **pRNFL quadrants** | **Macula** | **MV** | **Macular subfield** | **cfMacula** | **GCL** | **IPL** | **GCL-IPL** | **GCC** |
| --- | --- | --- | --- | --- | --- | --- | --- | --- | --- | --- | --- |
| Alici2019 | pooled | x | x |  |  |  |  | x |  |  |  |
| Altun2020 | both | x |  |  |  | x | x |  |  |  |  |
| Ayik2022 | both | x | x |  | x |  |  |  |  | x |  |
| Garcia2019 | random | x | x | x | x | x | x | x | x |  |  |
| Gokcinar2020 | random | x | x |  |  |  |  |  |  |  | x |
| Kalenderoglu2016a | pooled | x | x |  |  |  |  |  |  |  |  |
| Khalil2017 | both | x | x |  |  |  |  |  |  | < | x |
| Kilicarslan2022 | both | x |  | x |  |  | x |  |  |  |  |
| Koman2021 | pooled | x | x | x |  |  | x |  |  |  |  |
| Kurt2023 | both | x |  | x |  |  |  | x |  |  |  |
| Liu2021a | both | x | x | x |  | x | x |  |  |  |  |
| Mehraban2016 | pooled | x | x |  |  |  |  |  |  |  |  |
| Mustafa2022 | right | x | x | x |  |  |  |  |  | x |  |
| Özgedik2024 | right | x |  |  |  |  |  | x | x |  |  |
| Polo2019 | random | x | x | x |  |  |  |  |  |  |  |
| Sanchez2021 | random |  | x |  |  |  |  |  |  |  |  |
| Satue2022 | random | x | x |  |  |  |  |  |  |  |  |
| Torun2023 | right | x |  |  |  |  | x | x |  |  |  |

**Major Depressive Disorder**

*Table S 20 Data availability for retinal layers in major depressive disorder . pRNFL = peripheral retinal nerve fiber layer. pRNFL quad. = pRNFL quadrants. MV = Macula volume. Macula sbfld = Macula subfield. Cf-Macula = central foveal Macula. GCL = Ganglion Cell Layer. IPL = inner plexiform layer. GCL-IPL = combined Ganglion Cell and Inner Plexiform Layer. GCC = Ganglion Cell Complex.*

| **Key** | **eye** | **pRNFL** | **pRNFL quadrants** | **Macula** | **MV** | **Macular subfield** | **cfMacula** | **GCL** | **IPL** | **GCL-IPL** | **GCC** |
| --- | --- | --- | --- | --- | --- | --- | --- | --- | --- | --- | --- |
| Genc2019 | both | x | x |  |  |  |  |  |  |  |  |
| Jung2020 | pooled | x | x |  |  |  |  |  |  | x |  |
| Kalenderoglu2016 | pooled | x |  |  |  |  |  |  |  |  |  |
| Liu2021a | both | x | x | x |  | x | x |  |  |  |  |
| Liu2022 | pooled | x | x | x |  |  | x |  |  |  | x |
| Lubinski2023 | pooled | x |  |  |  |  |  |  |  |  |  |
| Schoenfeldt2017 | pooled | x |  |  |  |  |  |  |  | x |  |
| Soenmez2017 | both | x |  |  |  |  |  |  |  |  |  |
| Xiao2024 | pooled | x |  |  |  |  |  |  |  |  |  |
| Yildiz2016 | right | x | x | x | x |  | x |  |  | x |  |
